# Efficacy and Safety of Different Inhaler Types for Asthma and Chronic Obstructive Pulmonary Disease. A Systematic Review and Meta-Analysis

**DOI:** 10.1101/2025.06.01.25328719

**Authors:** Michael J. Loftus, Miranda S. Cumpston, Shannon Barnes, John Blakey, Allan Glanville, Steve McDonald, Loyal Pattuwage, Megan Rees, Rachel Silk, Heath White, Tari Turner, Karin Leder

**Author notes:** Corresponding Author: Dr Michael Loftus, Research Fellow, Division of Planetary Health, School of Public Health and Preventive Medicine, Monash University, Level 4, 553 St Kilda Road, Melbourne 3004 Australia. MJL and MSC are joint first authors. TT and KL are joint senior authors.

## Abstract

Pressurised metered dose inhalers (pMDIs) contain propellant gases with high global warming potential yet remain a cornerstone of management for asthma and chronic obstructive pulmonary disease (COPD). The aim of this study was to determine whether non-propellant alternatives of dry powder inhalers (DPIs) and soft mist inhalers (SMIs) had similar efficacy and safety. A systematic review and meta-analysis for differences in clinical outcomes and safety measures between devices was performed. No statistically significant or clinically important differences were found between inhaler types for any assessed measure. For asthma maintenance, the mean difference in peak expiratory flow rate between groups was 0.99L/min (95% confidence interval [CI] -1.11 to 3.09). For COPD, the mean difference in FEV_1_ between groups was 0.01L (95% CI -0.01 to 0.02). While the choice of optimal inhaler for an individual patient is a multifaceted decision, this review provides reassurance that non-pMDI devices can perform equally well.

## Introduction

Asthma and chronic obstructive pulmonary disease (COPD) are highly prevalent globally, affecting hundreds of millions of individuals.^1,2^ A cornerstone of therapy for these conditions is the directed delivery of effective medications via inhalers.^3,4^ Pressurised metered dose inhalers (pMDIs) are one of the most frequently used inhaler devices, with almost one billion manufactured every year.^5^ Current pMDIs contain fluorinated propellant gases (F-gases) which are potent greenhouse gases – they exert an effect thousands of times greater than the equivalent volume of carbon dioxide (CO_2_). For example, a single inhaler of a common long-acting beta-agonist and steroid combination product, using the propellant HFA-227a, has the same global warming potential as 36.5kg CO_2_ (equivalent to driving almost 200 kilometres in a petrol car).^6^ The healthcare sector contributes 4-5% of global greenhouse emissions,^7,8^ and the proportionally large contribution of inhalers (particularly pMDIs) is increasingly recognised. In the United Kingdom, pMDIs alone are responsible for around 3% of the entire National Health Service carbon footprint.^9^

A key component of this issue is the marked variation between countries in the proportion of prescribed inhalers that are pMDIs, due to both cultural and cost reasons. For example, there is a seven-fold difference in pMDI prescribing rates across northern European countries.^10^ These observations in the face of the climate crisis have led to a call for much greater use of propellant-free options such as dry powder inhalers (DPIs) and soft mist inhalers (SMIs), especially in countries that predominantly use pMDIs.^11^ A conscious consideration of the environmental impact is now included in inhaler guidelines in the UK.^12^ However, some uncertainty remains as to whether there is equipoise between the benefit of pMDIs and propellant-free inhalers for management of asthma and COPD. Previous systematic reviews with a narrow focus have considered this question for different drug classes across both conditions; however, these reviews are now 20 or more years old and did not include many drugs, formulations (e.g. extrafine particles) and devices (e.g. SMIs) that are now used routinely.^13–16^ Some countries have made calls to move away from pMDIs when clinically safe to do so,^17^ however strong contemporary data are needed to support such decisions. We therefore undertook a comprehensive systematic review and meta-analysis to establish if there was any evidence of a difference in clinical effectiveness or safety of treating individuals with asthma or COPD acutely or for maintenance therapy with pMDIs versus propellant-free devices, when the drugs and doses administered were broadly equivalent.

## Methods

This review was not registered, but we developed a protocol before commencing the review, containing additional detail on the methods used and a complete list of changes made to the planned methods.^18^ This review is reported in accordance with the Preferred Reporting Items for Systematic Reviews and Meta-Analyses statement.^19^

### Search strategy

We searched PubMed, Embase and the Cochrane Central Register of Controlled Trials to 30 January 2024, as well as reference lists of systematic reviews identified in the search. Full search strategies are provided in the Supplementary Information (Section A).

### Eligibility criteria

#### Study designs

Only randomised controlled trials (RCTs) were eligible for inclusion. In accordance with the protocol, observational studies were initially identified in the search but not included in the review as sufficient RCTs were available. We excluded studies not written in English and crossover studies (due to insufficient data regarding washout periods in many studies).

#### Population

We included studies of patients of any age with confirmed asthma or COPD. Asthma studies were categorised as management of acute asthma episodes (e.g. presenting for emergency care) or asthma maintenance therapy. Results were analysed separately for these three conditions.

#### Intervention

We included studies comparing the delivery of equivalent inhaled medication(s) by either DPI or SMI (grouped together as non-pMDI inhalers) versus pMDI. Medications were considered ‘equivalent’ if they were from the same drug class (e.g. short-acting beta-agonist (SABA), or inhaled corticosteroid (ICS)) and given at comparable doses. ICS doses were categorised into ‘high,’ ‘medium’ and ‘low’ dose according to National Institute for Health and Care Excellence (NICE) guidelines.^20^ Non-ICS medication doses were classified as ‘standard’ based on comparative clinical therapeutic equivalence, and ‘high’ if a multiple of that standard dose was used.^3,4^ Studies were excluded if there were a different number of drugs in each arm (e.g. ICS-alone versus ICS/SABA combination). We permitted slight differences in the frequency of medication administration (e.g. one versus two doses per day) but excluded larger differences (e.g. once versus three or four doses per day) due possible differences in adherence and pharmacokinetic properties. There were no restrictions on which drugs participants were taking prior to the trial, or on lead-in or washout periods. Results were analysed together for all drugs and doses.

#### Outcomes

Our primary outcomes were physiological lung function measurements (forced expiratory volume in 1 second (FEV_1_) and peak expiratory flow rate (PEFR)), symptom control (any scale), quality of life (any scale), exacerbations (as defined by each study) and use of additional reliever medication (any measure). Secondary outcomes focused on safety, including mortality, overall or treatment-related adverse events (AEs) and serious adverse events (SAEs) as defined by each study. Studies were eligible if they reported one or more primary or safety outcomes.

We had initially planned to report hospital admissions and emergency room attendance separately, but these were frequently incorporated by our included studies into composite measures of adverse events or exacerbations, and so we have reported these composite outcomes in this review. We had also planned to include additional secondary measures of satisfaction, adherence to therapy and inhaler technique, but found that many studies measuring these factors did not address effectiveness or safety outcomes and were therefore ineligible for inclusion in the review. To avoid presenting misleading estimates of these outcomes from a subset of the available literature, we decided to exclude these outcomes.

Studies were excluded if the duration of follow up after treatment was less than 48 hours for COPD and asthma maintenance, but such studies were included for acute asthma exacerbations.

Minimal clinically important differences (MCIDs) for various outcomes in asthma and COPD were identified from the published literature where possible or otherwise determined by consensus of our Respiratory Experts; these are summarised in the Supplementary Information (Section B).

### Study selection and data extraction

Title and abstract screening and full-text review were conducted using Covidence software.^21^ All identified titles were initially screened by one of MJL, LP or HW, before all abstracts of papers deemed potentially relevant were independently screened by two authors (MJL and LP). These two authors then performed independent full-text reviews of all remaining papers. Any disputes were resolved by consensus, or by consultation with the review content experts (AG, JB, MR) where needed.

We extracted data including the study design, clinical condition of participants, inclusion of children and/or adults, inhaler design, drugs and doses used, use of double-dummy placebos, particle size (normal or extra fine), use of spacers, and outcome data. Using a standardised extraction form, one researcher (SB, MJL, LP or RS) extracted and a second researcher verified the data, with inconsistencies discussed until consensus was reached. To improve consistency of data extracted from figures, a web-based plot digitising software was used.^22^ No additional data were sought from authors of included studies. Where studies reported results at more than one time point, the latest time point was used. Different measures of the same outcome and measures taken at different time points were combined in the analysis where possible.

### Risk of Bias

Studies were assessed for risk of bias using the Cochrane Risk of Bias tool.^23^ Assessment was completed independently for all included studies by two of SB, LP and RS, and reviewed by a third author (MC). Studies assessed as high risk for any domain were assessed overall as being at high risk of bias. Studies assessed as unclear risk for three or more domains, with no high-risk domains, were assessed overall as being at unclear risk of bias. Studies assessed as unclear risk for two or fewer domains, with no high-risk domains, were assessed overall as being at low risk of bias.

### Data analysis

#### Data synthesis

Included studies were grouped by condition (asthma maintenance, asthma acute exacerbations and COPD) for analysis and the available outcome measures were tabulated for selection as described above. Where two or more studies within a group reported results for an outcome of interest, we performed meta-analysis using a random-effects model and inverse variance statistical methods using RevMan Web.^24^ Where possible, we reported dichotomous outcomes as risk ratios (RRs) with 95% confidence intervals, and continuous outcomes as mean differences (where all studies used the same outcome measure), or standardised mean differences (where studies used a variety of outcome measures), with 95% confidence intervals.

Where additional studies reported outcome measures or data that could not be included in the meta-analysis, effect estimates were calculated where possible and all results were reported in table format. No synthesis was performed on these results.

#### Imputation and standardisation

Where results were reported with no measure of variance, we imputed standard deviations (SDs) for group-level data from other studies that used the same outcome measure, using the average of SDs from the intervention and control groups of the study with the highest available SD values, as a conservative approach to err on the side of including a clinically important difference within the confidence intervals. Sensitivity analysis was conducted excluding studies with imputed data from the analysis, and in all cases no meaningful change on the overall meta-analysis results was observed. For standardised analyses, where studies used a range of outcome measures with opposing directions of effect (e.g. scales for which a lower score indicated improvement, and scales for which a higher score indicated improvement), results for the least frequent direction were multiplied by -1 in the analysis. Also, for standardised analyses, either change or endpoint scores were used for all included studies, based on which was the most frequently reported for that outcome. Where outcome data was converted from endpoint to change scores, appropriate measures of variance were also calculated where possible, or otherwise imputed as described above.

#### Heterogeneity and subgroup analyses

Heterogeneity was assessed using the I^2^ statistic. Exploration of the causes of heterogeneity was planned using subgroup analyses, including adults and adolescents (13+ years) vs children (up to 12 years), and SMI vs DPI inhalers. An additional subgroup analysis was conducted to explore the influence of study funding by the manufacturer of either the pMDI or non-pMDI inhalers, or neither. These subgroup analyses were conducted using the primary outcome of FEV1, which was commonly measured across the greatest number of studies. For the outcomes of AEs and SAEs, an additional subgroup analysis was conducted by whether studies employed a “double dummy” design. AEs relating to a drug delivery system itself (e.g. constituents of the propellant, or dry powder) and not the active agent would be more equally distributed in double dummy studies where all participants are exposed to both devices; this could have obscured a true difference between device types.

#### Certainty in the evidence

The possibility of reporting bias was considered in the context of the available studies. The presence of small study effects that may indicate reporting bias was assessed using funnel plots where 10 or more studies contributed to a meta-analysis.

We assessed certainty in the evidence using the Grading of Recommendations, Assessment, Development, and Evaluations (GRADE) approach,^25^ incorporating the risk of bias, imprecision, inconsistency, indirectness and reporting bias. We summarised assessments for main outcomes in ‘summary of findings’ tables and used standardised language to report the certainty of results.^26^

## Results

### Description of studies

Of 3106 records identified in the search and 51 records identified from references of systematic reviews, 42 RCTs (reported in 43 papers) were included (30 on asthma maintenance therapy, five on acute asthma exacerbations, and seven on COPD) (**Figure 1** and **Table 1**). All studies of acute asthma exacerbations were in children. The majority of studies (37/42, 88.1%) were funded by device manufacturers, including all seven studies assessing patients with COPD (**Table 1**). Among these 37 studies, in 23 (62.2%) the study received funding from the manufacturer of the DPI/SMI device, in 9 (24.3%) the study received funding from the manufacturer of the pMDI device, and in 5 (13.5%) the same manufacturer produced the devices used in both study arms and funded the study (**Table S14** in the Supplementary Information).

**Figure 1.**
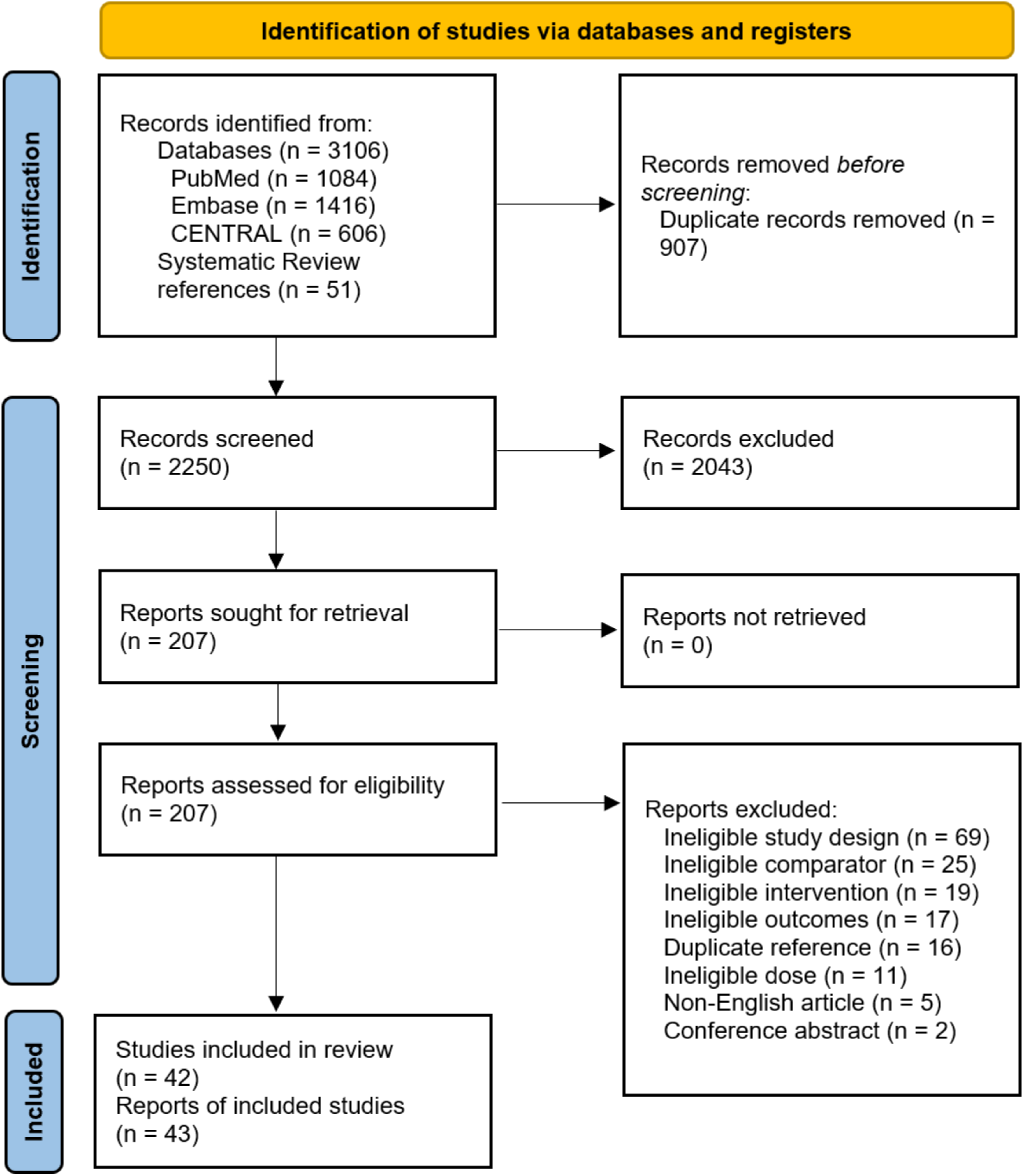
Flow diagram of the systematic review following the Preferred Reporting Items for Systematic Reviews and Meta-Analyses guidelines.

**Table 1.**
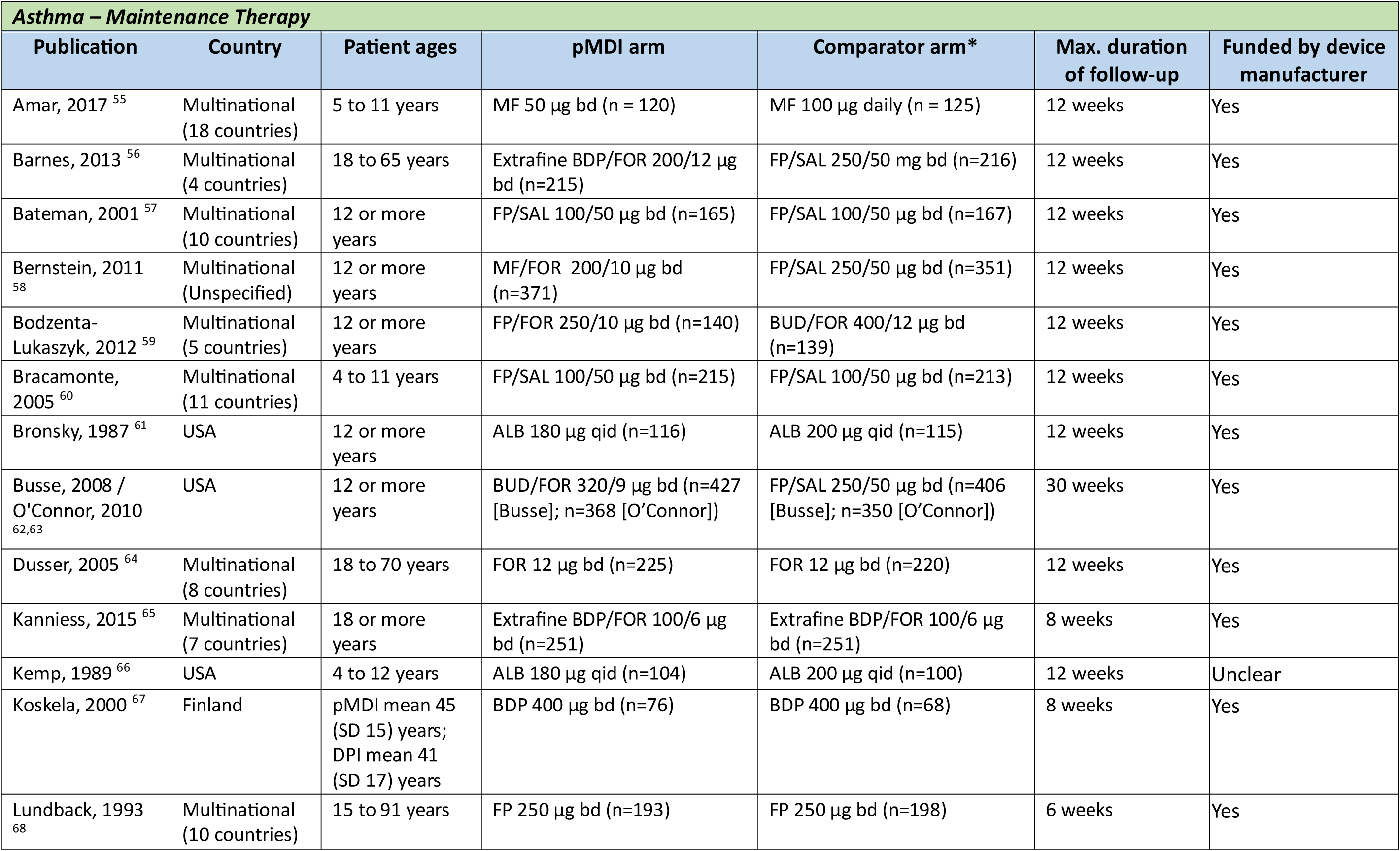
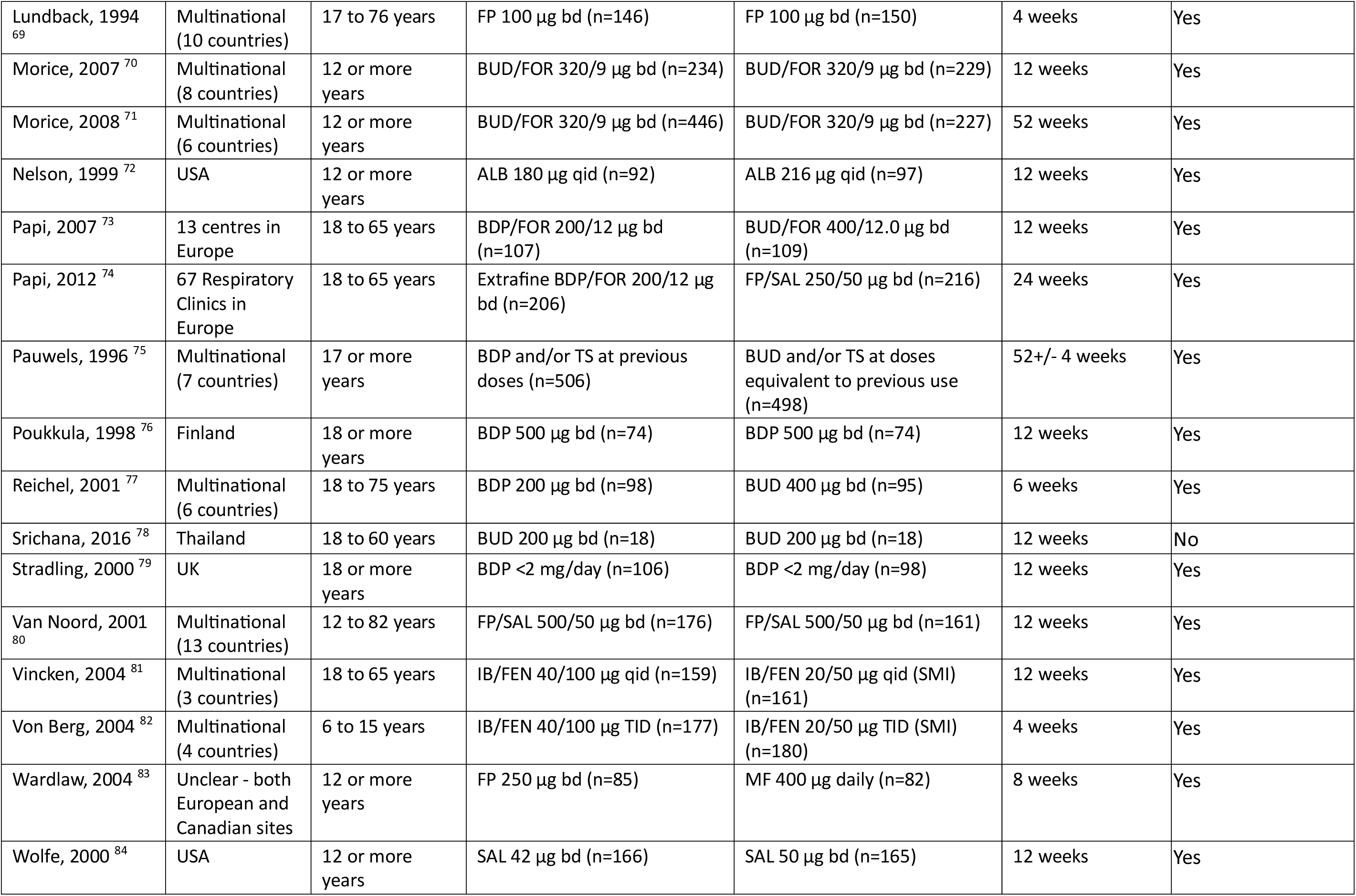
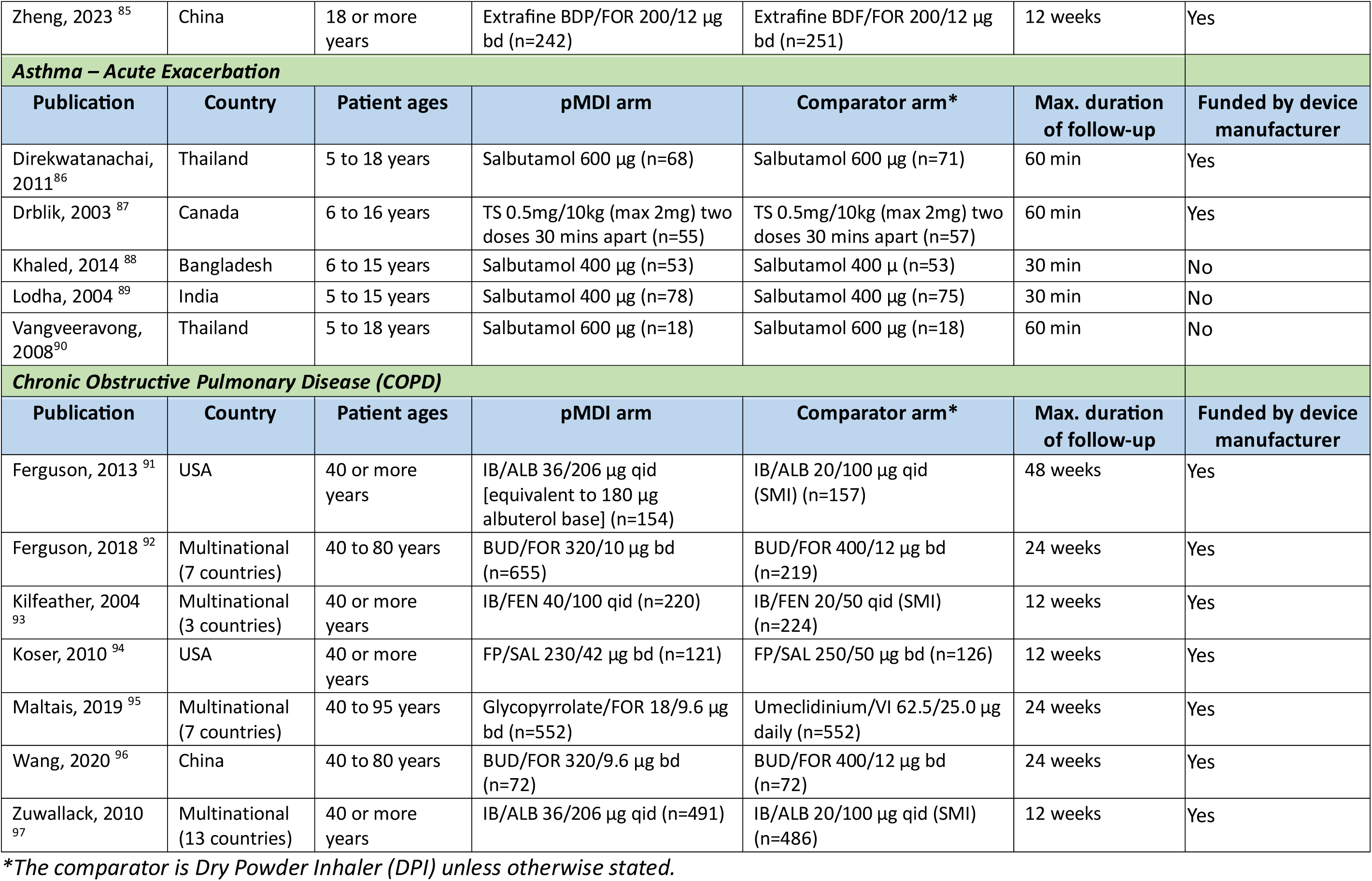
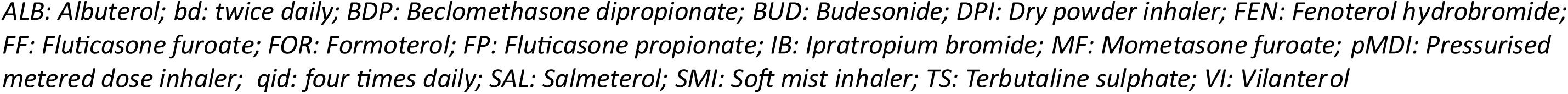
Characteristics of included studies.

| ***Asthma – Maintenance Therapy*** |  |  |  |  |  |  |
| --- | --- | --- | --- | --- | --- | --- |
| **Publication** | **Country** | **Patient ages** | **pMDI arm** | **Comparator arm\*** | **Max. duration of follow-up** | **Funded by device manufacturer** |
| Amar, 2017 <sup>55</sup> | Multinational (18 countries) | 5 to 11 years | MF 50 µg bd (n = 120) | MF 100 µg daily (n = 125) | 12 weeks | Yes |
| Barnes, 2013 <sup>56</sup> | Multinational (4 countries) | 18 to 65 years | Extrafine BDP/FOR 200/12 µg bd (n=215) | FP/SAL 250/50 mg bd (n=216) | 12 weeks | Yes |
| Bateman, 2001 <sup>57</sup> | Multinational (10 countries) | 12 or more years | FP/SAL 100/50 µg bd (n=165) | FP/SAL 100/50 µg bd (n=167) | 12 weeks | Yes |
| Bernstein, 2011 <sup>58</sup> | Multinational (Unspecified) | 12 or more years | MF/FOR 200/10 µg bd (n=371) | FP/SAL 250/50 µg bd (n=351) | 12 weeks | Yes |
| Bodzenta-Lukaszyk, 2012 <sup>59</sup> | Multinational (5 countries) | 12 or more years | FP/FOR 250/10 µg bd (n=140) | BUD/FOR 400/12 µg bd (n=139) | 12 weeks | Yes |
| Bracamonte, 2005 <sup>60</sup> | Multinational (11 countries) | 4 to 11 years | FP/SAL 100/50 µg bd (n=215) | FP/SAL 100/50 µg bd (n=213) | 12 weeks | Yes |
| Bronsky, 1987 <sup>61</sup> | USA | 12 or more years | ALB 180 µg qid (n=116) | ALB 200 µg qid (n=115) | 12 weeks | Yes |
| Busse, 2008 / O'Connor, 2010 <sup>62,63</sup> | USA | 12 or more years | BUD/FOR 320/9 µg bd (n=427 [Busse]; n=368 [O'Connor]) | FP/SAL 250/50 µg bd (n=406 [Busse]; n=350 [O'Connor]) | 30 weeks | Yes |
| Dusser, 2005 <sup>64</sup> | Multinational (8 countries) | 18 to 70 years | FOR 12 µg bd (n=225) | FOR 12 µg bd (n=220) | 12 weeks | Yes |
| Kanniess, 2015 <sup>65</sup> | Multinational (7 countries) | 18 or more years | Extrafine BDP/FOR 100/6 µg bd (n=251) | Extrafine BDP/FOR 100/6 µg bd (n=251) | 8 weeks | Yes |
| Kemp, 1989 <sup>66</sup> | USA | 4 to 12 years | ALB 180 µg qid (n=104) | ALB 200 µg qid (n=100) | 12 weeks | Unclear |
| Koskela, 2000 <sup>67</sup> | Finland | pMDI mean 45 (SD 15) years; DPI mean 41 (SD 17) years | BDP 400 µg bd (n=76) | BDP 400 µg bd (n=68) | 8 weeks | Yes |
| Lundback, 1993 <sup>68</sup> | Multinational (10 countries) | 15 to 91 years | FP 250 µg bd (n=193) | FP 250 µg bd (n=198) | 6 weeks | Yes |
| Lundback, 1994<sup>69</sup> | Multinational (10 countries) | 17 to 76 years | FP 100 µg bd (n=146) | FP 100 µg bd (n=150) | 4 weeks | Yes |
| Morice, 2007<sup>70</sup> | Multinational (8 countries) | 12 or more years | BUD/FOR 320/9 µg bd (n=234) | BUD/FOR 320/9 µg bd (n=229) | 12 weeks | Yes |
| Morice, 2008<sup>71</sup> | Multinational (6 countries) | 12 or more years | BUD/FOR 320/9 µg bd (n=446) | BUD/FOR 320/9 µg bd (n=227) | 52 weeks | Yes |
| Nelson, 1999<sup>72</sup> | USA | 12 or more years | ALB 180 µg qid (n=92) | ALB 216 µg qid (n=97) | 12 weeks | Yes |
| Papi, 2007<sup>73</sup> | 13 centres in Europe | 18 to 65 years | BDP/FOR 200/12 µg bd (n=107) | BUD/FOR 400/12.0 µg bd (n=109) | 12 weeks | Yes |
| Papi, 2012<sup>74</sup> | 67 Respiratory Clinics in Europe | 18 to 65 years | Extrafine BDP/FOR 200/12 µg bd (n=206) | FP/SAL 250/50 µg bd (n=216) | 24 weeks | Yes |
| Pauwels, 1996<sup>75</sup> | Multinational (7 countries) | 17 or more years | BDP and/or TS at previous doses (n=506) | BUD and/or TS at doses equivalent to previous use (n=498) | 52+/- 4 weeks | Yes |
| Poukkula, 1998<sup>76</sup> | Finland | 18 or more years | BDP 500 µg bd (n=74) | BDP 500 µg bd (n=74) | 12 weeks | Yes |
| Reichel, 2001<sup>77</sup> | Multinational (6 countries) | 18 to 75 years | BDP 200 µg bd (n=98) | BUD 400 µg bd (n=95) | 6 weeks | Yes |
| Srichana, 2016<sup>78</sup> | Thailand | 18 to 60 years | BUD 200 µg bd (n=18) | BUD 200 µg bd (n=18) | 12 weeks | No |
| Stradling, 2000<sup>79</sup> | UK | 18 or more years | BDP <2 mg/day (n=106) | BDP <2 mg/day (n=98) | 12 weeks | Yes |
| Van Noord, 2001<sup>80</sup> | Multinational (13 countries) | 12 to 82 years | FP/SAL 500/50 µg bd (n=176) | FP/SAL 500/50 µg bd (n=161) | 12 weeks | Yes |
| Vincken, 2004<sup>81</sup> | Multinational (3 countries) | 18 to 65 years | IB/FEN 40/100 µg qid (n=159) | IB/FEN 20/50 µg qid (SMI) (n=161) | 12 weeks | Yes |
| Von Berg, 2004<sup>82</sup> | Multinational (4 countries) | 6 to 15 years | IB/FEN 40/100 µg TID (n=177) | IB/FEN 20/50 µg TID (SMI) (n=180) | 4 weeks | Yes |
| Wardlaw, 2004<sup>83</sup> | Unclear - both European and Canadian sites | 12 or more years | FP 250 µg bd (n=85) | MF 400 µg daily (n=82) | 8 weeks | Yes |
| Wolfe, 2000<sup>84</sup> | USA | 12 or more years | SAL 42 µg bd (n=166) | SAL 50 µg bd (n=165) | 12 weeks | Yes |

| Zheng, 2023<sup>85</sup> | China | 18 or more years | Extrafine BDP/FOR 200/12 µg bd (n=242) | Extrafine BDF/FOR 200/12 µg bd (n=251) | 12 weeks | Yes |
| --- | --- | --- | --- | --- | --- | --- |
| **Asthma – Acute Exacerbation** |  |  |  |  |  |  |
| Publication | Country | Patient ages | pMDI arm | Comparator arm* | Max. duration of follow-up | Funded by device manufacturer |
| Direkwatanachai, 2011<sup>86</sup> | Thailand | 5 to 18 years | Salbutamol 600 µg (n=68) | Salbutamol 600 µg (n=71) | 60 min | Yes |
| Drblik, 2003<sup>87</sup> | Canada | 6 to 16 years | TS 0.5mg/10kg (max 2mg) two doses 30 mins apart (n=55) | TS 0.5mg/10kg (max 2mg) two doses 30 mins apart (n=57) | 60 min | Yes |
| Khaled, 2014<sup>88</sup> | Bangladesh | 6 to 15 years | Salbutamol 400 µg (n=53) | Salbutamol 400 µ (n=53) | 30 min | No |
| Lodha, 2004<sup>89</sup> | India | 5 to 15 years | Salbutamol 400 µg (n=78) | Salbutamol 400 µg (n=75) | 30 min | No |
| Vangveeravong, 2008<sup>90</sup> | Thailand | 5 to 18 years | Salbutamol 600 µg (n=18) | Salbutamol 600 µg (n=18) | 60 min | No |
| **Chronic Obstructive Pulmonary Disease (COPD)** |  |  |  |  |  |  |
| Publication | Country | Patient ages | pMDI arm | Comparator arm* | Max. duration of follow-up | Funded by device manufacturer |
| Ferguson, 2013<sup>91</sup> | USA | 40 or more years | IB/ALB 36/206 µg qid [equivalent to 180 µg albuterol base] (n=154) | IB/ALB 20/100 µg qid (SMI) (n=157) | 48 weeks | Yes |
| Ferguson, 2018<sup>92</sup> | Multinational (7 countries) | 40 to 80 years | BUD/FOR 320/10 µg bd (n=655) | BUD/FOR 400/12 µg bd (n=219) | 24 weeks | Yes |
| Kilfeather, 2004<sup>93</sup> | Multinational (3 countries) | 40 or more years | IB/FEN 40/100 qid (n=220) | IB/FEN 20/50 qid (SMI) (n=224) | 12 weeks | Yes |
| Koser, 2010<sup>94</sup> | USA | 40 or more years | FP/SAL 230/42 µg bd (n=121) | FP/SAL 250/50 µg bd (n=126) | 12 weeks | Yes |
| Maltais, 2019<sup>95</sup> | Multinational (7 countries) | 40 to 95 years | Glycopyrrolate/FOR 18/9.6 µg bd (n=552) | Umeclidinium/VI 62.5/25.0 µg daily (n=552) | 24 weeks | Yes |
| Wang, 2020<sup>96</sup> | China | 40 to 80 years | BUD/FOR 320/9.6 µg bd (n=72) | BUD/FOR 400/12 µg bd (n=72) | 24 weeks | Yes |
| Zuwallack, 2010<sup>97</sup> | Multinational (13 countries) | 40 or more years | IB/ALB 36/206 µg qid (n=491) | IB/ALB 20/100 µg qid (SMI) (n=486) | 12 weeks | Yes |

\*The comparator is Dry Powder Inhaler (DPI) unless otherwise stated.
*ALB: Albuterol; bd: twice daily; BDP: Beclomethasone dipropionate; BUD: Budesonide; DPI: Dry powder inhaler; FEN: Fenoterol hydrobromide; FF: Fluticasone furoate; FOR: Formoterol; FP: Fluticasone propionate; IB: Ipratropium bromide; MF: Mometasone furoate; pMDI: Pressurised metered dose inhaler; qid: four times daily; SAL: Salmeterol; SMI: Soft mist inhaler; TS: Terbutaline sulphate; VI: Vilanterol*

Risk of bias assessments are summarised in **Figure S1** in the Supplementary Information. Across all 42 studies, 21 were considered high-risk for bias, 5 were low-risk and the remaining 16 were unclear. Of those studies rated at high risk, the most frequent domain assessed as high risk of bias related to blinding of participants and personnel.

### Analyses

Therapy via either a pMDI or non-pMDI device had similar effects on all outcomes of interest. Most evidence was considered moderate certainty, with certainty downgraded due to the proportion of studies at high or unclear risk of bias, but the observed results were highly robust to all sensitivity analyses, and imprecision and heterogeneity were both very low across most outcomes. No indication of reporting bias was identified, and small study effects were not detected in funnel plots for any meta-analyses. Summary of findings tables including GRADE assessments of the certainty of the evidence are presented in the Supplementary Information (Section C).

#### Forced expiratory volume in 1 second (FEV_1_)

There was moderate certainty evidence of little or no difference in FEV_1_ between pMDI and non-pMDI devices for both asthma maintenance (standardised mean difference (SMD) 0.04, 95% CI -0.01 to 0.10; 27 studies; n = 9181) and COPD (SMD 0.03, 95% CI -0.03 to 0.09; seven studies; n = 3946) (**Figure 2**). In asthma maintenance, this equates to a mean difference in percent predicted FEV_1_ of 0.57% (95% CI -0.14% to 1.42%), well below the MCID of 20% ^27^ (see minimal clinically important differences in **Table S4** in the Supplementary Information). The effect estimate among COPD patients is equivalent to a mean difference of 0.01L (95% CI -0.01L to 0.02L), well below the MCID of 0.1L.^28^

**Figure 2.**
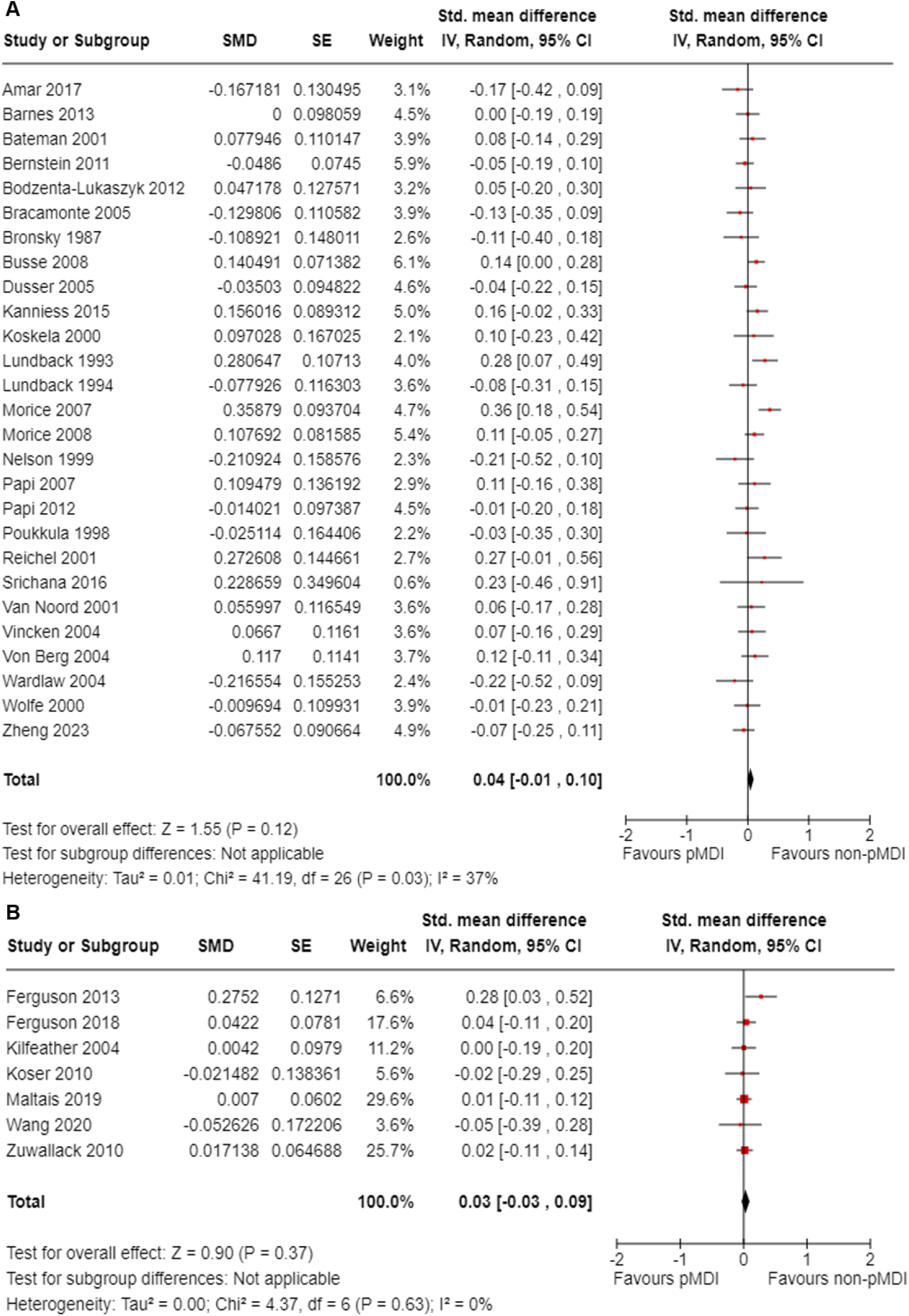
Meta-analysis of the association between device type and forced expiratory volume in 1 second (FEV_1_) in (A) asthma maintenance and (B) chronic obstructive pulmonary disease. The centre of the squares or diamonds indicates the point estimate and the width is the 95% confidence interval (CI). SE = standard error. SMD = standard mean difference. IV = inverse variance.

Only one study assessed FEV_1_ in acute asthma, which showed low certainty evidence of little to no difference in this population (mean difference 2% predicted, 95% CI -2.9% to 6.9%). One additional study reported FEV_1_ in asthma maintenance that could not be included in the meta-analysis (**Table S8** in the Supplementary Information).

#### Peak expiratory flow rate (PEFR)

There was moderate certainty evidence of little or no difference in PEFR between pMDI and non-pMDI devices for asthma maintenance (mean difference 0.99L/min, 95% CI -1.11 to 3.09; 24 studies; n = 8083) (**Figure 3**), with these values well short of the MCID of 18.8L/min.^29^

**Figure 3.**
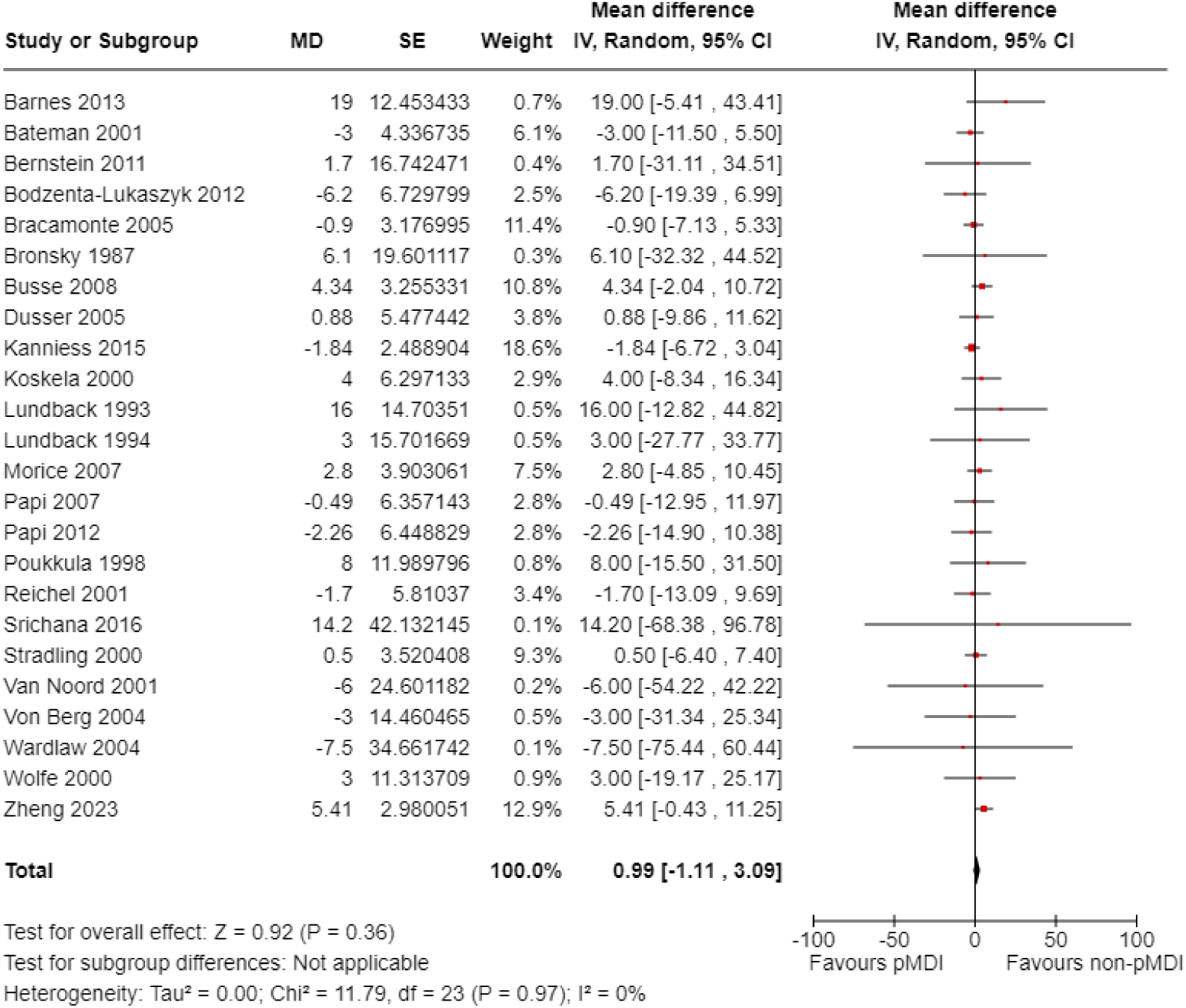
Meta-analysis of the association between device type and peak expiratory flow rate (PEFR) in asthma maintenance. The centre of the squares or diamond indicates the point estimate and the width is the 95% confidence interval (CI). SE = standard error. MD = mean difference. IV = inverse variance.

There was low certainty evidence of little or no difference between device types for acute asthma exacerbations (mean difference 1.38L/min, 95% CI -17.97 to 20.73; two studies; n = 259). This difference is smaller than the MCID used of 12% predicted (or approximately 25L/min in the populations studied). There was moderate certainty evidence of little or no difference among COPD patients (mean difference -2.56L/min, 95% CI -9.17 to 4.05; two studies; n = 644) (**Figure S3** in the Supplementary Information).

There was one additional study that measured PEFR in asthma maintenance that could not be included in the meta-analysis (**Table S9** in the Supplementary Information).

#### Symptom control

There was moderate certainty evidence of little or no difference in use of reliever medication between pMDI and non-pMDI devices for both asthma maintenance (SMD 0.02, 95% CI - 0.06 to 0.09; 13 studies; n = 4308) and COPD (mean difference -0.21 puffs/day, 95% CI = -0.51 to 0.10; three studies; n = 1265) (**Figure S11** in the Supplementary Information). The asthma maintenance SMD of 0.02 equates to approximately 0.05 puffs/day (95% CI -0.16 to 0.23), well below the minimal patient perceivable improvement value of 0.81 puffs/day.^29^

There were six additional studies that reported results on reliever use for asthma maintenance that could not be included in the meta-analysis (**Table S13** in the Supplementary Information). This was not a relevant outcome for people with acute asthma exacerbations.

Regarding symptom control scores, for asthma maintenance there was moderate certainty evidence of little or no difference between pMDI and non-pMDI devices (SMD -0.04, 95% CI - 0.11 to 0.02; eight studies; n = 3836). This SMD of -0.04 equates to approximately -0.02 points (95% CI -0.05 to 0.01) on the Asthma Control Questionnaire (ACQ-7), well below the MCID of 0.5 points. For COPD, using the COPD Assessment Test (CAT) score, there was moderate certainty evidence of little or no difference between pMDI and non-pMDI devices (mean difference -0.59 points, 95% CI = -1.19 to 0.01, one study; n = 1006). The observed mean difference was below the MCID of 2 points (**Figure S10** in the Supplementary Information).

For acute asthma exacerbations, using the Modified Wood Clinical Asthma Score there was low certainty evidence of little or no difference between pMDI and non-pMDI devices (mean difference -0.1 points, 95% CI -0.72 to 0.52, one study; n = 32) (**Figure S10** in the Supplementary Information).

There were six additional studies for asthma maintenance and one additional study for acute asthma exacerbations that reported symptom control results that could not be included in the meta-analysis (**Table S12** in the Supplementary Information).

#### Quality of life

Comparisons between studies were challenging due to the variety of quality of life scores used across studies. Meta-analysis was only possible for two asthma maintenance studies assessing the proportion of participants with an Asthma Quality of Life Questionnaire score improving ≥0.5 points. There was very low certainty evidence on the effect of device type on quality of life (relative risk (RR) 1.02, 95% CI 0.91 to 1.14; two studies; n = 871) (**Figure S9** in the Supplementary Information). Six further studies reported on quality of life measures (four for asthma maintenance, two for COPD) (**Table S11** in the Supplementary Information).

#### Exacerbations

There was moderate certainty evidence of little or no difference between pMDI and non-pMDI devices in the risk of experiencing ≥1 exacerbation for both asthma maintenance (RR 0.88, 95% CI 0.73 to 1.07; 17 studies; n = 6755) and COPD (RR 1.08, 95% CI 0.94 to 1.24; seven studies; n = 4101) (**Figure S4** in the Supplementary Information). This was not a relevant outcome for people with acute asthma exacerbations.

#### Adverse events

There was moderate certainty evidence of little or no difference between pMDI and non-pMDI devices in the risk of experiencing ≥1 AEs for asthma maintenance (RR 0.97, 95% CI 0.93 to 1.02; 23 studies; n = 7946), and low certainty evidence of little or no difference between these groups for COPD (RR 1.02, 95% CI 0.94 to 1.11; seven studies; n = 4106) (**Figure 4**). The evidence around the rate of AEs in acute asthma was very low certainty – it was only reported by one small RCT (RR 0.33, 95% CI 0.04 to 2.91, one study; n = 36) (**Figure S5** in the Supplementary Information).

**Figure 4.**
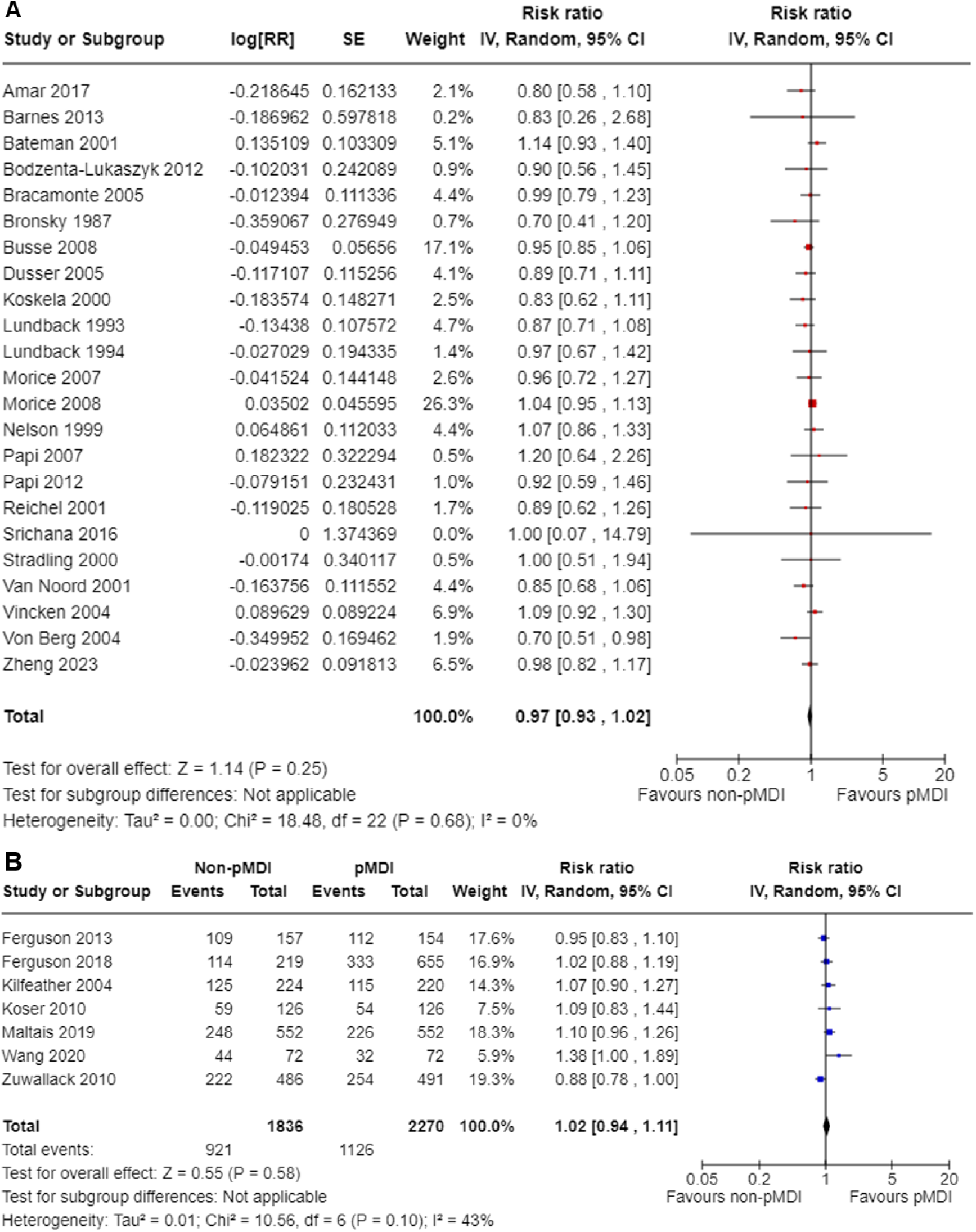
Meta-analysis of the association between device type and adverse events in (A) asthma maintenance and (B) chronic obstructive pulmonary disease. The centre of the squares or diamonds indicates the point estimate and the width is the 95% confidence interval (CI). RR = risk ratio. SE = standard error. IV = inverse variance.

Further assessments of SAEs, as well as only those AEs or SAEs that were deemed to be treatment-related, did not demonstrate clinically important differences between pMDI and non-pMDI devices for any condition. It should be noted that for each narrower definition of adverse events examined, fewer studies were included in the analysis and fewer events reported, increasing the imprecision of the estimates and reducing the certainty in the estimate (**Figures S6-S8** in the Supplementary Information).

#### Mortality

No deaths were reported in any studies of asthma maintenance or acute asthma. For COPD, there was low certainty evidence of little or no difference between pMDI and non-pMDI devices (RR 1.40, 95% CI 0.59 to 3.32; six studies; n = 3657) (**Figure S12** in the Supplementary Information). This translates to an absolute difference in deaths of three more per thousand, which did not meet our MCID of 1%.

#### Subgroup analyses

Subgroup analyses did not identify any factors that significantly modified the results, as little or no heterogeneity was observed. Studies in either children or adults were available for subgroup analysis by age group in asthma maintenance (**Figure S13** in the Supplementary Information, test for subgroup difference P = 0.4). Studies using either SMI or DPI were available for subgroup analysis by non-pMDI device type in COPD (**Figure S14** in the Supplementary Information, test for subgroup difference P = 0.48). Studies funded by the manufacturer of either the pMDI or non-pMDI device were available for subgrouping in asthma maintenance (**Figure S15** in the Supplementary Information, test for subgroup difference P = 0.32). Finally, subgroup analyses were conducted of AEs and SAEs by the presence or absence of double-dummy study design (**Figure S16** and **S17** in the Supplementary Information, test for subgroup difference range P = 0.24 to P = 0.75).

## Discussion

In this systematic review of 42 studies, we have found consistent evidence of no clinically meaningful difference between pMDIs and non-pMDI devices (DPIs and SMIs) in the management of asthma and COPD. The certainty of evidence was mainly rated as moderate rather than high due to the presence of some studies with high or unclear risk of bias and the possibility that biases could be operating in the direction of demonstrating equivalence. However, the overwhelming consistency of our findings and the robustness of these results to sensitivity analyses provides a clear conclusion. Results were uniform across FEV_1_ and PEFR measurements, disease control (exacerbations and reliever use) and adverse events. Our results are consistent with more focused previous systematic reviews around two decades ago that also showed no differences between devices regarding clinical effectiveness and adverse events.^13–16^ Importantly, our study contains updated data from newer studies, new drugs (or drug combinations), as well as a new device type (soft mist inhaler) that were not included in the previous reviews.

Climate change is the greatest global health threat of the twenty-first century,^30^ and patients with respiratory diseases are projected to be affected disproportionately.^31^ Globally, both governments and industry have made commitments to develop more sustainable healthcare systems, with a strong focus on decarbonisation. Increasing the proportion of inhalers without hydrofluorocarbons is an immediate step that can be taken to begin realising these ambitions. Our review supports that such a change may be possible without compromising patient care. However, we acknowledge that there may be further issues relating to device selection and/or switching at an individual level, which are beyond the scope of this review and cannot be directly answered by our research findings.

Selecting the most appropriate inhaler device for an individual patient is a complicated and multifaceted decision. This choice will be influenced by factors including the available agents (e.g. extrafine therapies are only widely available in pMDIs), patient familiarity and preference, patient ability and dexterity, as well as cost and accessibility.^32^ Increasingly, clinicians and patients are also being encouraged to consider the environmental impact of inhaler selection in position statements from peak bodies^33^ and decision aids.^34^ There are important ethical issues around potentially trading patient preference for population health,^35^ and avoiding any shaming of individuals who continue with pMDIs for strong user or disease reasons.^36^ Nevertheless, multiple surveys have shown that for most respiratory patients the environmental impact of their inhaler is an important consideration and one that could lead them to seek a device change.^37–39^

When contemplating a widespread move away from pMDIs to other devices (primarily DPIs), two potential concerns should be considered. The first concern is the suitability of DPIs for subgroups with limited lung function (e.g. the very young, the very old, or those experiencing an exacerbation), due to the need to generate sufficient inspiratory flow for adequate drug delivery.^40^ However, studies have shown that the majority of patients, including hospitalised patients nearing discharge, can achieve sufficient inspiratory flow to use low-resistance DPIs.^41–43^ A recent systematic review focusing of DPIs for primary school aged children with asthma found that the majority of children could use DPIs with adequate training and support.^44^ Additionally, inhaler use errors – a key contributor to inadequate drug delivery – appear more common with pMDIs,^45^ while there was no difference between devices in the rates of critical errors.^46^ Nevertheless, there are likely subpopulations for whom a pMDI will be the most appropriate device based on considerations of flow or other technical factors. The second concern is the potential negative impact of changing device type on disease control. Any switching of individuals between devices requires investment of time in education and training. Outcomes may be worse if this is not undertaken, but large-scale real-world evidence suggests it can be undertaken successfully.^47,48^ Changing inhaler devices does not have a predictable effect on outcomes,^49^ and large-scale switching for non-clinical reasons may lead to worsened disease control.^50^ Avoiding a loss of disease control is important not only for individual patient benefit, but also for sustainability. The greatest environmental impact of respiratory care comes from poor disease control leading to frequent use of reliever medications, unscheduled healthcare attendance and hospital admissions.^51^ A preventer inhaler that an individual can and will use is therefore preferable to one which is “greener” but remains in the cupboard.

Some strengths of our research are the large number of included studies, and the robust methodologies that were employed. Our research does have some limitations. First, to ensure that solely the inhaler device was being compared, we only included studies with equivalent drugs and doses in each arm – this limited the pool of available studies but does increase the confidence in our findings. Second, we acknowledge that participants in clinical trials are not wholly representative of the wider population with asthma or COPD, which potentially casts some doubt on the ability to generalise these findings. However, limiting our assessment to RCTs helped to reduce unmeasured confounders and other influences on results. Additionally, “real world” trials and analyses of primary care databases have also shown non-inferiority of dry powder inhalers in broader populations “treated as” asthma or COPD, giving greater support to our review’s conclusions.^47,48,52,53^ Third, a number of pharmaceutical companies are presently developing new propellants with lower global warming potential.^54^ While these propellants will not alter the validity of our systematic review’s findings, they may reduce the impetus to move patients away from pMDIs for environmental reasons.

The safest and most productive way to implement greater use of inhalers that do not contain propellants with a high global warming potential is uncertain. This review provides reassurance to patients, clinicians, and researchers that – at least based on RCT data – DPIs and SMIs are equivalent to pMDIs when administering corresponding drugs and doses. It lays a strong basis upon which to undertake future studies assessing optimal strategies to reduce the overall environmental impact of inhaler therapy.

## Author Contribution Statement

MJL: conceptualization, methodology, investigation, formal analysis, writing – original draft, writing – review and editing. MSC: methodology, investigation, visualisation, formal analysis, writing – original draft, writing – review and editing. SB: investigation, writing – review and editing. JB: conceptualization, methodology, supervision, writing – original draft, writing – original draft, writing – review and editing. AG: conceptualization, methodology, supervision, writing – original draft, writing – original draft, writing – review and editing. SM: methodology, investigation, writing – review and editing. LP: investigation, writing – review and editing. MR: conceptualization, methodology, supervision, writing – original draft, writing – original draft, writing – review and editing. RS: investigation, writing – review and editing. HW: methodology, investigation, writing – review and editing. TT: conceptualization, methodology, supervision, writing – review and editing. KL: conceptualization, methodology, supervision, writing – original draft, writing – original draft, writing – review and editing. All authors (except AG, who died in December 2024) read and approved the final manuscript.

## Data availability

Our research protocol is publicly available on the Monash University research repository (https://doi.org/10.26180/26065789.v2). The datasets generated and/or analysed during the current study are also available on the Monash University research repository (https://doi.org/10.26180/28916777).

## Acknowledgements

Michael Loftus is supported by a Royal Australasian College of Physicians (RACP) Research Establishment Fellowship. Miranda Cumpston is supported by philanthropic research funding from the Walter Thomas Cottman Charitable Trust and The Phyllis Connor Memorial Trust, managed by Equity Trustees. The funders played no role in study design, data collection, analysis and interpretation of data, or the writing of this manuscript. Thanks to Dr Kim Jachno of the Australian Living Evidence Collaboration in the School of Public Health and Preventive Medicine, Monash University, for statistical advice.

