## Supplementary Information for "Efficacy and Safety of Different Inhaler Types for Asthma and Chronic Obstructive Pulmonary Disease. A Systematic Review and Meta-Analysis"

#### Section A. Search Strategies

Search strategies were developed by an information specialist (SM). The PubMed search was developed first then translated for Embase and CENTRAL, using appropriate alternative thesaurus terms and search syntax. To test the reliability of the search, we used the studies included in the 2023 systematic review by Montoro et al.[1] The review by Montoro included 35 studies, 34 of which were included in PubMed. Our PubMed search strategy retrieved all 34 studies. Suggestions for names of common brands of inhalers were sought from clinical members of the review team and supplemented by searches of Google. No search limits were applied regarding date, language of publication or study design, but conference abstracts and trial registry records were excluded.

##### Table S1 PubMed search strategy

| Set | Concept | Search terms |
|---|---|---|
| #1 | Asthma or COPD | (Asthma[Mesh] OR Pulmonary Disease, Chronic Obstructive[Mesh] OR asthma[TIAB] OR "chronic obstructive pulmonary disease"[TIAB] OR COPD[TIAB] OR "respiratory disease*"[TIAB]) |
| #2 | Dry-powder inhalers | (Dry Powder Inhalers[Mesh] OR "dry powder inhaler*"[TIAB] OR "dry powdered inhaler*"[TIAB] OR DPI[TIAB] OR DPIs[TIAB] OR "soft mist inhaler*"[TIAB] OR "softmist inhaler*"[TIAB] OR SMI[TIAB] OR SMIs[TIAB] OR Turbuhaler*[TIAB] OR Breezhaler*[TIAB] OR Ellipta[TIAB] OR Diskus[TIAB] OR Genuair[TIAB] OR Accuhaler*[TIAB] OR Handihaler*[TIAB] OR Easyhaler*[TIAB] OR Rotahaler*[TIAB] OR Respimat[TIAB]) |
| #3 | Metered-dose inhalers | (Metered Dose Inhalers[Mesh] OR "metered dose inhaler*"[TIAB] OR "breath-actuated inhaler*"[TIAB] OR "aerosol inhaler*"[TIAB] OR MDI[TIAB] OR MDIs[TIAB] OR pMDI[TIAB] OR pMDIs[TIAB] OR BAI[TIAB] OR BAIs[TIAB]) |
| #4 | | #1 AND #2 AND #3 |
| #5 | Switching from inhaler type | #1 AND ((#2 AND switch*[TIAB]) OR (#3 AND switch*[TIAB])) |
| #6 | | #4 OR #5 |

**PubMed search string**

((Asthma[Mesh] OR Pulmonary Disease, Chronic Obstructive[Mesh] OR asthma[TIAB] OR "chronic obstructive pulmonary disease"[TIAB] OR COPD[TIAB] OR "respiratory disease*"[TIAB]) AND (Dry Powder Inhalers[Mesh] OR "dry powder inhaler*"[TIAB] OR "dry powdered inhaler*"[TIAB] OR DPI[TIAB] OR DPIs[TIAB] OR "soft mist inhaler*"[TIAB] OR "softmist inhaler*"[TIAB] OR SMI[TIAB] OR SMIs[TIAB] OR Turbuhaler*[TIAB] OR Breezhaler*[TIAB] OR Ellipta[TIAB] OR Diskus[TIAB] OR Genuair[TIAB] OR Accuhaler*[TIAB] OR Handihaler*[TIAB] OR Easyhaler*[TIAB] OR Rotahaler*[TIAB] OR Respimat[TIAB]) AND (Metered Dose Inhalers[Mesh] OR "metered dose inhaler*"[TIAB] OR "breath-actuated inhaler*"[TIAB] OR "aerosol inhaler*"[TIAB] OR MDI[TIAB] OR MDIs[TIAB] OR pMDI[TIAB] OR pMDIs[TIAB] OR BAI[TIAB] OR BAIs[TIAB])) OR ((Asthma[Mesh] OR Pulmonary Disease, Chronic Obstructive[Mesh] OR asthma[TIAB] OR "chronic obstructive pulmonary disease"[TIAB] OR COPD[TIAB] OR "respiratory disease*"[TIAB]) AND (((Dry Powder Inhalers[Mesh] OR "dry powder inhaler*"[TIAB] OR "dry powdered inhaler*"[TIAB] OR DPI[TIAB] OR DPIs[TIAB] OR Turbuhaler*[TIAB] OR Breezhaler*[TIAB] OR Ellipta[TIAB] OR Diskus[TIAB] OR Genuair[TIAB] OR Accuhaler*[TIAB] OR Handihaler*[TIAB] OR Easyhaler*[TIAB] OR Rotahaler*[TIAB] OR Respimat[TIAB]) AND switch*[TIAB]) OR ((Metered Dose Inhalers[Mesh] OR "metered dose inhaler*"[TIAB] OR "breath-actuated inhaler*"[TIAB] OR "aerosol inhaler*"[TIAB] OR MDI[TIAB] OR MDIs[TIAB] OR pMDI[TIAB] OR pMDIs[TIAB] OR BAI[TIAB] OR BAIs[TIAB]) AND switch*[TIAB])))

**Retrieved: 1084 records (1 February 2024)**

#### Table S2 Embase search strategy

**Embase (Ovid) 1947 to January 30, 2024**

| # | Search Statement | Results |
|---|---|---|
| 1 | exp Asthma/ or exp Chronic Obstructive Lung Disease/ or (asthma or chronic obstructive pulmonary disease or COPD or respiratory disease*).ti,ab,kf. | 578105 |
| 2 | Dry Powder Inhaler/ or (dry powder* inhaler* or soft mist inhaler* or softmist inhaler* or SMI or SMIs or DPI or DPIs or turbuhaler* or breezhaler* or ellipta or diskus or genuair or accuhaler* or handihaler* or easyhaler* or rotahaler or respimat).ti,ab,kf. | 30166 |
| 3 | exp Metered Dose Inhalers/ or (metered dose inhaler* or breath-actuated inhaler* or aerosol inhaler* or MDI or MDIs or pMDI or pMDIs or BAI or BAIs).ti,ab,kf. | 22687 |
| 4 | and/1-3 | 2568 |
| 5 | switch*.ti,ab,kf. | 280549 |
| 6 | 1 and ((2 and 5) or (3 and 5)) | 331 |
| 7 | 4 or 6 | 2768 |
| 8 | (note or letter or comment or editorial or review or conference abstract or conference paper or chapter).pt. | 12178369 |
| 9 | 7 not 8 | 1416 |

#### Table S3 Cochrane Central Register of Controlled Trials search strategy

**Cochrane Central Register of Controlled Trials (Cochrane Library) Issue 2 of 12, February 2024**

| # | Search | Hits |
|---|---|---|
| 1 | MeSH descriptor: [Asthma] explode all trees | 15076 |
| 2 | MeSH descriptor: [Pulmonary Disease, Chronic Obstructive] explode all trees | 8043 |
| 3 | (asthma or "chronic obstructive pulmonary disease" or COPD or "respiratory disease*"):ti,ab,kw (Word variations have been searched) | 58004 |
| 4 | #1 or #2 or #3 | 58778 |
| 5 | MeSH descriptor: [Dry Powder Inhalers] explode all trees | 225 |
| 6 | ((dry NEXT powder* NEXT inhaler*) or DPI or DPIs or (soft NEXT mist NEXT inhaler*) or (softmist NEXT inhaler*) or SMI or SMIs or turbuhaler* or breezhaler* or ellipta or diskus or genuair or accuhaler* or handihaler* or easyhaler* or rotahaler or respimat):ti,ab,kw (Word variations have been searched) | 5780 |
| 7 | #5 or #6 | 5780 |
| 8 | MeSH descriptor: [Metered Dose Inhalers] explode all trees | 538 |
| 9 | ((metered NEXT dose NEXT inhaler*) or (breath NEXT actuated NEXT inhaler*) or (aerosol NEXT inhaler*) or MDI or MDIs or pMDI or pMDIs or BAI or BAIs):ti,ab,kw (Word variations have been searched) | 5855 |
| 10 | #8 or #9 | 5864 |
| 11 | #4 and # 7 and #10 | 976 |
| 12 | (switch*):ti,ab,kw (Word variations have been searched) | 19759 |
| 13 | ((#7 and #12) or (#10 and #12)) | 122 |
| 14 | #11 or #13 | 1074 |
| 15 | ("conference proceeding" or "trial registry record"):pt | 725633 |
| 16 | #14 not #15 in Trials | 606 |

#### Section B. Additional methods

##### Figure S1 – Risk of Bias assessments for each study

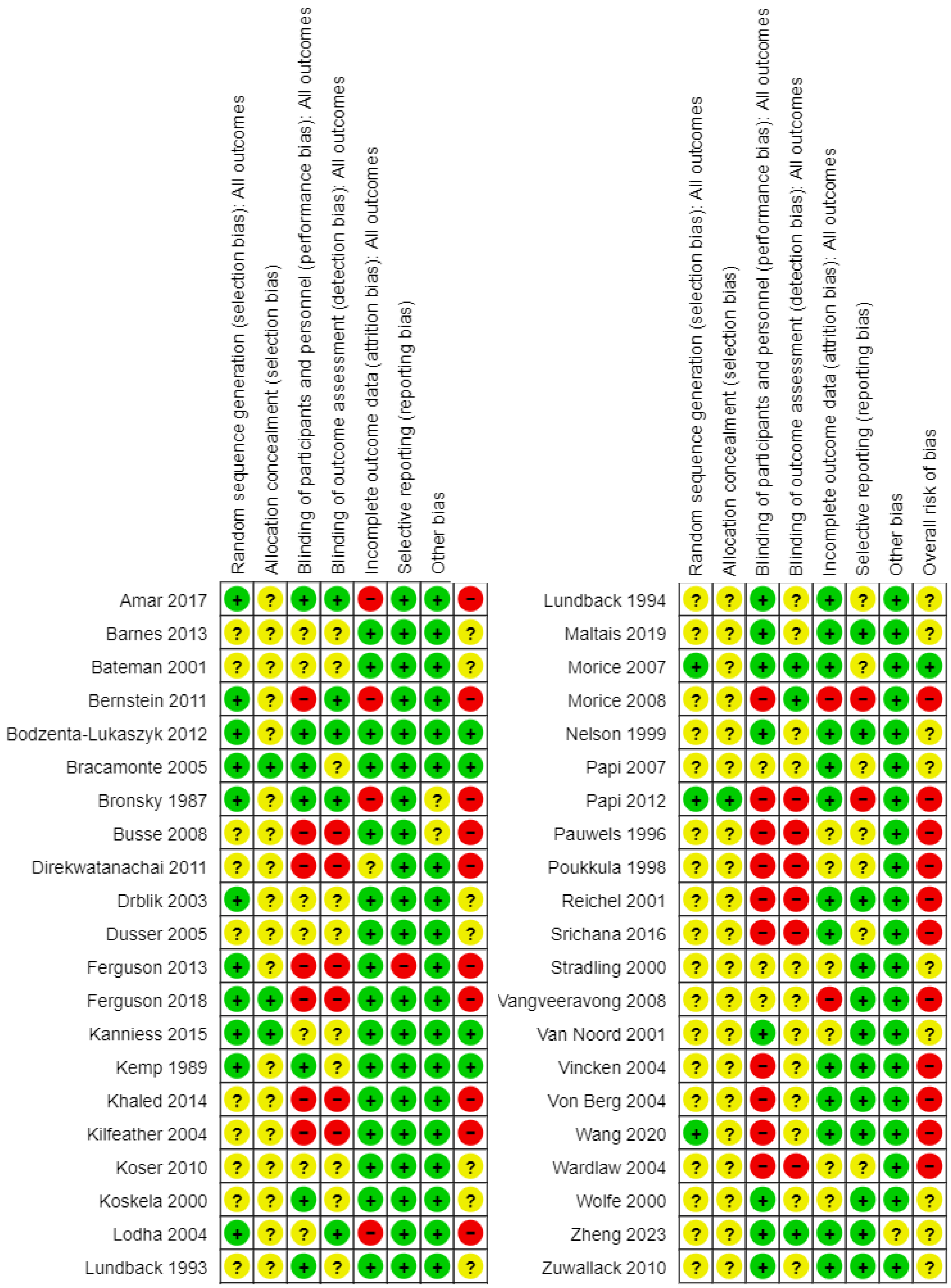

#### Table S4 – Summary of Minimal Clinically Important Differences Used for GRADE Assessment

*Asthma – Maintenance*

| Parameter | MCID used for GRADE assessment | Source | Comments |
|---|---|---|---|
| $FEV_1$ | 20% predicted value | Bonini et al, 2020 [2]<br>Pellegrino et al, 2005 [3] | Literatures suggests ≥15% for long-term trials (years) and ≥20% for short-term trials (weeks). Given the median follow-up time of 12 weeks for included studies, we have used this 20% value.<br>Of note these are consensus values without strong original evidence to support them. |
| Peak Expiratory Flow Rate (PEFR) | 18.8L/min | Santanello et al, 1999 [4] | |
| Asthma Control Questionnaire (ACQ) | 0.5 points | Bonini et al, 2020 [2] | Consensus value |
| Reliever use | 0.81 puffs/day | Santanello et al, 1999 [4] | |
| Exacerbations | 5% (absolute risk difference) | Bonini et al, 2020 [2] | Consensus value. A reduction in *annual* exacerbation rate or in the risk of having a severe asthma-related event ranging from 20–40% for a given asthma treatment regimen and/or intervention is considered clinically relevant in RCTs.<br>Given median follow up of 12 weeks for included studies, we have taken the conservative estimate of 5% (will make it less likely for no difference to be shown) |
| Adverse events | 5% (absolute risk difference) | Expert opinion | No published MCID estimates/values – determined following discussion with respiratory experts on authorship team, to permit GRADE assessment |

*Asthma – Acute*

| Parameter | MCID used for GRADE assessment | Source | Comments |
| --- | --- | --- | --- |
| $FEV_1$ | 20% predicted value | Extrapolated from asthma maintenance (Bonini et al, 2020) [2] | See comments above under Asthma maintenance. |
| Peak Expiratory Flow Rate (PEFR) | 12% predicted value | Karras et al, 2000 [5] | The two included acute asthma studies reporting PEFR did so using *absolute* PEFR values (not % predicted values). Both were paediatric studies.<br>We have conservatively estimated that the end of treatment PEFRs (approx. 208L/min in both arms) were those individuals' baseline values (a conservative assumption, as values immediately post-exacerbation may be lower than baseline). Using the Karras et al percent predicted MCID; 12% of 208L/min is 24.96, rounded up to 25L/min.<br>NB - this estimate would not apply to other groups (e.g. adults) |
| | 25L/min | Expert opinion (and only relevant to the studies included in this review) | |
| Modified Wood Clinical Asthma Score | 1 point | Expert opinion<br><br>(Influenced by Duarte-Dorado et al, 2013) [6] | No published MCID estimates/values. Duarte-Dorado showed 2-point difference between patients admitted to PICU versus regular ward. Inter-rater agreement showed that difference of 0.5 between assessors was common.<br>Hence in discussion with respiratory experts on authorship team have determined an MCID estimate of 1 point. |
| Adverse events | 5% (absolute risk difference) | Expert opinion | See comments above under Asthma maintenance. |

*COPD*

| **Parameter** | **MCID used for GRADE assessment** | **Source** | **Comments** |
|---|---|---|---|
| $FEV_1$ | 100 mL | Jones et al, 2014 [7]<br>Cazzola et al, 2008 [8] | |
| PEFR | 18.8L/min | Expert opinion | No published MCID estimates/values. Have extrapolated from asthma maintenance to permit GRADE assessment. |
| Exacerbations | 20% (absolute change) | Expert opinion (influenced by Jones et al, 2014) [7] | Jones et al states no validated MCID. Have used widely suggested value of 20% to permit GRADE assessment. |
| CAT score (COPD assessment test) | 2 points | Kon et al, 2014 [9] | |
| Reliever use | 0.81puffs/day | Expert opinion | No published MCID estimates/values. Have extrapolated from asthma maintenance to permit GRADE assessment. |
| Adverse events | 5% (absolute risk difference) | Expert opinion | See comments above under Asthma maintenance. |
| Mortality | 1% (absolute risk difference) | Expert opinion | No published MCID estimates/values – decided after discussion with respiratory experts on authorship team, to permit GRADE assessment. |

### Section C. Summary of findings

#### Table S5 – Summary of findings for asthma maintenance

| Outcome and follow-up | Patients (studies), N | Relative effect (95% CI) | Absolute effects (95% CI) | | | Certainty | What happens |
|---|---|---|---|---|---|---|---|
| | | | pMDIs | non-pMDIs | Difference | | |
| $FEV_1$, % predicted Follow-up: range 4 weeks to 52 weeks | 9181 (27 RCTs) | - | 83.18 % | **83.75 %** | **0.57** (-0.14 to 1.42) | ⊕⊕⊕◯ Moderate[a] | Non-pMDIs probably result in little to no difference in $FEV_1$. |
| PEFR, L/min Follow-up: range 4 weeks to 30 weeks | 8083 (24 RCTs) | - | 393.94 L/min | **394.93 L/min** | **0.99** (-1.11 to 3.09) | ⊕⊕⊕◯ Moderate[a] | Non-pMDIs probably result in little to no difference in PEFR. |
| Symptom control, ACQ-7 Follow-up: range 8 weeks to 30 weeks | 3836 (8 RCTs) | - | 0 points (change from baseline) | **-0.024 points (change from baseline)** | **-0.024** (-0.05 to 0.01) | ⊕⊕⊕◯ Moderate[a] | Non-pMDIs may result in little to no difference in symptom control. |
| Quality of life, AQLQ ≥0.5 improved from baseline Follow-up: range 12 weeks to 30 weeks | 871 (2 RCTs) | **RR = 1.02** (0.91 to 1.14) | 575 per 1,000 | **586 per 1,000** (523 to 655) | **11 more per 1,000** (from 52 fewer to 80 more) | ⊕◯◯◯ Very low[a,b] | The evidence is very uncertain about the effect of non-pMDIs on quality of life. |
| Reliever use, puffs/day Follow-up: range 4 weeks to 30 weeks | 4308 (13 RCTs) | - | 1.27 puffs/day | **1.32 puffs/day** | **0.05** (-0.16 to 0.23) | ⊕⊕⊕◯ Moderate[a] | Non-pMDIs probably result in little to no difference in reliever use. |
| Exacerbations, No. with at least one Follow-up: range 4 weeks to 52 weeks | 6755 (17 RCTs) | **RR = 0.88** (0.73 to 1.07) | 68 per 1,000 | **60 per 1,000** (50 to 73) | **8 fewer per 1,000** (from 18 fewer to 5 more) | ⊕⊕⊕◯ Moderate[a] | Non-pMDIs probably result in little to no difference in exacerbations. |
| Mortality | (0 studies) | No deaths were reported. | | | | - | |
| Adverse events, No. with at least one Follow-up: range 4 weeks to 52 weeks | 7946 (21 RCTs) | **RR = 0.97** (0.93 to 1.02) | 428 per 1,000 | **415 per 1,000** (398 to 437) | **13 fewer per 1,000** (from 30 fewer to 9 more) | ⊕⊕⊕◯ Moderate[a] | Non-pMDIs probably result in little to no difference in adverse events. |

**CI:** confidence interval; **MD:** mean difference; **RR:** risk ratio
a. Most included studies at high or unclear risk of bias.
b. Confidence intervals include the possibility of both a meaningful benefit and a meaningful harm

### Table S6 – Summary of findings for COPD

| Outcome and follow-up | Patients (studies), N | Relative effect (95% CI) | Absolute effects (95% CI) | | | Certainty | What happens |
|---|---|---|---|---|---|---|---|
| | | | pMDIs | non-pMDIs | Difference | | |
| $FEV_1$, L Follow-up: range 12 weeks to 48 weeks | 3946 (7 RCTs) | - | 0.23 L (change from baseline) | **0.24 L (change from baseline)** | **0.01** (-0.01 to 0.02) | ⊕⊕⊕◯ Moderate[a] | Non-pMDIs probably result in little to no difference in $FEV_1$. |
| PEFR, L/min Follow-up: 12 weeks | 644 (2 RCTs) | - | 232.35 L/min | **229.79 L/min** | **-2.56** (-9.17 to 4.05) | ⊕⊕⊕◯ Moderate[a] | Non-pMDIs probably result in little to no difference in PEFR. |
| Symptom control, CAT score Follow-up: 24 weeks | 1006 (1 RCT) | - | -3.56 points (change from baseline) | **-4.15 points (change from baseline)** | **-0.59** (-1.19 to 0.01) | ⊕⊕⊕◯ Moderate[a] | Non-pMDIs probably result in little to no difference in symptom control. |
| Quality of life | 871 (2 RCTs) | Two studies reported this outcome but the results could not be pooled. | | | | ⊕◯◯◯ Very low[a,b,c] | The evidence is very uncertain about the effect of non-pMDIs on quality of life. |
| Reliever use, puffs/day Follow-up: range 12 weeks to 24 weeks | 1265 (3 RCTs) | - | 2.3 puffs/day | **2.09 puffs/day** | **-0.21** (-0.51 to 0.1) | ⊕⊕⊕◯ Moderate[a] | Non-pMDIs probably result in little to no difference in reliever use. |
| Exacerbations, No. with at least one Follow-up: range 12 weeks to 48 weeks | 4101 (7 RCTs) | **RR = 1.08** (0.94 to 1.24) | 147 per 1,000 | **159 per 1,000** (139 to 183) | **12 more per 1,000** (from 9 fewer to 35 more) | ⊕⊕⊕◯ Moderate[a] | Non-pMDIs probably result in little to no difference in exacerbations. |
| Mortality Follow-up: range 12 weeks to 48 weeks | 3657 (6 RCTs) | **RR = 1.40** (0.59 to 3.32) | 5 per 1,000 | **8 per 1,000** (3 to 18) | **2 more per 1,000** (from 2 fewer to 12 more) | ⊕⊕◯◯ Low[a,c] | Non-pMDIs may result in little to no difference in mortality. |
| Adverse events, No. with at least one Follow-up: range 12 weeks to 48 weeks | 4106 (7 RCTs) | **RR = 1.02** (0.94 to 1.11) | 496 per 1,000 | **506 per 1,000** (466 to 551) | **10 more per 1,000** (from 30 fewer to 55 more) | ⊕⊕◯◯ Low[a,c] | Non-pMDIs may result in little to no difference in adverse events. |

**CI:** confidence interval; **MD:** mean difference; **RR:** risk ratio
a. Included studies at high or unclear risk of bias.
b. Studies were not pooled, therefore precision and heterogeneity could not be estimated. The studies differed in their direction of effect.
c. Confidence intervals include the possibility of both little or no effect and a meaningful harm.

### Table S7 – Summary of findings for acute asthma exacerbations

| Outcome and follow-up | Patients (studies), N | Relative effect (95% CI) | Absolute effects (95% CI) | | | Certainty | What happens |
|---|---|---|---|---|---|---|---|
| | | | **pMDIs** | **non-pMDIs** | **Difference** | | |
| $FEV_1$, % predicted Follow-up: 60 minutes | 103 (1 RCT) | - | 66.7 % | **68.7 %** | **2** (-2.9 to 6.9) | ⊕⊕◯◯ Low[a,b] | Non-pMDIs may result in little to no difference in $FEV_1$. |
| PEFR, L/min Follow-up: 30 minutes | 259 (2 RCTs) | - | 207.5 L/min | **208.88 L/min** | **1.38** (-17.97 to 20.73) | ⊕⊕◯◯ Low[a,b] | Non-pMDIs may result in little to no difference in PEFR. |
| Symptom control, Modified Wood Clinical Asthma Score Follow-up: 60 minutes | 36 (1 RCT) | - | 1.6 points | **1.5 points** | **-0.1** (-0.72 to 0.52) | ⊕⊕◯◯ Low[a,b] | Non-pMDIs may result in little to no difference in symptom control. |
| Symptom control | (0 studies) | No studies reported symptom control. | | | | | |
| Quality of life | (0 studies) | No studies reported quality of life. | | | | | |
| Reliever use | (0 studies) | No studies reported reliever use. | | | | | |
| Mortality | (0 studies) | No deaths were reported | | | | | |
| Adverse events, No. with at least one | 36 (1 RCT) | **RR = 0.33** (0.04 to 2.91) | 56 per 1,000 | **18 per 1,000** (2 to 162) | **37 fewer per 1,000** (from 53 fewer to 106 more) | ⊕◯◯◯ Very low[a,b,c] | The evidence is very uncertain about the effect of non-pMDIs on adverse events. |

**CI:** confidence interval; **MD:** mean difference; **RR:** risk ratio
a. Included studies at high or unclear risk of bias.
b. Insufficient information for precision, based on too few participants.
c. Confidence interval includes the possibility of both important benefit and important harm.

### Section D. Additional Results

#### $FEV_1$

*Table S8 Asthma maintenance: Additional $FEV_1$ results not included in the meta-analysis*

| Study ID | Timepoint | Outcome measure | Effect estimate | Direction |
|---|---|---|---|---|
| Kemp 1989 | Visit 7 (<12 weeks) | Change in $FEV_1$ (% predicted) between Hour 0 and Hour 8 | Non-pMDI mean: 7.89<br>pMDI mean: 6.85<br>No measure of variance reported. | Better with non-pMDI. |

*Figure S2. $FEV_1$ in Acute Asthma Exacerbations (% predicted)*

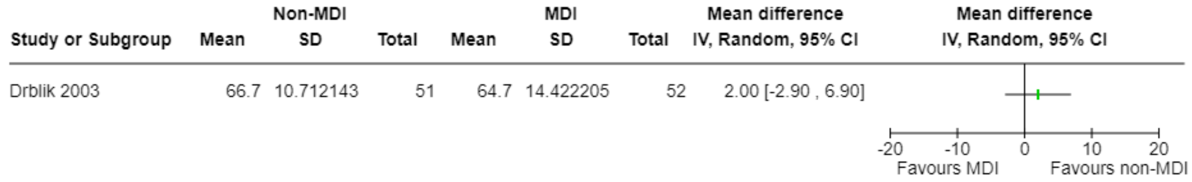

#### PEFR

*Table S9 Asthma maintenance: Additional PEFR results not included in meta-analysis*

| Study ID | Timepoint | Outcome measure | Effect estimate | Direction of effect |
|---|---|---|---|---|
| Pauwels 1996 | 52 weeks | Risk of at least one 'event'<br>(2 consecutive days PEF <80% baseline) | RR 0.80<br>(95% CI 0.68 to 0.95) | Better with non-pMDI. |

*Figure S3. PEFR (L/min) in (A) Acute Asthma Exacerbations and (B) COPD*

**A**

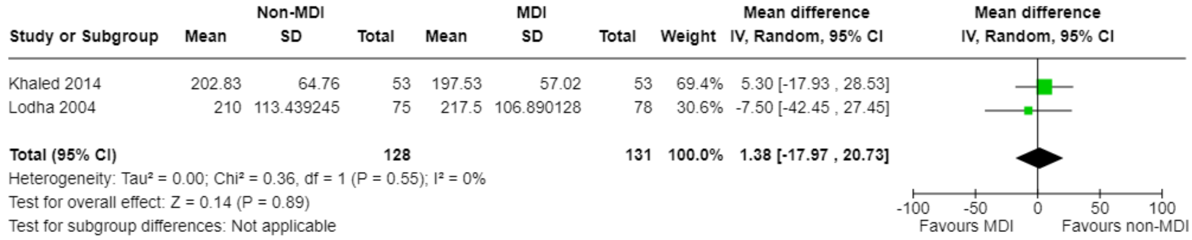

**B**

| Study or Subgroup | MD | SE | Weight | Mean difference IV, Random, 95% CI |
|---|---|---|---|---|
| Ferguson 2013 | -5 | 10.797666 | 9.8% | -5.00 [-26.16 , 16.16] |
| Koser 2010 | -2.3 | 3.55 | 90.2% | -2.30 [-9.26 , 4.66] |
| **Total (95% CI)** | | | **100.0%** | **-2.56 [-9.17 , 4.05]** |

Heterogeneity: Tau² = 0.00; Chi² = 0.06, df = 1 (P = 0.81); I² = 0%
Test for overall effect: Z = 0.76 (P = 0.45)
Test for subgroup differences: Not applicable

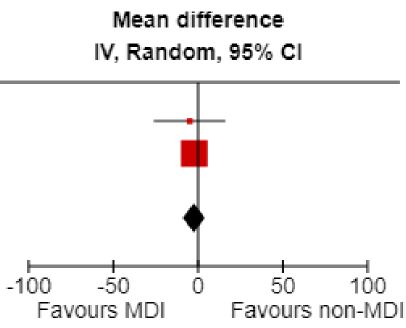

#### Disease exacerbations

*Figure S4. Disease exacerbations (risk of >1) in (A) Asthma Maintenance and (B) COPD*

**A**

| Study or Subgroup | Non-MDI Events | Non-MDI Total | MDI Events | MDI Total | Weight | Risk ratio IV, Random, 95% CI |
|---|---|---|---|---|---|---|
| Amar 2017 | 4 | 122 | 2 | 114 | 1.3% | 1.87 [0.35 , 10.01] |
| Barnes 2013 | 4 | 209 | 3 | 207 | 1.7% | 1.32 [0.30 , 5.83] |
| Bernstein 2011 | 21 | 351 | 20 | 371 | 9.5% | 1.11 [0.61 , 2.01] |
| Bodzenta-Lukaszyk 2012 | 2 | 120 | 1 | 126 | 0.7% | 2.10 [0.19 , 22.86] |
| Bracamonte 2005 | 6 | 160 | 6 | 168 | 2.9% | 1.05 [0.35 , 3.19] |
| Busse 2008 | 37 | 404 | 37 | 422 | 16.3% | 1.04 [0.68 , 1.61] |
| Dusser 2005 | 1 | 220 | 1 | 225 | 0.5% | 1.02 [0.06 , 16.25] |
| Kanniess 2015 | 4 | 251 | 3 | 251 | 1.7% | 1.33 [0.30 , 5.90] |
| Kemp 1989 | 13 | 100 | 26 | 104 | 9.2% | 0.52 [0.28 , 0.95] |
| Koskela 2000 | 6 | 68 | 2 | 76 | 1.5% | 3.35 [0.70 , 16.06] |
| Morice 2008 | 4 | 227 | 19 | 446 | 3.2% | 0.41 [0.14 , 1.20] |
| Papi 2007 | 12 | 109 | 17 | 107 | 7.3% | 0.69 [0.35 , 1.38] |
| Papi 2012 | 31 | 216 | 37 | 206 | 16.2% | 0.80 [0.52 , 1.24] |
| Stradling 2000 | 8 | 98 | 18 | 106 | 5.7% | 0.48 [0.22 , 1.06] |
| Vincken 2004 | 26 | 161 | 19 | 159 | 10.9% | 1.35 [0.78 , 2.34] |
| Von Berg 2004 | 10 | 180 | 16 | 177 | 6.0% | 0.61 [0.29 , 1.32] |
| Zheng 2023 | 12 | 252 | 11 | 242 | 5.5% | 1.05 [0.47 , 2.33] |
| **Total (95% CI)** | | **3248** | | **3507** | **100.0%** | **0.88 [0.73 , 1.07]** |
| Total events: | 201 | | 238 | | | |

Heterogeneity: Tau² = 0.01; Chi² = 17.07, df = 16 (P = 0.38); I² = 6%
Test for overall effect: Z = 1.29 (P = 0.20)
Test for subgroup differences: Not applicable

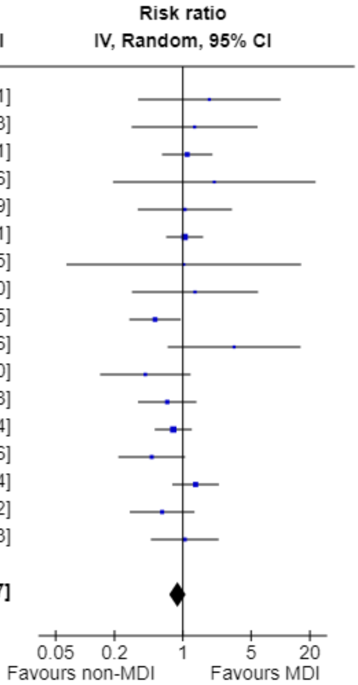

**B**

| Study or Subgroup | Non-MDI Events | Non-MDI Total | MDI Events | MDI Total | Weight | Risk ratio IV, Random, 95% CI |
|---|---|---|---|---|---|---|
| Ferguson 2013 | 39 | 157 | 41 | 154 | 13.2% | 0.93 [0.64 , 1.36] |
| Ferguson 2018 | 32 | 219 | 111 | 655 | 14.4% | 0.86 [0.60 , 1.24] |
| Kilfeather 2004 | 60 | 224 | 46 | 220 | 16.7% | 1.28 [0.92 , 1.79] |
| Koser 2010 | 16 | 126 | 10 | 121 | 3.4% | 1.54 [0.73 , 3.25] |
| Maltais 2019 | 97 | 552 | 92 | 552 | 28.0% | 1.05 [0.81 , 1.37] |
| Wang 2020 | 18 | 72 | 14 | 72 | 5.0% | 1.29 [0.69 , 2.38] |
| Zuwallack 2010 | 72 | 486 | 64 | 491 | 19.3% | 1.14 [0.83 , 1.55] |
| **Total (95% CI)** | | **1836** | | **2265** | **100.0%** | **1.08 [0.94 , 1.24]** |
| Total events: | 334 | | 378 | | | |

Heterogeneity: Tau² = 0.00; Chi² = 4.34, df = 6 (P = 0.63); I² = 0%
Test for overall effect: Z = 1.10 (P = 0.27)
Test for subgroup differences: Not applicable

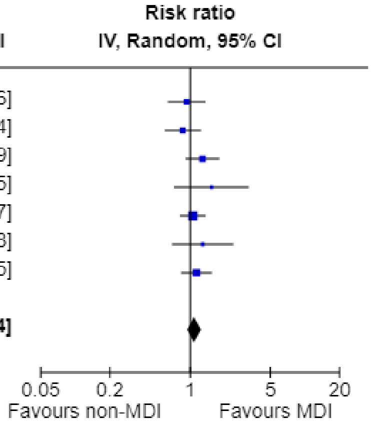

*Table S10 Asthma maintenance: Additional disease exacerbation results not included in meta-analysis*

| Study ID | Timepoint | Outcome measure | Effect estimate | Direction of effect |
|---|---|---|---|---|
| Bateman 2001 | 12 weeks | Asthma resulting in emergency treatment, hospitalization, or treatment with additional (excluded) asthma medication (eg, systemic glucocorticoids) | Both groups: 2-3% | Unknown |

#### Adverse events

*Figure S5. Adverse Events (risk of >1) in Acute Asthma Exacerbations*

| Study or Subgroup | Non-MDI Events | Non-MDI Total | MDI Events | MDI Total | Risk ratio IV, Random, 95% CI |
|---|---|---|---|---|---|
| Vangveeravong 2008 | 1 | 18 | 3 | 18 | 0.33 [0.04 , 2.91] |

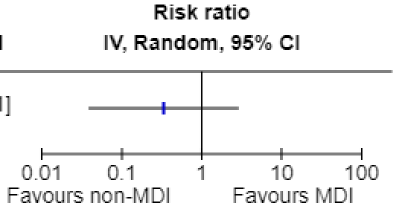

*Figure S6. Serious Adverse Events (risk of >1) in (A) Asthma maintenance and (B) COPD*

**A**

| Study or Subgroup | Non-MDI Events | Non-MDI Total | MDI Events | MDI Total | Weight | Risk ratio IV, Random, 95% CI |
|---|---|---|---|---|---|---|
| Amar 2017 | 2 | 122 | 1 | 114 | 1.8% | 1.87 [0.17 , 20.33] |
| Barnes 2013 | 2 | 209 | 2 | 207 | 2.6% | 0.99 [0.14 , 6.97] |
| Bateman 2001 | 3 | 167 | 3 | 165 | 4.0% | 0.99 [0.20 , 4.82] |
| Bernstein 2011 | 5 | 351 | 5 | 371 | 6.6% | 1.06 [0.31 , 3.62] |
| Bodzenta-Lukaszyk 2012 | 2 | 139 | 1 | 140 | 1.8% | 2.01 [0.18 , 21.96] |
| Bracamonte 2005 | 2 | 160 | 2 | 168 | 2.7% | 1.05 [0.15 , 7.37] |
| Bronsky 1987 | 3 | 115 | 5 | 116 | 5.1% | 0.61 [0.15 , 2.47] |
| Busse 2008 | 9 | 406 | 10 | 427 | 12.7% | 0.95 [0.39 , 2.31] |
| Dusser 2005 | 3 | 221 | 6 | 227 | 5.3% | 0.51 [0.13 , 2.03] |
| Koskela 2000 | 1 | 68 | 0 | 76 | 1.0% | 3.35 [0.14 , 80.83] |
| Lundback 1994 | 7 | 150 | 5 | 146 | 8.0% | 1.36 [0.44 , 4.20] |
| Morice 2007 | 0 | 229 | 2 | 233 | 1.1% | 0.20 [0.01 , 4.22] |
| Morice 2008 | 15 | 227 | 32 | 446 | 28.7% | 0.92 [0.51 , 1.66] |
| Nelson 1999 | 1 | 97 | 1 | 92 | 1.3% | 0.95 [0.06 , 14.94] |
| Papi 2007 | 0 | 109 | 0 | 109 | | Not estimable |
| Papi 2012 | 3 | 216 | 1 | 206 | 2.0% | 2.86 [0.30 , 27.28] |
| Reichel 2001 | 0 | 93 | 0 | 92 | | Not estimable |
| Van Noord 2001 | 3 | 161 | 5 | 176 | 5.0% | 0.66 [0.16 , 2.70] |
| Vincken 2004 | 5 | 161 | 2 | 159 | 3.8% | 2.47 [0.49 , 12.54] |
| Von Berg 2004 | 0 | 180 | 3 | 177 | 1.2% | 0.14 [0.01 , 2.70] |
| Wardlaw 2004 | 0 | 82 | 0 | 85 | | Not estimable |
| Zheng 2023 | 4 | 252 | 4 | 242 | 5.3% | 0.96 [0.24 , 3.80] |
| **Total (95% CI)** | | **3915** | | **4174** | **100.0%** | **0.96 [0.70 , 1.32]** |
| Total events: | 70 | | 90 | | | |

Heterogeneity: Tau² = 0.00; Chi² = 8.00, df = 18 (P = 0.98); I² = 0%
Test for overall effect: Z = 0.22 (P = 0.82)
Test for subgroup differences: Not applicable

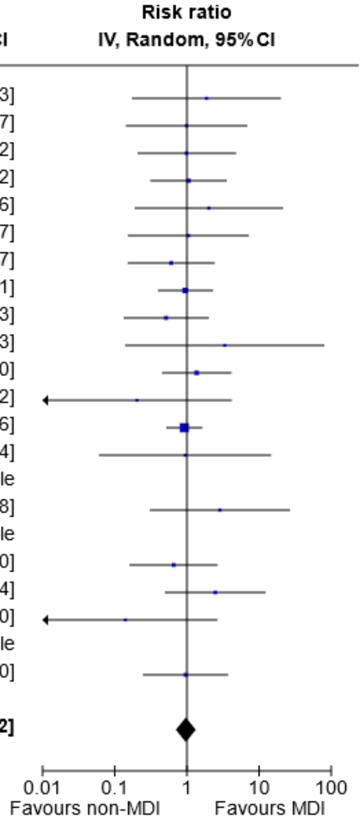

**B**

| Study or Subgroup | Non-MDI Events | Non-MDI Total | MDI Events | MDI Total | Weight | Risk ratio IV, Random, 95% CI |
|---|---|---|---|---|---|---|
| Ferguson 2013 | 23 | 157 | 20 | 154 | 22.2% | 1.13 [0.65 , 1.97] |
| Ferguson 2018 | 20 | 219 | 42 | 655 | 23.3% | 1.42 [0.86 , 2.37] |
| Maltais 2019 | 40 | 552 | 32 | 552 | 24.8% | 1.25 [0.80 , 1.96] |
| Wang 2020 | 11 | 72 | 2 | 72 | 7.9% | 5.50 [1.26 , 23.94] |
| Zuwallack 2010 | 17 | 486 | 33 | 491 | 21.8% | 0.52 [0.29 , 0.92] |
| **Total (95% CI)** | | **1486** | | **1924** | **100.0%** | **1.17 [0.73 , 1.89]** |
| Total events: | 111 | | 129 | | | |

Heterogeneity: Tau² = 0.19; Chi² = 12.48, df = 4 (P = 0.01); I² = 68%
Test for overall effect: Z = 0.64 (P = 0.52)
Test for subgroup differences: Not applicable

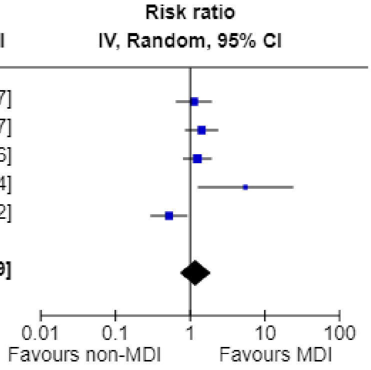

*Figure S7. Treatment-related adverse events (risk of >1) in (A) Asthma maintenance and (B) COPD*

**A**

| Study or Subgroup | log[RR] | SE | Weight | Risk ratio IV, Random, 95% CI |
|---|---|---|---|---|
| Amar 2017 | -0.067823 | 1.408201 | 0.6% | 0.93 [0.06 , 14.76] |
| Bateman 2001 | 0.313374 | 0.34703 | 10.7% | 1.37 [0.69 , 2.70] |
| Bernstein 2011 | 0.055416 | 0.251835 | 20.3% | 1.06 [0.65 , 1.73] |
| Bodzenta-Lukaszyk 2012 | -0.244146 | 0.489521 | 5.4% | 0.78 [0.30 , 2.04] |
| Bracamonte 2005 | 0.009346 | 0.700467 | 2.6% | 1.01 [0.26 , 3.98] |
| Dusser 2005 | -0.301717 | 0.294322 | 14.9% | 0.74 [0.42 , 1.32] |
| Kanniess 2015 | -0.405465 | 0.908496 | 1.6% | 0.67 [0.11 , 3.96] |
| Morice 2007 | 0.86762 | 0.978779 | 1.3% | 2.38 [0.35 , 16.22] |
| Morice 2008 | -0.24516 | 0.63251 | 3.2% | 0.78 [0.23 , 2.70] |
| Poukkula 1998 | 0.49443 | 0.92467 | 1.5% | 1.64 [0.27 , 10.04] |
| Reichel 2001 | -0.927102 | 0.823637 | 1.9% | 0.40 [0.08 , 1.99] |
| Van Noord 2001 | -0.156043 | 0.295196 | 14.8% | 0.86 [0.48 , 1.53] |
| Wardlaw 2004 | 0.487917 | 0.458046 | 6.1% | 1.63 [0.66 , 4.00] |
| Wolfe 2000 | 0.206713 | 0.435816 | 6.8% | 1.23 [0.52 , 2.89] |
| Zheng 2023 | 0.200671 | 0.39273 | 8.3% | 1.22 [0.57 , 2.64] |
| **Total (95% CI)** | | | **100.0%** | **1.01 [0.81 , 1.26]** |

Heterogeneity: Tau² = 0.00; Chi² = 6.74, df = 14 (P = 0.94); I² = 0%
Test for overall effect: Z = 0.10 (P = 0.92)
Test for subgroup differences: Not applicable

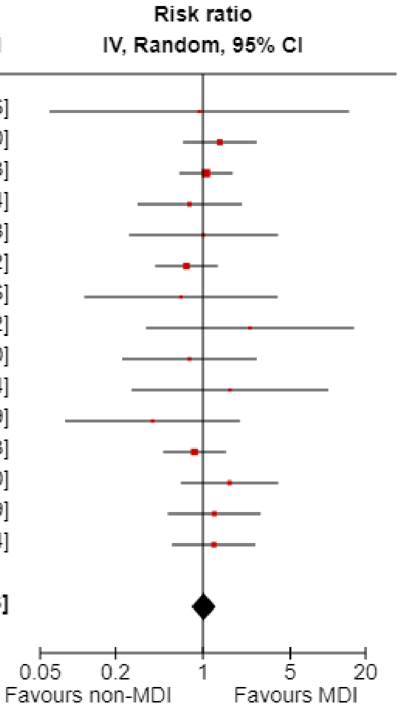

**B**

| Study or Subgroup | Non-MDI Events | Non-MDI Total | MDI Events | MDI Total | Weight | Risk ratio IV, Random, 95% CI |
|---|---|---|---|---|---|---|
| Ferguson 2018 | 13 | 219 | 57 | 655 | 43.5% | 0.68 [0.38 , 1.22] |
| Maltais 2019 | 3 | 552 | 7 | 552 | 13.8% | 0.43 [0.11 , 1.65] |
| Wang 2020 | 19 | 72 | 15 | 72 | 42.7% | 1.27 [0.70 , 2.29] |
| **Total (95% CI)** | | **843** | | **1279** | **100.0%** | **0.83 [0.48 , 1.44]** |
| Total events: | 35 | | 79 | | | |

Heterogeneity: Tau² = 0.09; Chi² = 3.27, df = 2 (P = 0.19); I² = 39%
Test for overall effect: Z = 0.65 (P = 0.51)
Test for subgroup differences: Not applicable

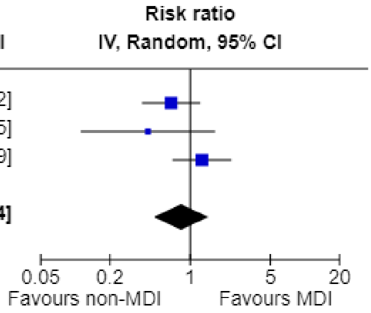

*Figure S8. Treatment-related serious adverse events (risk of >1) in (A) Asthma maintenance and (B) COPD*

**A**

| Study or Subgroup | Non-MDI Events | Non-MDI Total | MDI Events | MDI Total | Weight | Risk ratio IV, Random, 95% CI |
|---|---|---|---|---|---|---|
| Bateman 2001 | 1 | 167 | 1 | 165 | 30.8% | 0.99 [0.06 , 15.66] |
| Bernstein 2011 | 0 | 351 | 1 | 371 | 23.0% | 0.35 [0.01 , 8.62] |
| Bodzenta-Lukaszyk 2012 | 0 | 139 | 0 | 140 | | Not estimable |
| Bracamonte 2005 | 0 | 160 | 0 | 168 | | Not estimable |
| Busse 2008 | 0 | 406 | 0 | 427 | | Not estimable |
| Dusser 2005 | 0 | 221 | 1 | 227 | 23.1% | 0.34 [0.01 , 8.36] |
| Nelson 1999 | 0 | 97 | 0 | 92 | | Not estimable |
| Van Noord 2001 | 1 | 161 | 0 | 176 | 23.1% | 3.28 [0.13 , 79.89] |
| Vincken 2004 | 0 | 161 | 0 | 159 | | Not estimable |
| Von Berg 2004 | 0 | 180 | 0 | 177 | | Not estimable |
| Zheng 2023 | 0 | 252 | 0 | 242 | | Not estimable |
| **Total (95% CI)** | | **2295** | | **2344** | **100.0%** | **0.80 [0.17 , 3.73]** |
| Total events: | 2 | | 3 | | | |

Heterogeneity: Tau² = 0.00; Chi² = 1.30, df = 3 (P = 0.73); I² = 0%
Test for overall effect: Z = 0.28 (P = 0.78)
Test for subgroup differences: Not applicable

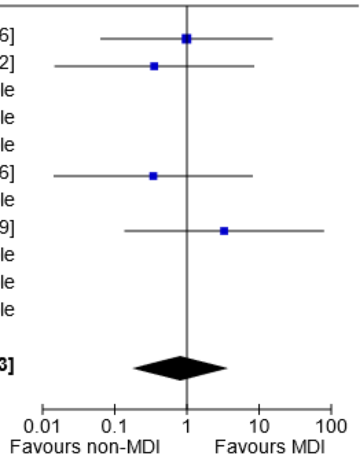

**B**

| Study or Subgroup | Non-MDI Events | Non-MDI Total | MDI Events | MDI Total | Weight | Risk ratio IV, Random, 95% CI |
|---|---|---|---|---|---|---|
| Ferguson 2018 | 0 | 219 | 2 | 655 | 33.8% | 0.60 [0.03 , 12.37] |
| Wang 2020 | 4 | 72 | 1 | 72 | 66.2% | 4.00 [0.46 , 34.92] |
| **Total (95% CI)** | | **291** | | **727** | **100.0%** | **2.10 [0.36 , 12.27]** |
| Total events: | 4 | | 3 | | | |

Heterogeneity: Tau² = 0.00; Chi² = 1.00, df = 1 (P = 0.32); I² = 0%
Test for overall effect: Z = 0.82 (P = 0.41)
Test for subgroup differences: Not applicable

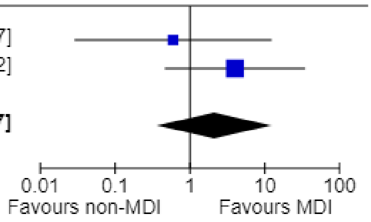

#### Quality of life

*Figure S9. Quality of life (AQLQ ≥0.5 improved from baseline) in Asthma Maintenance*

| Study or Subgroup | Non-MDI Events | Non-MDI Total | MDI Events | MDI Total | Weight | Risk ratio IV, Random, 95% CI |
|---|---|---|---|---|---|---|
| Busse 2008 | 128 | 207 | 134 | 212 | 57.0% | 0.98 [0.84 , 1.13] |
| Morice 2007 | 128 | 229 | 116 | 223 | 43.0% | 1.07 [0.91 , 1.27] |
| **Total (95% CI)** | | **436** | | **435** | **100.0%** | **1.02 [0.91 , 1.14]** |
| Total events: | 256 | | 250 | | | |

Heterogeneity: Tau² = 0.00; Chi² = 0.66, df = 1 (P = 0.42); I² = 0%
Test for overall effect: Z = 0.32 (P = 0.75)
Test for subgroup differences: Not applicable

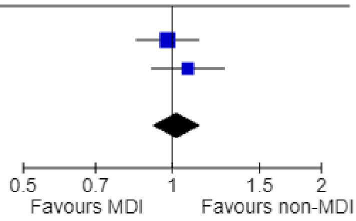

*Table S11 Asthma maintenance and COPD: Additional quality of life results not included in meta-analysis*

| **Study ID** | **Timepoint** | **Outcome measure** | **Effect estimate** | **Direction of effect** |
|---|---|---|---|---|
| **Asthma maintenance** | | | | |
| Amar 2017 | 12 weeks | PAQLQ[S]) | Mean in pMDI group 0.35 (range 0.23 to 0.48). NO data reported for non-pMDI group. | Unknown |
| Bernstein 2011 | 12 weeks | AQLQ[S] | MD 0 (no measure of variance) (difference in least square means) | No difference |
| Bodzenta-Lukaszyk 2012 | 12 weeks | Change in AQLQ[S] | MD 0 (95% CI -0.2 to 0.1) | No difference |
| Koskela 2000 | 8 weeks | SGRQ (Parts I–II) | Difference in medians: -0.8 (no measure of variance) | Better with non-pMDI |
| **COPD** | | | | |
| Ferguson 2018 | 24 weeks | SGRQ (proportion achieving MID ≥4 units) | RD -2 (95% CI -10.18 to 6.19) | Better with non-pMDI |
| Wang 2020 | 12-24 weeks | SGRQ | MD 1 (95% CI -3.14 to 5.14) (difference in least square means) | Worse with non-pMDI |

#### Symptom control

*Figure S10. Symptom control in (A) Asthma Maintenance (SMD, 8-30 weeks), (B) Acute Asthma Exacerbations (Modified Wood Clinical Asthma Score) and (C) COPD (CAT score)*

**A**

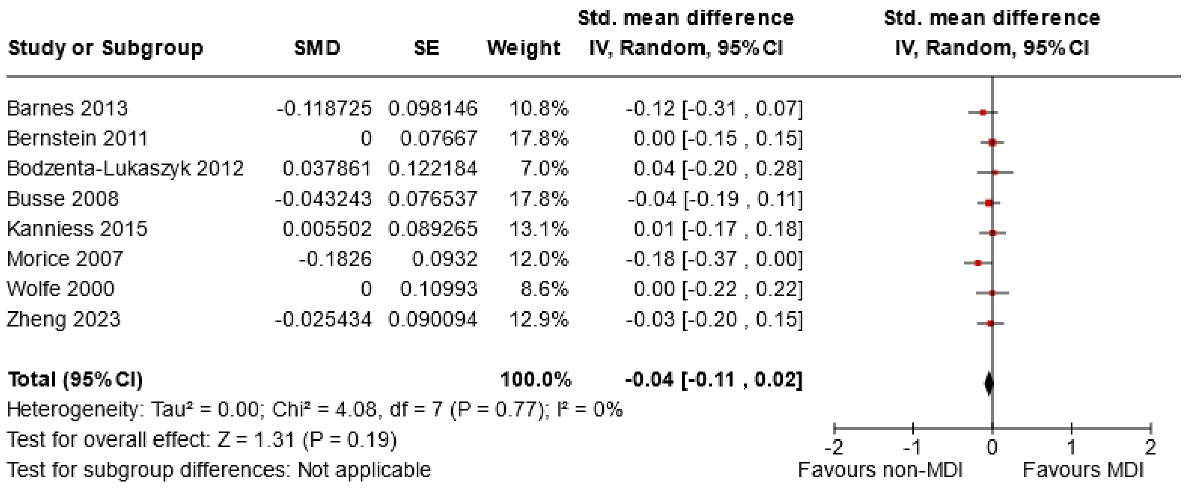

**B**

| Study or Subgroup | Non-MDI Mean | Non-MDI SD | Non-MDI Total | MDI Mean | MDI SD | MDI Total | Mean difference IV, Random, 95% CI |
|---|---|---|---|---|---|---|---|
| Vangveeravong 2008 | 1.6 | 0.90913729 | 18 | 1.7 | 0.995721794 | 18 | -0.10 [-0.72 , 0.52] |

Mean difference IV, Random, 95% CI

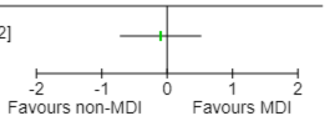

**C**

| Study or Subgroup | Non-MDI Mean | Non-MDI SD | Non-MDI Total | MDI Mean | MDI SD | MDI Total | Mean difference IV, Random, 95% CI |
|---|---|---|---|---|---|---|---|
| Maltais 2019 | -3.56 | 4.948778 | 506 | -2.97 | 4.695743 | 500 | -0.59 [-1.19 , 0.01] |

Mean difference IV, Random, 95% CI

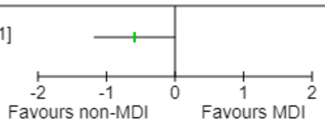

*Table S12 Asthma maintenance and acute asthma exacerbations: Additional symptom control results not included in meta-analysis for asthma maintenance*

| Study ID | Timepoint | Outcome measure | Effect estimate | Direction of effect |
|---|---|---|---|---|
| **Asthma maintenance** | | | | |
| Bateman 2001 | 12 weeks | People with symptom free days | RR 0.94 (95% CI 0.77 to 1.15) | Worse with non-pMDI |
| Lundback 1994 | 4 weeks | People with median day symptom score >2 (lower is better) | RR 0.75 (95% CI 0.38 to 1.49) | Better with non-pMDI |
| Papi 2012 | 26 weeks | Controlled & partly controlled asthma (defined using a composite) (%) (higher is better) | RR 1.01 (95% CI, 0.94 to 1.10) | Better with non-pMDI |
| Poukkula 1998 | 12 weeks | Severity sum scores (lower is better) | MD 0.9 (no measure of variance) | Worse with non-pMDI |
| Srichana 2016 | 13 weeks | Symptom free days during study period | MD 6.6 (95% CI -10.64 to 23.84) | Better with non-pMDI |
| Van Noord 2001 | 12 weeks | Symptom-free days (higher is better) | Difference in medians -12 (no measure of variance) | Worse with non-pMDI |
| **Acute asthma exacerbations** | | | | |
| Direkwatanachai 2011 | 60 min | Modified Wood Clinical Asthma Score – no. with a score that has fallen by >=50%; or raw score <=3 | Relative risk 1.00 [0.81, 1.23] | None |

#### Reliever use

*Figure S11 Reliever Use in (A) Asthma Maintenance (SMD) and (B) COPD (puffs/day)*

**A**

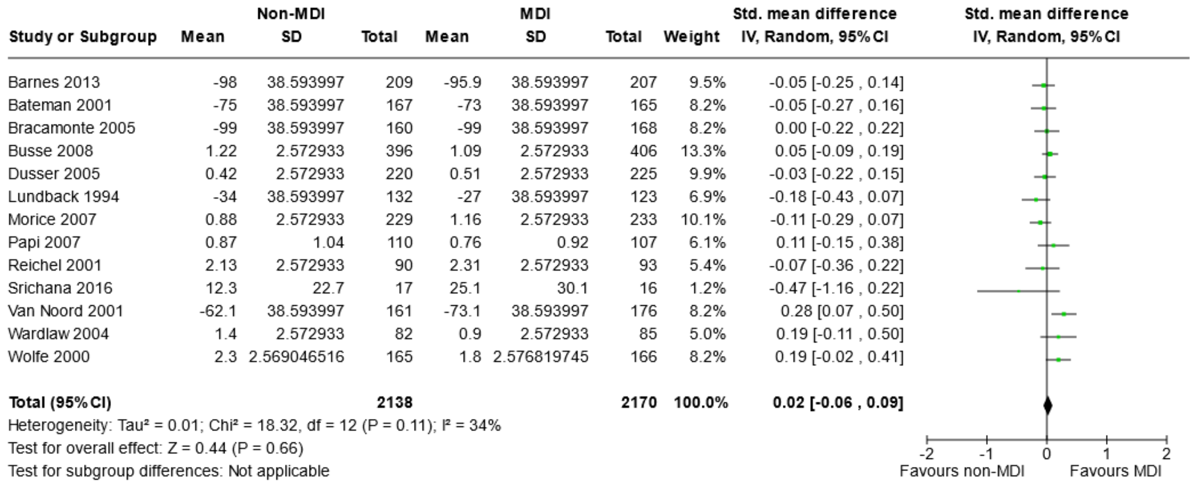

**B**

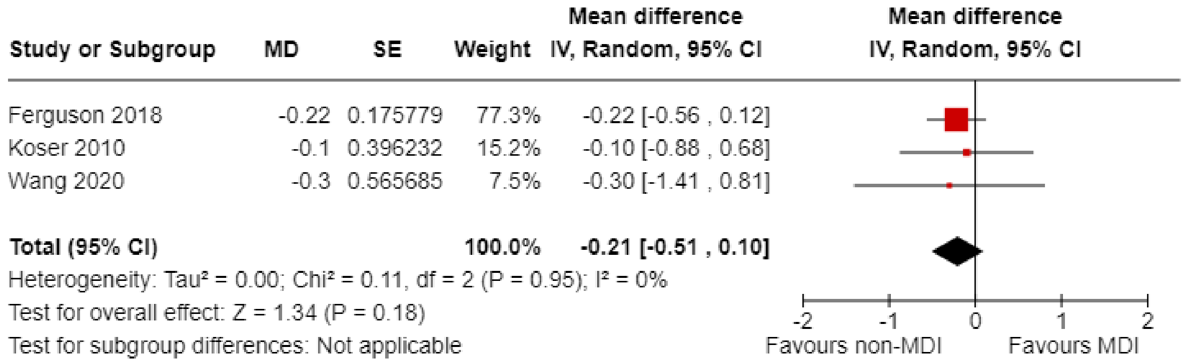

*Table S13 Asthma maintenance: Additional reliever control results not included in meta-analysis*

| Study ID | Timepoint | Outcome measure | Effect estimate | Direction of effect |
|---|---|---|---|---|
| Bernstein 2011 | 12 weeks | Change in proportion of nights with nocturnal awakenings due to asthma that required use of a SABA | MD -0.02 (no measure of variance) | Better with non-pMDIs |
| Bodzenta-Lukaszyk 2012 | 12 weeks | Change in rescue medication free days | Least square MD -2.58% (95% CI -10.25% to 5.09%) | Worse with non-pMDIs |
| Kanniess 2015 | 12 weeks | Change in average use of rescue | MD 0 (95%CI -0.09 to 0.08) | No difference |

| | | | | |
|---|---|---|---|---|
| | | medication (number of inhalations/day) | | |
| Lundback 1993 | 6 weeks | Patients with same/reduced requirement for rescue medication-days (higher is better) | RR 1.0 (95%CI 0.91 to 1.11) | No difference |
| Papi 2012 | 24 weeks | Day time: Number of patients with inhaled rescue salbutamol-free days in study period (6/12) (higher is better) | RR 0.94 (95%CI 0.77 to 1.16) | Worse with non-pMDIs |
| Zheng 2023 | 12 weeks | Change in rescue medication-free days | MD -0.29 (95%CI -1.27 to 0.7) | Worse with non-pMDIs |

#### Mortality

*Figure S12. Mortality in COPD*

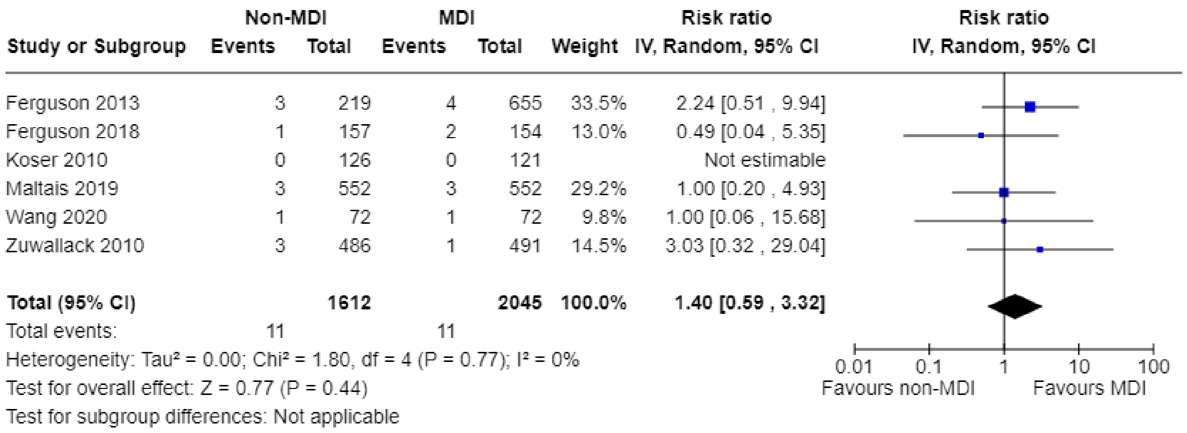

#### Subgroup analyses

*Figure S13. Subgroup analysis of FEV1 by age of participants in Asthma Maintenance*

| Study or Subgroup | SMD | SE | Weight | Std. mean difference IV, Random, 95% CI |
|---|---|---|---|---|
| **1.12.1 Adults and adolescents ≥ 13 years** | | | | |
| Barnes 2013 | 0 | 0.098059 | 5.5% | 0.00 [-0.19 , 0.19] |
| Bateman 2001 | 0.077946 | 0.110147 | 4.5% | 0.08 [-0.14 , 0.29] |
| Bernstein 2011 | -0.0486 | 0.0745 | 8.4% | -0.05 [-0.19 , 0.10] |
| Bodzenta-Lukaszyk 2012 | 0.047178 | 0.127571 | 3.5% | 0.05 [-0.20 , 0.30] |
| Bronsky 1987 | -0.108921 | 0.148011 | 2.7% | -0.11 [-0.40 , 0.18] |
| Busse 2008 | 0.140491 | 0.071382 | 8.9% | 0.14 [0.00 , 0.28] |
| Dusser 2005 | -0.03503 | 0.094822 | 5.8% | -0.04 [-0.22 , 0.15] |
| Koskela 2000 | 0.097028 | 0.167025 | 2.2% | 0.10 [-0.23 , 0.42] |
| Lundback 1993 | 0.280647 | 0.10713 | 4.7% | 0.28 [0.07 , 0.49] |
| Lundback 1994 | -0.077926 | 0.116303 | 4.1% | -0.08 [-0.31 , 0.15] |
| Nelson 1999 | -0.210924 | 0.158576 | 2.4% | -0.21 [-0.52 , 0.10] |
| Papi 2007 | 0.109479 | 0.136192 | 3.1% | 0.11 [-0.16 , 0.38] |
| Papi 2012 | -0.014021 | 0.097387 | 5.6% | -0.01 [-0.20 , 0.18] |
| Poukkula 1998 | -0.025114 | 0.164406 | 2.2% | -0.03 [-0.35 , 0.30] |
| Reichel 2001 | 0.272608 | 0.144661 | 2.8% | 0.27 [-0.01 , 0.56] |
| Van Noord 2001 | 0.055997 | 0.116549 | 4.1% | 0.06 [-0.17 , 0.28] |
| Vincken 2004 | 0.0667 | 0.1161 | 4.1% | 0.07 [-0.16 , 0.29] |
| Wardlaw 2004 | -0.216554 | 0.155253 | 2.5% | -0.22 [-0.52 , 0.09] |
| Wolfe 2000 | -0.009694 | 0.109931 | 4.5% | -0.01 [-0.23 , 0.21] |
| Zheng 2023 | -0.067552 | 0.090664 | 6.2% | -0.07 [-0.25 , 0.11] |
| **Subtotal (Wald[a])** | | | **87.9%** | **0.02 [-0.03 , 0.08]** |
| Test for overall effect: Z = 0.90 (P = 0.37) | | | | |
| Heterogeneity: Tau² (DL[b]) = 0.00; Chi² = 21.35, df = 19 (P = 0.32); I² = 11% | | | | |
| **1.12.2 Children < 13 years** | | | | |
| Amar 2017 | -0.167181 | 0.130495 | 3.4% | -0.17 [-0.42 , 0.09] |
| Bracamonte 2005 | -0.129806 | 0.110582 | 4.5% | -0.13 [-0.35 , 0.09] |
| Von Berg 2004[c] | 0.117 | 0.1141 | 4.3% | 0.12 [-0.11 , 0.34] |
| **Subtotal (Wald[a])** | | | **12.1%** | **-0.06 [-0.23 , 0.12]** |
| Test for overall effect: Z = 0.62 (P = 0.54) | | | | |
| Heterogeneity: Tau² (DL[b]) = 0.01; Chi² = 3.47, df = 2 (P = 0.18); I² = 42% | | | | |
| **Total (Wald[a])** | | | **100.0%** | **0.01 [-0.04 , 0.06]** |

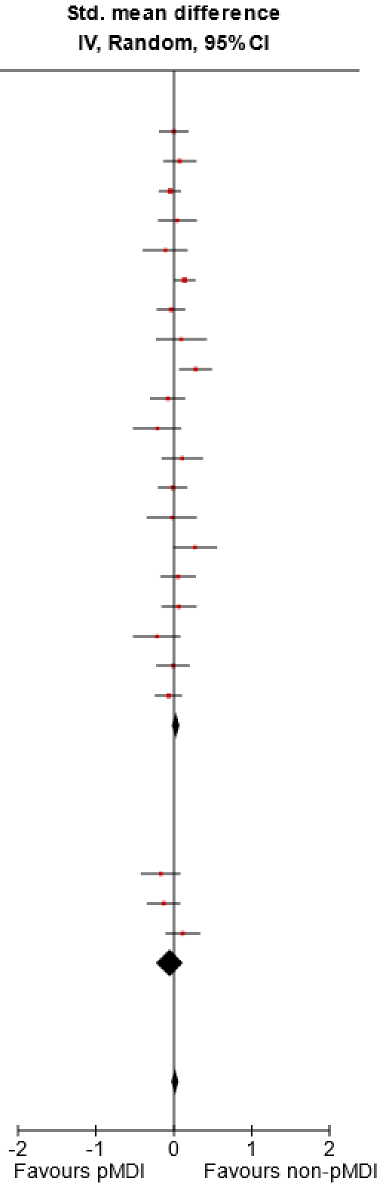

Test for overall effect: Z = 0.55 (P = 0.58)
Test for subgroup differences: Chi² = 0.72, df = 1 (P = 0.40), I² = 0%
Heterogeneity: Tau² (DL[b]) = 0.00; Chi² = 25.96, df = 22 (P = 0.25); I² = 15%

**Footnotes**

[a]CI calculated by Wald-type method.
[b]Tau² calculated by DerSimonian and Laird method.
[c]Von Berg included children up to 15 years.

*Figure S14. Subgroup analysis of FEV1 by non-pMDI device type in COPD*

| Study or Subgroup | SMD | SE | Weight | Std. mean difference IV, Random, 95% CI |
|---|---|---|---|---|
| **3.11.1 DPI** | | | | |
| Ferguson 2018 | 0.0422 | 0.0781 | 17.6% | 0.04 [-0.11 , 0.20] |
| Koser 2010 | -0.021482 | 0.138361 | 5.6% | -0.02 [-0.29 , 0.25] |
| Maltais 2019 | 0.007 | 0.0602 | 29.6% | 0.01 [-0.11 , 0.12] |
| Wang 2020 | -0.052626 | 0.172206 | 3.6% | -0.05 [-0.39 , 0.28] |
| **Subtotal (Wald[a])** | | | **56.5%** | **0.01 [-0.07 , 0.10]** |
| Test for overall effect: Z = 0.26 (P = 0.80) | | | | |
| Heterogeneity: Tau$^2$ (DL[b]) = 0.00; Chi$^2$ = 0.36, df = 3 (P = 0.95); $I^2$ = 0% | | | | |
| **3.11.2 SMI** | | | | |
| Ferguson 2013 | 0.2752 | 0.1271 | 6.6% | 0.28 [0.03 , 0.52] |
| Kilfeather 2004 | 0.0042 | 0.0979 | 11.2% | 0.00 [-0.19 , 0.20] |
| Zuwallack 2010 | 0.017138 | 0.064688 | 25.7% | 0.02 [-0.11 , 0.14] |
| **Subtotal (Wald[a])** | | | **43.5%** | **0.07 [-0.07 , 0.21]** |
| Test for overall effect: Z = 0.98 (P = 0.33) | | | | |
| Heterogeneity: Tau$^2$ (DL[b]) = 0.01; Chi$^2$ = 3.61, df = 2 (P = 0.16); $I^2$ = 45% | | | | |
| **Total (Wald[a])** | | | **100.0%** | **0.03 [-0.03 , 0.09]** |

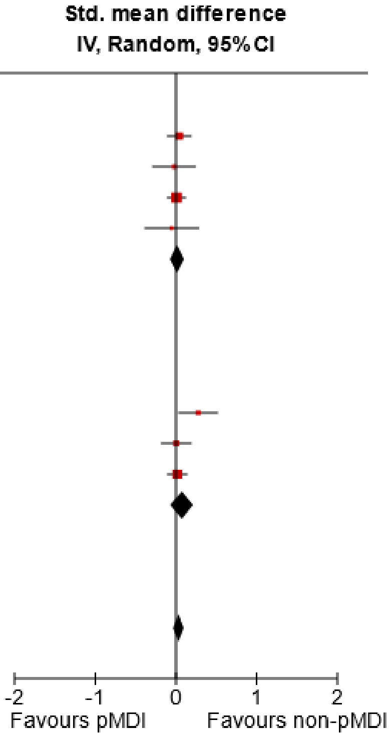

Test for overall effect: Z = 0.90 (P = 0.37)
Test for subgroup differences: Chi$^2$ = 0.50, df = 1 (P = 0.48), $I^2$ = 0%
Heterogeneity: Tau$^2$ (DL[b]) = 0.00; Chi$^2$ = 4.37, df = 6 (P = 0.63); $I^2$ = 0%

**Footnotes**
[a]CI calculated by Wald-type method.
[b]Tau$^2$ calculated by DerSimonian and Laird method.

*Figure S15. Subgroup analysis of FEV1 by manufacturer funding in Asthma Maintenance*

| Study or Subgroup | SMD | SE | Weight | Std. mean difference IV, Random, 95% CI |
|---|---|---|---|---|
| **1.11.1 pMDI funded by manufacturer** | | | | |
| Barnes 2013 | 0 | 0.098059 | 5.5% | 0.00 [-0.19 , 0.19] |
| Bernstein 2011 | -0.0486 | 0.0745 | 8.4% | -0.05 [-0.19 , 0.10] |
| Bodzenta-Lukaszyk 2012 | 0.047178 | 0.127571 | 3.5% | 0.05 [-0.20 , 0.30] |
| Busse 2008 | 0.140491 | 0.071382 | 8.9% | 0.14 [0.00 , 0.28] |
| Dusser 2005 | -0.03503 | 0.094822 | 5.8% | -0.04 [-0.22 , 0.15] |
| Papi 2007 | 0.109479 | 0.136192 | 3.1% | 0.11 [-0.16 , 0.38] |
| Papi 2012 | -0.014021 | 0.097387 | 5.6% | -0.01 [-0.20 , 0.18] |
| Reichel 2001 | 0.272608 | 0.144661 | 2.8% | 0.27 [-0.01 , 0.56] |
| **Subtotal (Wald[a])** | | | **43.5%** | **0.04 [-0.03 , 0.11]** |
| Test for overall effect: Z = 1.17 (P = 0.24) | | | | |
| Heterogeneity: Tau² (DL[b]) = 0.00; Chi² = 7.34, df = 7 (P = 0.39); I² = 5% | | | | |
| **1.11.2 Non-pMDI funded by manufacturer** | | | | |
| Amar 2017 | -0.167181 | 0.130495 | 3.4% | -0.17 [-0.42 , 0.09] |
| Bateman 2001 | 0.077946 | 0.110147 | 4.5% | 0.08 [-0.14 , 0.29] |
| Bracamonte 2005 | -0.129806 | 0.110582 | 4.5% | -0.13 [-0.35 , 0.09] |
| Bronsky 1987 | -0.108921 | 0.148011 | 2.7% | -0.11 [-0.40 , 0.18] |
| Koskela 2000 | 0.097028 | 0.167025 | 2.2% | 0.10 [-0.23 , 0.42] |
| Lundback 1993 | 0.280647 | 0.10713 | 4.7% | 0.28 [0.07 , 0.49] |
| Lundback 1994 | -0.077926 | 0.116303 | 4.1% | -0.08 [-0.31 , 0.15] |
| Nelson 1999 | -0.210924 | 0.158576 | 2.4% | -0.21 [-0.52 , 0.10] |
| Poukkula 1998 | -0.025114 | 0.164406 | 2.2% | -0.03 [-0.35 , 0.30] |
| Van Noord 2001 | 0.055997 | 0.116549 | 4.1% | 0.06 [-0.17 , 0.28] |
| Vincken 2004 | 0.0667 | 0.1161 | 4.1% | 0.07 [-0.16 , 0.29] |
| Von Berg 2004 | 0.117 | 0.1141 | 4.3% | 0.12 [-0.11 , 0.34] |
| Wardlaw 2004 | -0.216554 | 0.155253 | 2.5% | -0.22 [-0.52 , 0.09] |
| Wolfe 2000 | -0.009694 | 0.109931 | 4.5% | -0.01 [-0.23 , 0.21] |
| Zheng 2023 | -0.067552 | 0.090664 | 6.2% | -0.07 [-0.25 , 0.11] |
| **Subtotal (Wald[a])** | | | **56.5%** | **-0.01 [-0.08 , 0.06]** |
| Test for overall effect: Z = 0.23 (P = 0.81) | | | | |
| Heterogeneity: Tau² (DL[b]) = 0.00; Chi² = 17.59, df = 14 (P = 0.23); I² = 20% | | | | |
| **Total (Wald[a])** | | | **100.0%** | **0.01 [-0.04 , 0.06]** |

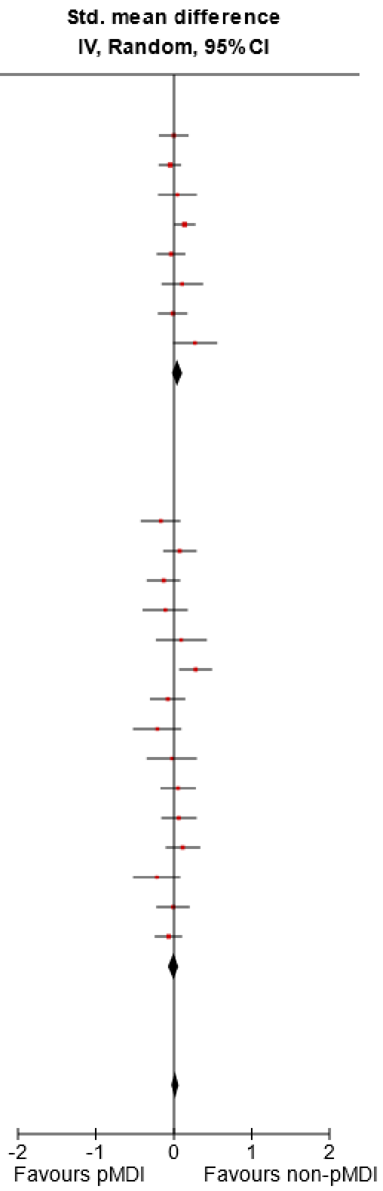

Test for overall effect: Z = 0.55 (P = 0.58)
Test for subgroup differences: Chi² = 0.98, df = 1 (P = 0.32), I² = 0%
Heterogeneity: Tau² (DL[b]) = 0.00; Chi² = 25.96, df = 22 (P = 0.25); I² = 15%

**Footnotes**
[a]CI calculated by Wald-type method.
[b]Tau² calculated by DerSimonian and Laird method.

*Figure S16. Subgroup analysis of adverse events (risk of >1) by use of 'double-dummy' study design in (A) Asthma Maintenance and (B) COPD*

**A**

| Study or Subgroup | log[RR] | SE | Weight | Risk ratio IV, Random, 95% CI |
|---|---|---|---|---|
| **Double dummy** | | | | |
| Amar 2017 | -0.218645 | 0.162133 | 2.1% | 0.80 [0.58 , 1.10] |
| Barnes 2013 | -0.186962 | 0.597818 | 0.2% | 0.83 [0.26 , 2.68] |
| Bateman 2001 | 0.135109 | 0.103309 | 5.1% | 1.14 [0.93 , 1.40] |
| Bodzenta-Lukaszyk 2012 | -0.102031 | 0.242089 | 0.9% | 0.90 [0.56 , 1.45] |
| Bracamonte 2005 | -0.012394 | 0.111336 | 4.4% | 0.99 [0.79 , 1.23] |
| Bronsky 1987 | -0.359067 | 0.276949 | 0.7% | 0.70 [0.41 , 1.20] |
| Dusser 2005 | -0.117107 | 0.115256 | 4.1% | 0.89 [0.71 , 1.11] |
| Koskela 2000 | -0.183574 | 0.148271 | 2.5% | 0.83 [0.62 , 1.11] |
| Lundback 1993 | -0.13438 | 0.107572 | 4.7% | 0.87 [0.71 , 1.08] |
| Lundback 1994 | -0.027029 | 0.194335 | 1.4% | 0.97 [0.67 , 1.42] |
| Morice 2007 | -0.041524 | 0.144148 | 2.6% | 0.96 [0.72 , 1.27] |
| Nelson 1999 | 0.064861 | 0.112033 | 4.4% | 1.07 [0.86 , 1.33] |
| Papi 2007 | 0.182322 | 0.322294 | 0.5% | 1.20 [0.64 , 2.26] |
| Stradling 2000 | -0.00174 | 0.340117 | 0.5% | 1.00 [0.51 , 1.94] |
| Van Noord 2001 | -0.163756 | 0.111552 | 4.4% | 0.85 [0.68 , 1.06] |
| Zheng 2023 | -0.023962 | 0.091813 | 6.5% | 0.98 [0.82 , 1.17] |
| **Subtotal (95% CI)** | | | **45.1%** | **0.95 [0.89 , 1.01]** |
| Heterogeneity: Tau² (DL{fn}) = 0.00; Chi² = 10.23, df = 15 (P = 0.80); I² = 0% | | | | |
| Test for overall effect: Z = 1.54 (P = 0.12) | | | | |
| **No double dummy** | | | | |
| Busse 2008 | -0.049453 | 0.05656 | 17.1% | 0.95 [0.85 , 1.06] |
| Morice 2008 | 0.03502 | 0.045595 | 26.3% | 1.04 [0.95 , 1.13] |
| Papi 2012 | -0.079151 | 0.232431 | 1.0% | 0.92 [0.59 , 1.46] |
| Reichel 2001 | -0.119025 | 0.180528 | 1.7% | 0.89 [0.62 , 1.26] |
| Srichana 2016 | 0 | 1.374369 | 0.0% | 1.00 [0.07 , 14.79] |
| Vincken 2004 | 0.089629 | 0.089224 | 6.9% | 1.09 [0.92 , 1.30] |
| Von Berg 2004 | -0.349952 | 0.169462 | 1.9% | 0.70 [0.51 , 0.98] |
| **Subtotal (95% CI)** | | | **54.9%** | **0.99 [0.91 , 1.07]** |
| Heterogeneity: Tau² (DL{fn}) = 0.00; Chi² = 7.16, df = 6 (P = 0.31); I² = 16% | | | | |
| Test for overall effect: Z = 0.31 (P = 0.75) | | | | |
| **Total (95% CI)** | | | **100.0%** | **0.97 [0.93 , 1.02]** |
| Heterogeneity: Tau² = 0.00; Chi² = 18.48, df = 22 (P = 0.68); I² = 0% | | | | |
| Test for overall effect: Z = 1.14 (P = 0.25) | | | | |
| Test for subgroup differences: Chi² = 0.62, df = 1 (P = 0.43), I² = 0% | | | | |

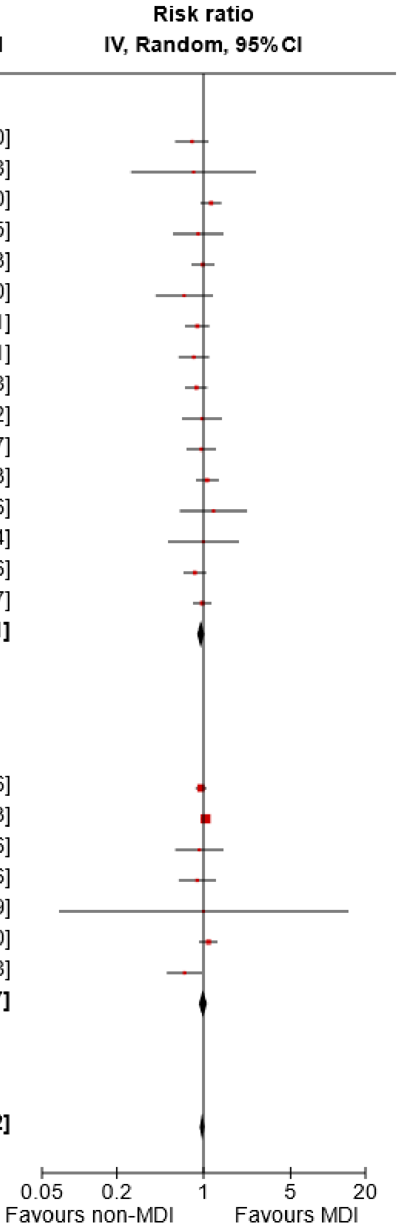

**B**

| | Non-MDI | | MDI | | | Risk ratio |
|---|---|---|---|---|---|---|
| **Study or Subgroup** | **Events** | **Total** | **Events** | **Total** | **Weight** | **IV, Random, 95% CI** |
| **Double dummy** | | | | | | |
| Koser 2010 | 59 | 126 | 54 | 126 | 7.5% | 1.09 [0.83 , 1.44] |
| Maltais 2019 | 248 | 552 | 226 | 552 | 18.3% | 1.10 [0.96 , 1.26] |
| Zuwallack 2010 | 222 | 486 | 254 | 491 | 19.3% | 0.88 [0.78 , 1.00] |
| **Subtotal (95% CI)** | | **1164** | | **1169** | **45.1%** | **1.01 [0.85 , 1.18]** |
| Total events: | 529 | | 534 | | | |
| Heterogeneity: Tau² = 0.01; Chi² = 5.70, df = 2 (P = 0.06); I² = 65% | | | | | | |
| Test for overall effect: Z = 0.06 (P = 0.95) | | | | | | |
| **No double dummy** | | | | | | |
| Ferguson 2013 | 109 | 157 | 112 | 154 | 17.6% | 0.95 [0.83 , 1.10] |
| Ferguson 2018 | 114 | 219 | 333 | 655 | 16.9% | 1.02 [0.88 , 1.19] |
| Kilfeather 2004 | 125 | 224 | 115 | 220 | 14.3% | 1.07 [0.90 , 1.27] |
| Wang 2020 | 44 | 72 | 32 | 72 | 5.9% | 1.38 [1.00 , 1.89] |
| **Subtotal (95% CI)** | | **672** | | **1101** | **54.9%** | **1.04 [0.94 , 1.16]** |
| Total events: | 392 | | 592 | | | |
| Heterogeneity: Tau² = 0.00; Chi² = 4.46, df = 3 (P = 0.22); I² = 33% | | | | | | |
| Test for overall effect: Z = 0.75 (P = 0.46) | | | | | | |
| **Total (95% CI)** | | **1836** | | **2270** | **100.0%** | **1.02 [0.94 , 1.11]** |
| Total events: | 921 | | 1126 | | | |
| Heterogeneity: Tau² = 0.01; Chi² = 10.56, df = 6 (P = 0.10); I² = 43% | | | | | | |
| Test for overall effect: Z = 0.55 (P = 0.58) | | | | | | |
| Test for subgroup differences: Chi² = 0.13, df = 1 (P = 0.72), I² = 0% | | | | | | |

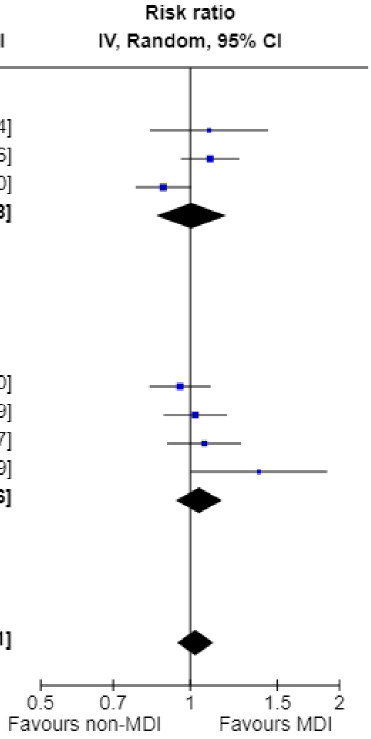

*Figure S17. Subgroup analysis of serious adverse events (risk of >1) by use of ‘double-dummy’ study design in (A) Asthma Maintenance and (B) COPD*

**A**

| Study or Subgroup | Non-MDI Events | Non-MDI Total | MDI Events | MDI Total | Weight | Risk ratio IV, Random, 95% CI |
|---|---|---|---|---|---|---|
| **Double dummy** | | | | | | |
| Amar 2017 | 2 | 122 | 1 | 114 | 1.8% | 1.87 [0.17 , 20.33] |
| Barnes 2013 | 2 | 209 | 2 | 207 | 2.6% | 0.99 [0.14 , 6.97] |
| Bateman 2001 | 3 | 167 | 3 | 165 | 4.0% | 0.99 [0.20 , 4.82] |
| Bodzenta-Lukaszyk 2012 | 2 | 139 | 1 | 140 | 1.8% | 2.01 [0.18 , 21.96] |
| Bracamonte 2005 | 2 | 160 | 2 | 168 | 2.7% | 1.05 [0.15 , 7.37] |
| Bronsky 1987 | 3 | 115 | 5 | 116 | 5.1% | 0.61 [0.15 , 2.47] |
| Dusser 2005 | 3 | 221 | 6 | 227 | 5.3% | 0.51 [0.13 , 2.03] |
| Koskela 2000 | 1 | 68 | 0 | 76 | 1.0% | 3.35 [0.14 , 80.83] |
| Lundback 1994 | 7 | 150 | 5 | 146 | 8.0% | 1.36 [0.44 , 4.20] |
| Morice 2007 | 0 | 229 | 2 | 233 | 1.1% | 0.20 [0.01 , 4.22] |
| Nelson 1999 | 1 | 97 | 1 | 92 | 1.3% | 0.95 [0.06 , 14.94] |
| Papi 2007 | 0 | 109 | 0 | 109 | | Not estimable |
| Van Noord 2001 | 3 | 161 | 5 | 176 | 5.0% | 0.66 [0.16 , 2.70] |
| Zheng 2023 | 4 | 252 | 4 | 242 | 5.3% | 0.96 [0.24 , 3.80] |
| **Subtotal (95% CI)** | | **2199** | | **2211** | **45.0%** | **0.91 [0.57 , 1.46]** |
| Total events: | 33 | | 37 | | | |
| Heterogeneity: Tau² (DL{fn}) = 0.00; Chi² = 4.09, df = 12 (P = 0.98); I² = 0% | | | | | | |
| Test for overall effect: Z = 0.39 (P = 0.70) | | | | | | |
| **No double dummy** | | | | | | |
| Bernstein 2011 | 5 | 351 | 5 | 371 | 6.6% | 1.06 [0.31 , 3.62] |
| Busse 2008 | 9 | 406 | 10 | 427 | 12.7% | 0.95 [0.39 , 2.31] |
| Morice 2008 | 15 | 227 | 32 | 446 | 28.7% | 0.92 [0.51 , 1.66] |
| Papi 2012 | 3 | 216 | 1 | 206 | 2.0% | 2.86 [0.30 , 27.28] |
| Reichel 2001 | 0 | 93 | 0 | 92 | | Not estimable |
| Vincken 2004 | 5 | 161 | 2 | 159 | 3.8% | 2.47 [0.49 , 12.54] |
| Von Berg 2004 | 0 | 180 | 3 | 177 | 1.2% | 0.14 [0.01 , 2.70] |
| Wardlaw 2004 | 0 | 82 | 0 | 85 | | Not estimable |
| **Subtotal (95% CI)** | | **1716** | | **1963** | **55.0%** | **1.01 [0.66 , 1.55]** |
| Total events: | 37 | | 53 | | | |
| Heterogeneity: Tau² (DL{fn}) = 0.00; Chi² = 3.81, df = 5 (P = 0.58); I² = 0% | | | | | | |
| Test for overall effect: Z = 0.05 (P = 0.96) | | | | | | |
| **Total (95% CI)** | | **3915** | | **4174** | **100.0%** | **0.96 [0.70 , 1.32]** |
| Total events: | 70 | | 90 | | | |
| Heterogeneity: Tau² = 0.00; Chi² = 8.00, df = 18 (P = 0.98); I² = 0% | | | | | | |
| Test for overall effect: Z = 0.22 (P = 0.82) | | | | | | |
| Test for subgroup differences: Chi² = 0.10, df = 1 (P = 0.75), I² = 0% | | | | | | |

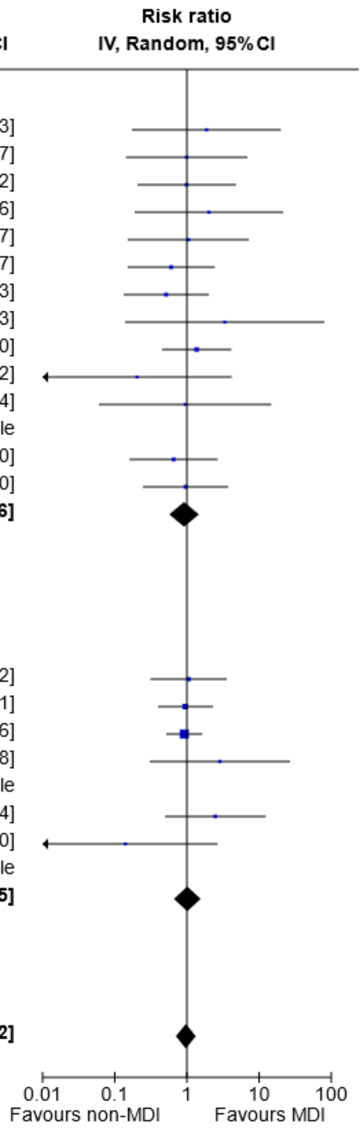

**B**

| Study or Subgroup | Non-MDI Events | Non-MDI Total | MDI Events | MDI Total | Weight | Risk ratio IV, Random, 95% CI |
|---|---|---|---|---|---|---|
| **Double dummy** | | | | | | |
| Maltais 2019 | 40 | 552 | 32 | 552 | 24.8% | 1.25 [0.80 , 1.96] |
| Zuwallack 2010 | 17 | 486 | 33 | 491 | 21.8% | 0.52 [0.29 , 0.92] |
| **Subtotal (95% CI)** | | **1038** | | **1043** | **46.6%** | **0.82 [0.35 , 1.94]** |
| Total events: | 57 | | 65 | | | |
| Heterogeneity: Tau² = 0.32; Chi² = 5.58, df = 1 (P = 0.02); I² = 82% | | | | | | |
| Test for overall effect: Z = 0.45 (P = 0.65) | | | | | | |
| **No double dummy** | | | | | | |
| Ferguson 2013 | 23 | 157 | 20 | 154 | 22.2% | 1.13 [0.65 , 1.97] |
| Ferguson 2018 | 20 | 219 | 42 | 655 | 23.3% | 1.42 [0.86 , 2.37] |
| Wang 2020 | 11 | 72 | 2 | 72 | 7.9% | 5.50 [1.26 , 23.94] |
| **Subtotal (95% CI)** | | **448** | | **881** | **53.4%** | **1.53 [0.86 , 2.70]** |
| Total events: | 54 | | 64 | | | |
| Heterogeneity: Tau² = 0.12; Chi² = 3.91, df = 2 (P = 0.14); I² = 49% | | | | | | |
| Test for overall effect: Z = 1.45 (P = 0.15) | | | | | | |
| **Total (95% CI)** | | **1486** | | **1924** | **100.0%** | **1.17 [0.73 , 1.89]** |
| Total events: | 111 | | 129 | | | |
| Heterogeneity: Tau² = 0.19; Chi² = 12.48, df = 4 (P = 0.01); I² = 68% | | | | | | |
| Test for overall effect: Z = 0.64 (P = 0.52) | | | | | | |
| Test for subgroup differences: Chi² = 1.39, df = 1 (P = 0.24), I² = 27.9% | | | | | | |

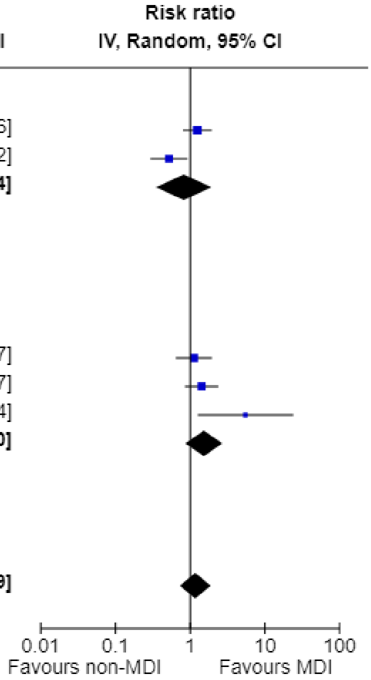

#### Section F. Data characteristics

##### Table S14 – Funder for each Study

| Publication | Manufacturer funded? | Company | Which arm(s)? |
|---|---|---|---|
| Asthma Maintenance | | | |
| Amar 2017 | Yes | Merck & Co. | DPI only |
| Barnes 2013 | Yes | Chiesi | pMDI only |
| Bateman 2001 | Yes | Glaxo Wellcome | DPI only |
| Bernstein 2011 | Yes | Merck & Co. | pMDI only |
| Bodzenta-Lukaszyk 2012 | Yes | Mundipharma Research Limited | pMDI only |
| Bracamonte 2005 | Yes | GlaxoSmithKline | DPI only |
| Bronsky 1987 | Yes | Glaxo | DPI only |
| Busse 2008 / O'Connor 2010 | Yes | AstraZeneca | pMDI only |
| Dusser 2005 | Yes | Chiesi | pMDI only |
| Kanniess 2015 | Yes | Chiesi | Both |
| Kemp 1989 | Unclear* | Unclear* | Unclear* |
| Koskela 2000 | Yes | Orion Pharma | DPI only |
| Lundback 1993 | Yes | Glaxo | DPI only |
| Lundback 1994 | Yes | Glaxo | DPI only |
| Morice 2007 | Yes | AstraZeneca | Both |
| Morice 2008 | Yes | AstraZeneca | Both |
| Nelson 1999 | Yes | Dura Pharmaceuticals | DPI only |
| Papi 2007 | Yes | Chiesi | pMDI only |
| Papi 2012 | Yes | Chiesi | pMDI only |
| Pauwels 1996 | Yes | Astra Draco | DPI only |
| Poukkula 1998 | Yes | Orion Pharma | DPI only |
| Reichel 2001 | Yes | 3M pharmaceuticals | pMDI only |
| Srichana 2016 | No | N/A | N/A |
| Stradling, 2000 | Yes | Innovata Biomed Ltd | DPI only |
| Van Noord 2001 | Yes | Glaxo | DPI only |
| Vincken 2004 | Yes | Boehringer Ingelheim | SMI only |

| | | | |
|---|---|---|---|
| von Berg 2004 | Yes | Boehringer Ingelheim | SMI only |
| Wardlaw 2004 | Yes | Schering-Plough | DPI only |
| Wolfe 2000 | Yes | Glaxo | DPI only |
| Zheng 2023 | Yes | Chiesi | DPI only |
| Asthma Acute | | | |
| Direkwatanachai 2011 | Yes | Harn Thai | DPI only |
| Drblik 2003 | Yes | AstraZeneca | DPI only |
| Khaled 2014 | No | N/A | N/A |
| Lodha 2004 | No | N/A | N/A |
| Vangveeravong 2008 | No | N/A | N/A |
| COPD | | | |
| Ferguson 2013 | Yes | Boehringer Ingelheim | SMI only |
| Ferguson 2018 | Yes | Pearl (member of AstraZeneca Group) | DPI only |
| Kilfeather 2004 | Yes | Boehringer Ingelheim | SMI only |
| Koser 2010 | Yes | GlaxoSmithKline | DPI only |
| Maltais 2019 | Yes | AstraZeneca | MDI only |
| Wang 2020 | Yes | AstraZeneca | Both |
| Zuwallack 2010 | Yes | Boehringer Ingelheim | Both |

* No information available concerning funding for this study. A Glaxo device (Rotahaler) was used in the DPI arm.

#### Table S15 – Measured Outcomes in each Study

**Key to 'Included in meta-analysis' column:**

Yes: the study was included in the meta-analysis

No (not used): the study was included in the meta-analysis, but used a different value/time point

No: the study was excluded from meta-analysis due to differences in outcome measurement; values instead presented in Tables S8-S13 above

| Publication | How outcome measured | Time point assessed | Change from baseline or endpoint value | Included in meta-analysis |
|---|---|---|---|---|
| **Outcome: Forced Expiratory Volume in 1 second ($FEV_1$)** | | | | |
| Asthma Maintenance | | | | |
| Amar 2017 | % predicted value | 12 weeks | Endpoint value | Yes |
| Barnes 2013 | % predicted value | 12 weeks | Endpoint value | Yes |
| Bateman 2001 | Litres | 12 weeks | Endpoint value | Yes |
| Bateman 2001 | Litres | 12 weeks | Change from baseline | No (not used) |
| Bernstein 2011 | Litres | 12 weeks | Endpoint value | Yes |
| Bodzenta-Lukaszyk 2012 | Litres | 12 weeks | Endpoint value | Yes |
| Bracamonte 2005 | Litres | 12 weeks | Endpoint value | Yes |
| Bronsky 1987 | Litres | 12 weeks | Endpoint value | Yes |
| O'Connor 2010 | Litres | 28 weeks | Endpoint value | Yes |
| Dusser 2005 | Litres | 12 weeks | Endpoint value | Yes |
| Kanniess 2015 | Litres | 8 weeks | Endpoint value | Yes |
| Kemp 1989 | Litres | 12 weeks | Endpoint value | Yes |
| Kemp 1989 | % predicted value | 12 weeks | Change from baseline | No (not used) |
| Koskela 2000 | % predicted value | 8 weeks | Endpoint value | Yes |
| Lundback 1993 | Litres | 6 weeks | Endpoint value | Yes |
| Lundback 1993 | Litres (change) | 6 weeks | Change from baseline | No (not used) |
| Lundback 1994 | Litres | 4 weeks | Endpoint value | Yes |
| Morice 2007 | Litres | 12 weeks | Endpoint value | Yes |
| Morice 2008 | Litres | 52 weeks | Endpoint value | Yes |
| Nelson 1999 | Litres | 12 weeks | Endpoint value | Yes |
| Papi 2007 | Litres | 12 weeks | Endpoint value | Yes |

| | | | | |
|---|---|---|---|---|
| Papi 2012 | % predicted value | 23-24 weeks mean | Endpoint value | Yes |
| Papi 2012 | Litres | 23-24 weeks mean | Endpoint value | No (not used) |
| Papi 2012 | % predicted value | 23-24 weeks mean | Change from baseline | No (not used) |
| Papi 2012 | Litres | 23-24 weeks mean | Change from baseline | No (not used) |
| Poukkula 1998 | Litres | 12 weeks | Endpoint value | Yes |
| Reichel 2001 | % predicted value | 6 weeks | Endpoint value | Yes |
| Srichana 2016 | Litres | 12 weeks | Endpoint value | Yes |
| Van Noord 2001 | Litres | 12 weeks | Endpoint value | Yes |
| Van Noord 2001 | % predicted value | 12 weeks | Change from baseline | No (not used) |
| Van Noord 2001 | Litres | 12 weeks | Change from baseline | No (not used) |
| Vincken 2004 | Litres | 12 weeks | Endpoint value | Yes |
| von Berg 2004 | Litres | 4 weeks | Endpoint value | Yes |
| von Berg 2004 | Litres | 4 weeks | Change from baseline | No (not used) |
| Wardlaw 2004 | % predicted value | 8 weeks | Endpoint value | Yes |
| Wolfe 2000 | Litres | 12 weeks | Endpoint value | Yes |
| Zheng 2023 | Litres | 12 weeks | Endpoint value | Yes |
| Zheng 2023 | Litres | 12 weeks | Change from baseline | No (not used) |
| Asthma Acute | | | | |
| Drblik 2003 | % predicted value | 60 minutes | Endpoint value | Yes |
| COPD | | | | |
| Ferguson 2013 | Litres | 48 weeks | Change from baseline | Yes |
| Ferguson 2013 | % predicted value | 48 weeks | endpoint | No (not used) |
| Ferguson 2018 | Litres | 24 weeks | Change from baseline | Yes |
| Kilfeather 2004 | Litres | 12 weeks | Change from baseline | Yes |
| Koser 2010 | Litres | 12 weeks | Change from baseline | Yes |
| Koser 2010 | Litres | 12 weeks | endpoint | No (not used) |
| Maltais 2019 | Litres | 24 weeks | Change from baseline | Yes |
| Wang 2020 | Litres | 12-24 weeks | Change from baseline | Yes |
| Zuwallack 2010 | Litres | 12 weeks | Change from baseline | Yes |
| **Outcome: Peak Expiratory Flow Rate (PEFR)** | | | | |
| Asthma Maintenance | | | | |
| Barnes 2013 | PEFR in L/min | 12 weeks | Endpoint value | Yes |

| | | | | |
|---|---|---|---|---|
| Bateman 2001 | PEFR in L/min | 12 weeks | Change from baseline | Yes |
| Bernstein 2011 | PEFR in L/min | 12 weeks | Endpoint value | Yes |
| Bodzenta-Lukaszyk 2012 | PEFR in L/min | 12 weeks | Endpoint value | Yes |
| Bracamonte 2005 | PEFR in L/min | 12 weeks | Change from baseline | Yes |
| Bronsky 1987 | PEFR in L/min | 12 weeks | Endpoint value | Yes |
| Busse 2008 | PEFR in L/min | 28 weeks | Change from baseline | Yes |
| Dusser 2005 | PEFR in L/min | 12 weeks | Endpoint value | Yes |
| Kanniess 2015 | PEFR in L/min | 8 weeks | Endpoint value | Yes |
| Koskela 2000 | PEFR in L/min | 8 weeks | Endpoint value | Yes |
| Lundback 1993 | PEFR in L/min | 6 weeks | Endpoint value | Yes |
| Lundback 1994 | PEFR in L/min | 4 weeks | Endpoint value | Yes |
| Morice 2007 | PEFR in L/min | 90 days | Change from baseline | Yes |
| Papi 2007 | PEFR in L/min | 12 weeks | Endpoint value | Yes |
| Papi 2012 | PEFR in L/min | 24 weeks | Endpoint value | Yes |
| Pauwels 1996 | number of weeks with mean PEF less than 90% of baseline | 52 weeks | Endpoint value | No |
| Poukkula 1998 | PEFR in L/min | 12 weeks | Endpoint value | Yes |
| Reichel 2001 | PEFR in L/min | 6 weeks | Endpoint value | Yes |
| Srichana 2016 | PEFR in L/min | 12 weeks | Endpoint value | Yes |
| Stradling 2000 | PEFR in L/min | 12 weeks | Endpoint value | Yes |
| Van Noord 2001 | PEFR in L/min | 12 weeks | Change from baseline | Yes |
| Von Berg 2004 | PEFR in L/min | 4 weeks | Endpoint value | Yes |
| Wardlaw 2004 | PEFR in L/min | 8 weeks | Change from baseline | Yes |
| Wolfe 2000 | PEFR in L/min | 12 weeks | Endpoint value | Yes |
| Zheng 2023 | PEFR in L/min | 12 weeks | Endpoint value | Yes |
| Asthma acute | | | | |
| Khaled 2014 | PEFR in L/min | 30-min post treatment | Endpoint value | Yes |
| Lodha 2004 | PEFR in L/min | 30-min post treatment | Endpoint value | Yes |
| COPD | | | | |
| Ferguson 2013 | PEFR in L/min | 48 weeks | Change from baseline | Yes |
| Koser 2010 | PEFR in L/min | 12 weeks | Change from baseline | Yes |
| **Reliever use** | | | | |
| Asthma Maintenance | | | | |
| Barnes 2013 | rescue medication free days (%) | 12 weeks | Endpoint value | Yes |
| Bateman 2001 | rescue medication free days (median percentage) | 12 weeks | Endpoint value | Yes |
| Bateman 2001 | rescue medication free nights (median percentage) | 12 weeks | Endpoint value | No (not used) |
| Bernstein 2011 | rescue medication free nights (proportion) | 12 weeks | Change from baseline | No |

| | | | | |
|---|---|---|---|---|
| Bodzenta-Lukaszyk 2012 | rescue medication free days | 12 weeks | Change from baseline | No |
| Bracamonte 2005 | rescue medication free nights | 12 weeks | Endpoint value | No (not used) |
| Bracamonte 2005 | rescue medication free days | 12 weeks | Endpoint value | Yes |
| Busse 2008 | rescue medication free days | 7 months | Endpoint value | No (not used) |
| Busse 2008 | Rescue medication use puffs/day | 7 months | Endpoint value | Yes |
| Dusser 2005 | Puffs/day | 12 weeks | Endpoint value | Yes |
| Kanniess 2015 | rescue medication free days | 12 weeks | Change from baseline | No (not used) |
| Kanniess 2015 | Average use of rescue medication (number of inhalations/day) | 12 weeks | Change from baseline | No |
| Koskela 2000 | Number of rescue medication inhalations (number) | 8 weeks | Endpoint value | No |
| Lundback 1993 | rescue medication required days | 6 weeks | Endpoint value | No |
| Lundback 1994 | rescue medication free days (mean) | 4 weeks | Endpoint value | Yes |
| Morice 2007 | rescue medication free days (mean) | 12 weeks | Change from baseline | No (not used) |
| Morice 2007 | Rescue medication use, inhalations/day | 12 weeks | Endpoint value | Yes |
| Papi 2007 | rescue medication free days (%) | 12 weeks | Endpoint value | No (not used) |
| Papi 2007 | Daily use of rescue medication puffs/day | 12 weeks | Endpoint value | Yes |
| Papi 2012 | rescue medication free days at daytime (number of patients) | 6 months | Endpoint value | No |
| Papi 2012 | rescue medication free nights at night-time (number of patients) | 6 months | Endpoint value | No |
| Reichel 2001 | Puffs/day | 6 weeks | Change from baseline | Yes |
| Srichana 2016 | Days of rescue medication used | 12 weeks | Endpoint value | Yes |
| van Noord 2001 | rescue medication free days | 12 weeks | Endpoint value | Yes |
| van Noord 2001 | rescue medication free nights | 12 weeks | Endpoint value | No (not used) |
| Wardlaw 2004 | Puffs +/- nebulizer treatments/day | 8 weeks | Change from baseline | Yes |
| Wolfe 2000 | Puffs / day | 12 weeks | Endpoint value | Yes |
| Wolfe 2000 | rescue medication free days | 12 weeks | Endpoint value | No (not used) |
| Zheng 2023 | rescue medication free days (average) | 12 weeks | Change from baseline | No |
| Asthma acute – no studies | | | | |
| COPD | | | | |
| Ferguson 2018 | Daily use of rescue medication puffs/day | 24 weeks | Change from baseline | Yes |
| Koser 2010 | Daily use of rescue medication puffs/day | 12 weeks | Over the study duration | Yes |
| Wang 2020 | Daily use of rescue medication puffs/day | 24 weeks | Over the study duration | Yes |
| **Exacerbations** | | | | |
| Asthma Maintenance | | | | |
| Amar 2017 | number: not defined | 12 weeks | endpoint | Yes |
| Barnes 2013 | number: not defined | 12 weeks | endpoint | Yes |
| Bateman 2001 | number: not defined | 12 weeks | endpoint | Yes |
| Bernstein 2011 | number: resulting in emergency treatment | 12 weeks | endpoint | Yes |
| Bodzenta-Lukaszyk 2012 | number: resulting in emergency treatment | 12 weeks | endpoint | Yes |

| | | | | |
|---|---|---|---|---|
| Bracamonte 2005 | number: resulting in emergency treatment | 12 weeks | endpoint | Yes |
| Busse 2008 | number: requiring oral corticosteroid treatment | 7 months | endpoint | Yes |
| Dusser 2005 | number: not defined | 12 weeks | endpoint | Yes |
| Kanniess 2015 | number: deterioration in symptoms and lung function | 12 weeks | endpoint | No (not used) |
| Kanniess 2015 | number: needed systemic corticosteroids | 8 weeks | endpoint | Yes |
| Kemp 1989 | number: requiring medical intervention | 12 weeks | endpoint | Yes |
| Koskela 2000 | number: not defined | 8 weeks | endpoint | Yes |
| Morice 2008 | number: requiring medical intervention | 52 weeks | endpoint | Yes |
| Papi 2007 | number | 12 weeks | endpoint | Yes |
| Papi 2012 | number: mild | 24 weeks | endpoint | Yes (combined) |
| Papi 2012 | number: severe | 24 weeks | endpoint | Yes (combined) |
| Stradling 2000 | number: mild | 12 weeks | endpoint | Yes |
| Stradling 2000 | number: moderate | 12 weeks | endpoint | No (not used) |
| Stradling 2000 | number: severe | 12 weeks | endpoint | No (not used) |
| Vincken 2004 | number: not defined | 12 weeks | endpoint | Yes |
| von Berg 2004 | number: not defined | 4 weeks | endpoint | Yes |
| Zheng 2023 | number: not defined | 12 weeks | endpoint | Yes |
| Asthma acute – no studies | | | | |
| COPD | | | | |
| Ferguson 2018 | number: change in usual symptoms beyond normal day-to-day variation | 24 weeks | endpoint | Yes |
| Ferguson 2013 | number: At least one exacerbation | 48 weeks | endpoint | Yes |
| Kilfeather 2004 | number: not defined | 12 weeks | endpoint | Yes |
| Koser 2010 | number: worsening of dyspnea etc. | 12 weeks | endpoint | Yes |
| Maltais 2019 | number: At least one exacerbation | 24 weeks | endpoint | Yes |
| Wang 2020 | number: change in the patient's usual COPD symptoms etc. | 24 weeks | endpoint | Yes |
| Zuwallack 2010 | number: not defined | 12 weeks | endpoint | Yes |
| **Adverse events (AE) / Treatment-related adverse events (trAE) / Serious adverse events (sAE)** | | | | |
| Asthma Maintenance | | | | |
| **Adverse events (AE)** | | | | |
| Amar 2017 | number: Four AEs of clinical interest | 12 weeks | endpoint | Yes |
| Barnes 2013 | number: not defined | 12 weeks | endpoint | Yes |
| Bateman 2001 | number: not defined | 12 weeks | endpoint | Yes |
| Bodzenta-Lukaszyk 2012 | number: not defined | 12 weeks | endpoint | Yes |
| Bracamonte 2005 | number: not defined | 12 weeks | endpoint | Yes |
| Bronsky 1987 | number: not defined | 12 weeks | endpoint | Yes |
| Busse 2008 | number: not defined | 7 months | endpoint | Yes |
| Dusser 2005 | number: any untoward medical occurrences | 12 weeks | endpoint | Yes |
| Koskela 2000 | number: not defined | 8 weeks | endpoint | Yes |

| | | | | |
|---|---|---|---|---|
| Lundback 1993 | number: not defined | 6 weeks | endpoint | Yes |
| Lundback 1994 | number: not defined | 4 weeks | endpoint | Yes |
| Morice 2008 | number: not defined, listed | 52 weeks | endpoint | Yes |
| Morice 2007 | number: not defined | 12 weeks | endpoint | Yes |
| Nelson 1999 | number: not defined | 12 weeks | endpoint | Yes |
| Papi 2007 | number: not defined | 12 weeks | endpoint | Yes |
| Papi 2012 | number: not defined | 24 weeks | endpoint | Yes |
| Reichel 2001 | number: not defined | 6 weeks | endpoint | Yes |
| Srichana 2016 | number: not defined | 12 weeks | endpoint | Yes |
| Stradling 2000 | | | endpoint | Yes |
| Van Noord 2001 | number: not defined | 12 weeks | endpoint | Yes |
| Vincken 2004 | number: not defined | 12 weeks | endpoint | Yes |
| Von Berg 2004 | number: not defined | 4 weeks | endpoint | Yes |
| Zheng 2023 | number: not defined | 12 weeks | endpoint | Yes |
| **Treatment-related adverse events (trAE)** | | | | |
| Amar 2017 | number: determined by the investigator | 12 weeks | endpoint | Yes |
| Bateman 2001 | number: determined by the investigator | 12 weeks | endpoint | Yes |
| Bernstein 2011 | number: Not defined but 'considered related to study therapy' | 12 weeks | endpoint | Yes |
| Bodzenta-Lukaszyk 2012 | number: not defined | 12 weeks | endpoint | Yes |
| Bracamonte 2005 | number: not defined | 12 weeks | endpoint | Yes |
| Dusser 2005 | number: any untoward medical occurrences | | endpoint | Yes |
| Kanniess 2015 | number: not defined | 8 weeks | endpoint | Yes |
| Morice 2008 | number: not defined, listed | 52 weeks | endpoint | Yes |
| Morice 2007 | number: not defined, listed | 12 weeks | endpoint | Yes |
| Poukkula 1998 | number of events | 12 weeks | endpoint | Yes |
| Reichel 2001 | number: not defined | 6 weeks | endpoint | Yes |
| Van Noord 2001 | number: not defined | 12 weeks | endpoint | Yes |
| Wardlaw 2004 | number: Any event documented by patient and assessed by the physician | 8 weeks | endpoint | Yes |
| Wolfe 2000 | number: considered by the investigators | 12 weeks | endpoint | Yes |
| Zheng 2023 | number: considered by the investigators | 12 weeks | endpoint | Yes |
| **Treatment-related serious/severe adverse events (trsAE)** | | | | |
| Bateman 2001 | number: Any event which was fatal, life-threatening, disabling or incapacitating, or which required or prolonged hospitalization | 12 weeks | endpoint | Yes |
| Bernstein 2011 | number: not defined | 12 weeks | endpoint | Yes |
| Bodzenta-Lukaszyk 2012 | number: not defined | 12 weeks | endpoint | Yes |
| Bracamonte 2005 | number: not defined | 12 weeks | endpoint | Yes |

| | | | | |
|---|---|---|---|---|
| Busse 2008 | number: Adverse effects that resulted in death, were immediately life-threatening, required hospitalization, or resulted in significant disability or incapacity | 12 weeks | endpoint | Yes |
| Dusser 2005 | number: any untoward medical occurrences | 12 weeks | endpoint | Yes |
| Nelson 1999 | number: not defined | 12 weeks | endpoint | Yes |
| Van Noord 2001 | number: any event that was fatal, life-threatening, disabling or incapacitating, or which required or prolonged hospitalisation | 12 weeks | endpoint | Yes |
| Vincken 2004 | number: not defined | 12 weeks | endpoint | Yes |
| Von Berg 2004 | number: not defined | 4 weeks | endpoint | Yes |
| Zheng 2023 | number: considered by the investigators | 12 weeks | endpoint | Yes |
| **Serious adverse events (sAE)** | | | | |
| Amar 2017 | number: not defined | 12 weeks | endpoint | Yes |
| Barnes 2013 | number: not defined | 12 weeks | endpoint | Yes |
| Bateman 2001 | number: Any event which was fatal, life-threatening, disabling or incapacitating, or which required or prolonged hospitalization | 12 weeks | endpoint | Yes |
| Bernstein 2011 | number: not defined | 12 weeks | endpoint | Yes |
| Bodzenta-Lukaszyk 2012 | number: not defined | 12 weeks | endpoint | Yes |
| Bracamonte 2005 | number: not defined | 12 weeks | endpoint | Yes |
| Bronsky 1987 | number: not defined | 12 weeks | endpoint | Yes |
| Busse 2008 | number: Adverse effects that resulted in death, were immediately life-threatening, required hospitalization, or resulted in significant disability or incapacity | 7 months | endpoint | Yes |
| Dusser 2005 | number: any untoward medical occurrences | 12 weeks | endpoint | Yes |
| Koskela 2000 | number: not defined | 8 weeks | endpoint | Yes |
| Lundback 1994 | number: not defined | 4 weeks | endpoint | Yes |
| Morice 2008 | number: not defined, listed | 52 weeks | endpoint | Yes |
| Morice 2007 | number: not defined | 12 weeks | endpoint | Yes |
| Nelson 1999 | number: not defined | 12 weeks | endpoint | Yes |
| Papi 2007 | number: not defined | 12 weeks | endpoint | Yes |
| Papi 2012 | number: not defined | 24 weeks | endpoint | Yes |
| Reichel 2001 | number: not defined | 6 weeks | endpoint | Yes |
| Van Noord 2001 | number: any event that was fatal, life-threatening, disabling or incapacitating, or which required or prolonged hospitalisation | 12 weeks | endpoint | Yes |
| Vincken 2004 | number: not defined | 12 weeks | endpoint | Yes |
| Von Berg 2004 | number: not defined | 4 weeks | endpoint | Yes |
| Wardlaw 2004 | number: not defined | 8 weeks | endpoint | Yes |
| Zheng 2023 | number: not defined | 12 weeks | endpoint | Yes |
| Asthma acute | | | | |
| **Adverse events (AE)** | | | | |

| | | | | |
|---|---|---|---|---|
| Vangveeravong 2008 | number: Tremor or palpitations during treatment | 60 min | endpoint | Yes |
| COPD | | | | |
| **Adverse events (AE)** | | | | |
| Ferguson 2013 | number: not defined | 48 weeks | endpoint | Yes |
| Ferguson 2018 | number: not defined | 24 weeks | endpoint | Yes |
| Kilfeather 2004 | number: not defined | 12 weeks | endpoint | Yes |
| Koser 2010 | number: not defined, listed | 12 weeks | endpoint | Yes |
| Maltais 2019 | number: not defined, listed | 24 weeks | endpoint | Yes |
| Wang 2020 | number: not defined | 24 weeks | endpoint | Yes |
| Zuwallack 2010 | number: not defined, listed | 12 weeks | endpoint | Yes |
| **Serious adverse events (sAE)** | | | | |
| Ferguson 2013 | number: not defined | 48 weeks | endpoint | Yes |
| Ferguson 2018 | number: not defined | 24 weeks | endpoint | Yes |
| Maltais 2019 | number: not defined | 24 weeks | endpoint | Yes |
| Wang 2020 | number: not defined | 24 weeks | endpoint | Yes |
| Zuwallack 2010 | number: not defined, listed | 12 weeks | endpoint | Yes |
| **Treatment-related adverse events (trAE)** | | | | |
| Ferguson 2018 | number: not defined | 24 weeks | endpoint | Yes |
| Maltais 2019 | number: investigator determined | 24 weeks | endpoint | Yes |
| Wang 2020 | number: investigator determined | 24 weeks | endpoint | Yes |
| **Treatment-related serious adverse events (trsAE)** | | | | |
| Ferguson 2018 | number: not defined | 24 weeks | endpoint | Yes |
| Wang 2020 | number: investigator determined | 24 weeks | endpoint | Yes |
| **Asthma control (symptom control)** | | | | |
| Asthma Maintenance | | | | |
| Barnes 2013 | ACQ-7 | 12 weeks | change from baseline | Yes |
| Bateman 2001 | Symptom free days | 12 weeks | endpoint | No |
| Bateman 2001 | Symptom free nights | 12 weeks | endpoint | No |
| Bernstein 2011 | ACQ score | 4 weeks | endpoint | No (not used) |
| Bernstein 2011 | ACQ score | 12 weeks | endpoint | Yes |
| Bodzenta-Lukaszyk 2012 | asthma control days (%) | 12 weeks | change from baseline | Yes |
| O'Connor 2010 | ACQ score | 7 months | endpoint | Yes |
| Kanniess 2015 | ACQ-7 | 8 weeks | change from baseline | Yes |
| Lundback 1994 | Median day symptom score $\geq 2$ | 4 weeks | endpoint | No |
| Lundback 1994 | Median night symptom score $\geq 2$ | 4 weeks | endpoint | No |
| Morice 2007 | Asthma control days (%) | 12 weeks | change from baseline | No |

| | | | | |
|---|---|---|---|---|
| Papi 2012 | Asthma control defined using a composite | 6 months | endpoint | No |
| Poukkula 1998 | Severity sum scores | 4 weeks | endpoint | No |
| Poukkula 1998 | Severity sum scores | 8 weeks | endpoint | No |
| Poukkula 1998 | Severity sum scores | 12 weeks | endpoint | No |
| Srichna 2016 | Symptom free days | 90 days | endpoint | No |
| Van Noord 2001 | Symptom free days | 12 weeks | endpoint | No |
| Wolfe 2000 | Self-rated asthma symptom score | 12 weeks | change from baseline | Yes |
| Zheng 2023 | ACQ-6 | 12 weeks | change from baseline | Yes |
| Asthma acute | | | | |
| Vangveeravong 2008 | Modified Wood Clinical Asthma Score | 20 min | endpoint | No (not used) |
| Vangveeravong 2008 | Modified Wood Clinical Asthma Score | 40 min | endpoint | No (not used) |
| Vangveeravong 2008 | Modified Wood Clinical Asthma Score | 60 min | endpoint | Yes |
| Direkwatanachai 2011 | Modified Wood Clinical Asthma Score<br>Either a score falling by >50% from baseline, or below raw score of 3 (number of patients) | 60 min | endpoint | No |
| COPD | | | | |
| Maltais 2019 | CAT score | 24 weeks | endpoint | Yes |
| **Quality of life** | | | | |
| Asthma Maintenance | | | | |
| Amar 2017 | PAQLQ[S]) | 12 weeks | Change from baseline | No |
| Bernstein 2011 | AQLQ[S] | 4 weeks | Endpoint | No |
| Bernstein 2011 | AQLQ[S] | 12 weeks | Endpoint | No |
| Bodzenta-Lukaszyk 2012 | Proportion of responders | 12 weeks | Endpoint | No |
| Bodzenta-Lukaszyk 2012 | AQLQ | 12 weeks | Endpoint | No |
| O'Connor 2010 | AQLQ[S]<br>Improvements (≥0.5) in overall score from baseline (number of patients) | 7 months | Change from baseline | Yes |
| Koskela 2000 | SGRQ, Parts I–II | 8 weeks | Endpoint | No |
| Morice 2007 | AQLQ[S] | 12 weeks | Change from baseline | No |
| Morice 2007 | AQLQ[S]<br>Improvements (≥0.5) in overall score from baseline (number of patients) | 12 weeks | Change from baseline | Yes |
| Asthma acute – no studies | | | | |
| COPD | | | | |
| Ferguson 2018 | SGRQ | 24 weeks | Endpoint | No |
| Wang 2020 | SGRQ | 12-24 weeks | Change from baseline | No |

| | | | | |
|---|---|---|---|---|
| Wang 2020 | TDI Score | 12-24 weeks | Change from baseline | No |

#### Table S16 – Excluded studies that might appear to meet inclusion criteria

| Number | Citation | Reason for exclusion |
|---|---|---|
| 1 | Ali Rizvi, D., M. Tariq Salman, J. Sircar and A. Ahmad (2013). "Effect of Budesonide by metered dose inhaler with or without spacer & dry powder inhaler on Lung Function." International Journal of Drug Development and Research **5**(4): 233-240. | Wrong outcomes |
| 2 | Bensch, G., R. J. Lapidus, B. E. Levine, W. Lumry, U. Yegen, P. Kiselev and G. Della Cioppa (2001). "A randomized, 12-week, double-blind, placebo-controlled study comparing formoterol dry powder inhaler with albuterol metered-dose inhaler." Ann Allergy Asthma Immunol **86**(1): 19-27. | Wrong comparator |
| 3 | Berger, R. and W. E. Berger (2013). "Particle size and small airway effects of mometasone furoate delivered by dry powder inhaler." Allergy Asthma Proc **34**(1): 52-58. | Wrong outcomes |
| 4 | Bogdan, M. A., H. Aizawa, Y. Fukuchi, M. Mishima, M. Nishimura and M. Ichinose (2011). "Efficacy and safety of inhaled formoterol 4.5 and 9 mug twice daily in Japanese and European COPD patients: Phase III study results." BMC Pulmonary Medicine **11**: 51. | Wrong comparator |
| 5 | Bulac, S., A. Cimrin and H. Ellidokuz (2015). "The effect of beclometasone dipropionate/formoterol extra-fine fixed combination on the peripheral airway inflammation in controlled asthma." J Aerosol Med Pulm Drug Deliv **28**(2): 82-87. | Wrong outcomes |
| 6 | Calverley, P. M., P. Kuna, E. Monsó, M. Costantini, S. Petruzzelli, F. Sergio, G. Varoli, A. Papi and V. Brusasco (2010). "Beclomethasone/formoterol in the management of COPD: a randomised controlled trial." Respir Med **104**(12): 1858-1868. | Wrong dose |
| 7 | Capanoglu, M., E. Dibek Misirlioglu, M. Toyran, E. Civelek and C. N. Kocabas (2015). "Evaluation of inhaler technique, adherence to therapy and their effect on disease control among children with asthma using metered dose or dry powder inhalers." J Asthma **52**(8): 838-845. | Wrong outcomes |
| 8 | Chang, T. Y., J. Y. Chien, C. H. Wu, Y. H. Dong and F. J. Lin (2019). "Comparative Safety and Effectiveness of Inhaled Corticosteroids and Long-Acting beta2 Agonist Combinations in Patients with Chronic Obstructive Pulmonary Disease." Chest **157**(5): 1117-1129. | Wrong comparator |
| 9 | Chapman, K. R., K. Friberg, M. S. Balter, R. H. Hyland, M. Alexander, R. T. Abboud, S. Peters and B. H. Jennings (1997). "Albuterol via Turbuhaler versus albuterol via pressurized metered-dose inhaler in asthma." Ann Allergy Asthma Immunol **78**(1): 59-63. | Wrong outcomes |
| 10 | Chuchalin, A. G., H. J. Kremer, P. Metzenauer, E. O'Keefe and R. Hermann (2002). "Clinical equivalence trial on budesonide delivered either by the Novolizer multidose dry powder inhaler or the Turbuhaler in asthmatic patients." Respiration **69**(6): 502-508. | Wrong comparator |
| 11 | Corren, J., P. E. Korenblat, C. J. Miller, C. D. O'Brien and W. S. Mezzanotte (2007). "Twelve-week, randomized, placebo-controlled, multicenter study of the efficacy and tolerability of budesonide and formoterol in one metered-dose inhaler compared with budesonide alone and formoterol alone in adolescents and adults with asthma." Clin Ther **29**(5): 823-843. | Wrong comparator |
| 12 | Crompton, G. K., R. Sanderson, M. H. Dewar, S. P. Matusiewicz, A. C. Ning, A. H. Jamieson, A. McLean and A. P. Greening (2000). "Comparison of Pulmicort pMDI plus Nebuhaler and Pulmicort Turbuhaler in asthmatic patients with dysphonia." Respir Med **94**(5): 448-453. | Wrong outcomes |
| 13 | Cuvelier, A., J. F. Muir, D. Benhamou, E. Weitzenblum, P. Zuck, R. Delacenserie, A. Taytard and P. Iacono (2002). "Dry powder ipratropium bromide is as safe and effective as metered-dose inhaler formulation: a cumulative dose-response study in chronic obstructive pulmonary disease patients." Respir Care **47**(2): 159-166. | Wrong outcomes |
| 14 | Du, Y., W. Wang, W. Yang and B. He (2014). "Interleukin-32, not reduced by salmeterol/fluticasone propionate in smokers with chronic obstructive pulmonary disease." Chin Med J (Engl) **127**(9): 1613-1618. | Wrong outcomes |
| 15 | Ferguson, G. T., N. Brown, C. Compton, T. C. Corbridge, K. Dorais, C. Fogarty, C. Harvey, M. C. Kaisermann, D. A. Lipson, N. Martin, F. Sciurba, M. Stiegler, C. Q. Zhu and D. Bernstein (2020). "Once-daily single-inhaler versus twice-daily multiple-inhaler triple therapy in patients with COPD: lung function and health status results from two replicate randomized controlled trials." Respir Res **21**(1): 131. | Wrong comparator |
| 16 | Giraud, V. and F. A. Allaert (2009). "Improved asthma control with breath-actuated pressurized metered dose inhaler (pMDI): the SYSTER survey." Eur Rev Med Pharmacol Sci **13**(5): 323-330. | Wrong comparator |

| | | |
|---|---|---|
| 17 | Hirsch, T., M. Peter-Kern, R. Koch and W. Leupold (1997). "Influence of inspiratory capacity on bronchodilatation via Turbuhaler or pressurized metered-dose inhaler in asthmatic children: a comparison." Respir Med **91**(6): 341-346. | Wrong outcomes |
| 18 | Horiguchi, T., N. Hayashi, D. Ohira, H. Torigoe, T. Ito, M. Hirose, Y. Sasaki, M. Shiga, J. Miyazaki, R. Kondo and S. Tachikawa (2006). "Usefulness of HFA-BDP for adult patients with bronchial asthma: randomized crossover study with fluticasone." J Asthma **43**(7): 509-512. | Wrong comparator |
| 19 | Huber, B., C. Keller, M. Jenkins, A. Raza and M. Aurivillius (2022). "Effect of inhaled budesonide/formoterol fumarate dihydrate delivered via two different devices on lung function in patients with COPD and low peak inspiratory flow." Ther Adv Respir Dis **16**: 17534666221107312. | Wrong outcomes |
| 20 | Ige, O. M. and O. M. Sogaolu (2004). "A single blinded randomised trial to compare the efficacy and safety of once daily budesonide (400microg) administered by turbuhaler with beclomethasone dipropionate (400microg) given twice daily through a metered-dose inhaler in patients with mild to moderate asthma." Afr J Med Med Sci **33**(2): 155-160. | Wrong dose |
| 21 | Kawai, M., A. Sakai, S. Takaori, A. Hiura, N. Sakata, M. Nakashima and T. Miyamoto (2005). "Pharmacodynamic study of procaterol hydrochloride dry powder inhaler: evaluation of pharmacodynamic equivalence between procaterol hydrochloride dry powder inhaler and procaterol hydrochloride metered-dose inhaler in asthma patients in a randomized, double-dummy, double-blind crossover manner." Methods Find Exp Clin Pharmacol **27**(6): 385-389. | Wrong outcomes |
| 22 | Kerwin, E., A. Wachtel, L. Sher, J. Nyberg, P. Darken, S. Siddiqui, E. A. Duncan, C. Reisner and P. Dorinsky (2018). "Efficacy, safety, and dose response of glycopyrronium administered by metered dose inhaler using co-suspension delivery technology in subjects with intermittent or mild-to-moderate persistent asthma: A randomized controlled trial." Respir Med **139**: 39-47. | Wrong comparator |
| 23 | Kolasani, B. P., V. M. Lanke and S. Diyya (2013). "Influence of delivery devices on efficacy of inhaled fluticasone propionate: a comparative study in stable asthma patients." J Clin Diagn Res **7**(9): 1908-1912. | Wrong outcomes |
| 24 | Kostikas, K., J. F. Maspero, K. R. Chapman, K. Mezzi, X. Jaumont, D. Lawrence and R. van Zyl-Smit (2023). "Efficacy of mometasone/indacaterol/glycopyrronium in patients with inadequately controlled asthma with respect to baseline eosinophil count: Post hoc analysis of IRIDIUM study." Respiratory Medicine **217**: 107334. | Wrong comparator |
| 25 | Kraszko, P., D. Vondra, J. Malolepszy, M. Svensson, J. Baly, J. Fiserova, J. Jirkal, K. Kalandrova, P. Pancner, I. Michl, E. Ohnutkova, L. Pavelkova, P. Petrik, T. Sykora, R. Vodrazka, D. Chvatalova, G. Berta, B. Gautier, G. B. Nagy, I. Tallosy, Z. Gonczi, G. Czerniawska-Mysik, T. Hofman, R. Sopel, M. Szmidt and M. L. Kowalski (1999). "Budesonide via Turbuhaler, 400 mug daily, is as effective as beclomethasone dipropionate via pressurised MDI, 800 mug daily, for control of mild to moderate asthmatic patients." Journal of Clinical Research **2**(47-55): 47-55. | Wrong dose |
| 26 | Kupczyk, M., P. Majak, P. Kuna, B. Asankowicz-Bargiel, E. Barańska, R. Dobek, S. Garbicz, J. Jerzyńska, A. Latos, W. Machowiak, B. Majorek-Olechowska, A. Olech-Cudzik, I. Poziomkowska-Gęsicka, M. Rulewicz-Warniełło, A. Świderska, M. Tarnowski and P. Kopyto (2021). "A new formulation of fluticasone propionate/salmeterol in a metered-dose inhaler (MDI HFA) allows for the reduction of a daily dose of corticosteroid and provides optimal asthma control - A randomized, multi-center, non-inferiority, phase IV clinical study." Respir Med **176**: 106274. | Wrong dose |
| 27 | LaForce, C., B. M. Prenner, K. Andriano, C. Lavecchia and U. Yegen (2005). "Efficacy and safety of formoterol delivered via a new multidose dry powder inhaler (Certihaler) in adolescents and adults with persistent asthma." J Asthma **42**(2): 101-106. | Wrong comparator |
| 28 | Lal, S., S. M. Malhotra, M. D. Gribben and A. G. Butler (1980). "Beclomethasone dipropionate aerosol compared with dry powder in the treatment of asthma." Clin Allergy **10**(3): 259-262. | Wrong comparator |
| 29 | Löfdahl, C. G., L. Andersson, E. Bondesson, L. G. Carlsson, K. Friberg, J. Hedner, Y. Hörnblad, P. Jemsby, A. Källén, A. Ullman, S. Werner and N. Svedmyr (1997). "Differences in bronchodilating potency of salbutamol in Turbuhaler as compared with a pressurized metered-dose inhaler formulation in patients with reversible airway obstruction." Eur Respir J **10**(11): 2474-2478. | Wrong outcomes |
| 30 | McCarthy, P., T. Iliadis and K. Zaiken (2022). "Clinical Response and Cost-Savings Associated With Generic Fluticasone | Wrong comparator |

| | | |
|---|---|---|
| | Propionate/Salmeterol Multidose, Dry-Powder Inhaler in Asthma Patients Managed in an Ambulatory Care Practice Setting." J Pharm Pract **35**(2): 274-280. | |
| 31 | Miyamoto, T., T. Takahashi, S. Nakajima, S. Makino, M. Yamakido, K. Mano, M. Nakashima, U. Tollemar and O. Selroos (2001). "Efficacy of budesonide Turbuhaler compared with that of beclomethasone dipropionate pMDI in Japanese patients with moderately persistent asthma." Respirology **6**(1): 27-35. | Wrong dose |
| 32 | Noonan, M., L. J. Rosenwasser, P. Martin, C. D. O'Brien and L. O'Dowd (2006). "Efficacy and safety of budesonide and formoterol in one pressurised metered-dose inhaler in adults and adolescents with moderate to severe asthma: a randomised clinical trial." Drugs **66**(17): 2235-2254. | Wrong comparator |
| 33 | O'Callaghan, C., M. L. Everard, A. Bush, E. J. Hiller, R. Ross-Russell, P. O'Keefe and P. Weller (2002). "Salbutamol dry powder inhaler: efficacy, tolerability, and acceptability study." Pediatr Pulmonol **33**(3): 189-193. | Wrong outcomes |
| 34 | Ohaju-Obodo, J. O., C. Chukwu, J. Okpapi, E. Egbagbe, M. O. Ige, C. Chukwuka and D. A. Adedapo (2005). "Comparison of the efficacy and safety of budesonide turbuhaler administered once daily with twice the dose of beclomethasone dipropionate using pressurised metered dose inhaler in patients with mild to moderate asthma." West Afr J Med **24**(3): 190-195. | Wrong dose |
| 35 | Ohbayashi, H., S. Kudo and M. Ariga (2018). "Evaluation of Rapid Onset of Action of ICS/LABA Combination Therapies on Respiratory Function in Asthma Patients: A Single-Center, Open-Label, Randomized, Crossover Trial." Pulm Ther **4**(2): 159-169. | Wrong outcomes |
| 36 | Piquet, J., P. Zuck, G. Dennewald, P. Dugue, M. Grivaux, P. Brun, J. C. Severac, J. Ostinelli and K. H. Cheeseman (1996). "Equally efficacious asthma management with budesonide 800 micrograms administered by Turbuhaler or with beclomethasone dipropionate > or = 1500 micrograms given through a pressurized metered-dose inhaler with spacer. The French Budesonide Trial Group." Adv Ther **13**(1): 38-50. | Wrong dose |
| 37 | Price, D., V. Thomas, J. von Ziegenweidt, S. Gould, C. Hutton and C. King (2014). "Switching patients from other inhaled corticosteroid devices to the Easyhaler(®): historical, matched-cohort study of real-life asthma patients." J Asthma Allergy **7**: 31-51. | Wrong comparator |
| 38 | Razzouk, H., L. dos Santos, J. Giudicelli, M. Queirós, M. de Lurdes Chieira, A. Castro, C. Ramos and C. Lindbladh (1999). "A comparison of the bronchodilatory effect of 50 and 100 microg salbutamol via Turbuhaler and 100 microg salbutamol via pressurized metered dose inhaler in children with stable asthma." Int J Pharm **180**(2): 169-175. | Wrong outcomes |
| 39 | Rodrigo, G. J., H. Neffen, F. D. Colodenco and J. A. Castro-Rodriguez (2010). "Formoterol for acute asthma in the emergency department: a systematic review with meta-analysis." Ann Allergy Asthma Immunol **104**(3): 247-252. | Wrong comparator |
| 40 | Salvi, S., A. K. Deb, M. Agarwal, V. R. Tummuru, R. Kodgule, V. S. Hemalatha, A. K. Awasthi, K. P. Suraj, V. K. Pavitran, S. P. Mourya, P. Thomas, A. Vaidya, S. Chhowala and J. Gogtay (2020). "Fixed-dose combination of three drugs, i.e. LABA/LAMA/ICS for COPD: Results of a real-world study from India." Pulm Pharmacol Ther **63**: 101932. | Wrong comparator |
| 41 | Schurmann, W., S. Schmidtmann, P. Moroni, D. Massey and M. Qidan (2005). "Respimat Soft MistTM inhaler versus hydrofluoroalkane metered dose inhaler: Patient preference and satisfaction." Treatments in Respiratory Medicine **4**(1): 53-61. | Wrong dose |
| 42 | Selroos, O., R. Backman, K. O. Forsen, A. B. Lofroos, M. Niemisto, A. Pietinalho and H. Riska (1994). "Clinical efficacy of budesonide Turbuhalerregistered trade mark compared with that of beclomethasone dipropionate pMDI with volumatic spacer. A 2-year randomized study in 102 asthma patients." ALLERGY-EUR-J-ALLERGY-CLIN-IMMUNOL **49**(10): 833-836. | Wrong comparator |
| 43 | Singh, D., G. Nicolini, E. Bindi, M. Corradi, D. Guastalla, J. Kampschulte, W. Pierzchala, A. Sayiner, M. Szilasi, C. Terzano and J. Vestbo (2014). "Extrafine beclomethasone/formoterol compared to fluticasone/salmeterol combination therapy in COPD." BMC Pulmonary Medicine **14**(1): 43. | Wrong dose |
| 44 | Tamási, L., M. Szilasi and G. Gálffy (2018). "Clinical Effectiveness of Budesonide/Formoterol Fumarate Easyhaler(®) for Patients with Poorly Controlled Obstructive Airway Disease: a Real-World Study of Patient-Reported Outcomes." Adv Ther **35**(8): 1140-1152. | Wrong comparator |
| 45 | Tammivaara, R., E. Aalto, K. Lehtonen, V. Vilkka, K. Laurikainen, M. Silvasti, P. Toivanen and H. Tukiainen (1997). "Comparison of a novel salbutamol multidose powder inhaler with a salbutamol metered dose inhaler in patients with asthma." Current Therapeutic Research - | Wrong comparator |

| | | |
|---|---|---|
| | Clinical and Experimental **58**(10): 734-744. | |
| 46 | van Geffen, W. H., W. R. Douma, D. J. Slebos and H. A. Kerstjens (2016). "Bronchodilators delivered by nebuliser versus pMDI with spacer or DPI for exacerbations of COPD." Cochrane Database Syst Rev **2016**(8): Cd011826. | Wrong comparator |
| 47 | Vidgren, P., M. Silvasti, A. Poukkula, K. Laasonen and M. Vidgren (1994). "Easyhaler powder inhaler - A new alternative in the anti-inflammatory treatment of asthma." Acta Therapeutica **20**(3-4): 117-131. | Wrong outcomes |
| 48 | Vinge, I., J. Syk, A. Xanthopoulos, H. Laßmann, M. Vahteristo, U. Sairanen, S. Lähelmä, R. Hennig and M. Müller (2021). "A non-interventional switch study in adult patients with asthma or COPD on clinical effectiveness of salmeterol/fluticasone Easyhaler(®) in routine clinical practice." Ther Adv Respir Dis **15**: 17534666211027787. | Wrong comparator |
| 49 | Voshaar, T., R. Lapidus, R. Maleki-Yazdi, W. Timmer, E. Rubin, L. Lowe and E. Bateman (2008). "A randomized study of tiotropium Respimat Soft Mist inhaler vs. ipratropium pMDI in COPD." Respiratory Medicine **102**(1): 32-41. | Wrong comparator |
| 50 | Woodcock, A., C. Janson, J. Rees, L. Frith, M. Löfdahl, A. Moore, M. Hedberg and D. Leather (2022). "Effects of switching from a metered dose inhaler to a dry powder inhaler on climate emissions and asthma control: post-hoc analysis." Thorax **77**(12): 1187-1192. | Wrong dose |
| 51 | Worth, H., J. F. Muir and W. R. Pieters (2001). "Comparison of hydrofluoroalkane-beclomethasone dipropionate AutohalerTM with budesonide TurbuhalerTM in asthma control." Respiration **68**(5): 517-526. | Wrong dose |
| 52 | Zheng, J. P., L. Yang, Y. M. Wu, P. Chen, Z. G. Wen, W. J. Huang, Y. Shi, C. Z. Wang, S. G. Huang, T. Y. Sun, G. F. Wang, S. D. Xiong and N. S. Zhong (2007). "The efficacy and safety of combination salmeterol (50 microg)/fluticasone propionate (500 microg) inhalation twice daily via accuhaler in Chinese patients with COPD." Chest **132**(6): 1756-1763. | Wrong comparator |
